# Predicting the causal relationship between polyunsaturated fatty acids and cerebral aneurysm risk from a Mendelian randomization study

**DOI:** 10.1101/2023.11.15.23298570

**Authors:** Weijie Yu, Liwei Zhou, Chongfei Li, Zhenwei Lu, Xiaoyu Chen, Hao Yu, Xiaoyan Chen, Qionghui Huang, Zhangyu Li, Deyong Xiao, Yunyun Mei, Zhanxiang Wang

**Affiliations:** Department of Neurosurgery, The First Affiliated Hospital of Fujian Medical University, Fuzhou, China; The School of Clinical Medicine, Fujian Medical University, Fuzhou 350122, China; Department of Neurosurgery, Department of Neurosurgery Quality Control Center, The First Affiliated Hospital of Xiamen University, Xiamen Key Laboratory of Brain Center, Xiamen 361003, China; School of Medicine, Xiamen University, Xiamen 361005, China; Internal medicine, The First Affiliated Hospital of Xiamen University, Xiamen 361003, China; Department of Anesthesiology, The First Affiliated Hospital of Xiamen University, Xiamen Key Laboratory of Brain Center, Xiamen 361003, China; Department of Neurosurgery, Fudan University Shanghai Cancer Center Xiamen Hospital, Xiamen 361026, China

**Author notes:** **Co-first author** Weijie Yu, Liwei Zhou and Chongfei Li contributed equally as the first authors. **Corresponding Author** Yunyun Mei, MS. Department of Neurosurgery, Department of Neurosurgery, Fudan University Shanghai Cancer Center Xiamen Hospital, 99 Dongfu West Road, Haicang District, Xiamen City, Fujian, 361003, P.R. China.; Zhanxiang Wang, MD. Department of Neurosurgery, The First Affiliated Hospital of Xiamen University, Xiamen Key Laboratory of Brain Center, 55 Zhenhai Road, Xiamen Siming District, Xiamen City, Fujian, 361003, P.R. China.

**Keywords:** polyunsaturated fatty acids, intracranial aneurysm, subarachnoid hemorrhage, Mendelian randomization

## Abstract

**Introduction:** No conclusive evidence for a link between polyunsaturated fatty acids (PUFA) and cerebral aneurysm has been found in observational research. The aim of our study was to determine the causal impact of PUFA on cerebral aneurysm.

**Methods:** Two sample Mendelian randomization (MR) was performed using genetic instruments derived from a recent genome wide association study (GWAS) of fatty acids from UK Biobank and outcome data obtained from the large-scale cerebral aneurysm GWASs in European ancestry which include IA, aneurysmal subarachnoid hemorrhage (aSAH) and unruptured intracranial aneurysm (uIA). Sensitivity analyses were implemented with MVMR, MR-Egger intercept test, MR-PRESSO, leave-one-out analysis and so on. Bayesian colocalization (COLOC) methods was conducted to focus on the association between the fatty acid gene expression and cerebral aneurysm.

**Results:** Genetically predicted assessed omega-3 fatty acids decreased the risk for IA (OR = 0.80, 95% CI: 0.69 - 0.91, P = 1.01ⅹ10^-3^) and aSAH (OR = 0.71, 95% CI: 0.61 - 0.84, P = 3.73ⅹ10^-5^). Furthermore, the Docosahexaenoic acid decreased the risk for IA (OR = 0.75, 95% CI: 0.63 - 0.87, P = 3.12ⅹ10^-4^) and aSAH (OR = 0.67, 95% CI: 0.55 - 0.8, P = 2.32ⅹ10^-5^). The same results were discovered from ratio of omega-3 fatty acids to total fatty acids. While the ratio of omega-6 fatty acids to omega-3 fatty acids increased the risk of IA (OR = 1.27, 95% CI: 1.12 – 1.44, P = 1.53ⅹ10^-4^) and aSAH (OR = 1.35, 95% CI: 1.17 – 1.56, P = 5.78ⅹ10^-5^). The result of the COLOC suggested that the above four kinds of fatty acids and IA, aSAH likely share causal variants in gene fatty acid desaturase 2, separately.

**Conclusion:** This study utilized integrative analysis of MR and colocalization to discover causal relationships between genetic variants, PUFA and cerebral aneurysm.

**Funding:** This study was funded by the Natural Science Foundation of China (82072777), the Natural Science Foundation of Xiamen (3502Z20227097), Fujian Provincial Health Commission, Provincial Health and Health Young and Middle-aged Backbone Talent Training Project (2022GGB010).

## Introduction

Intracranial aneurysm (IA) is confined, pathological dilatations of the walls of intracranial arteries, which currently considered a chronic inflammatory disease that affects the intracranial arteries (Aoki et al., 2017) with a prevalence of approximately 3% in the general population (Vlak et al., 2011). About 85% of spontaneous subarachnoid hemorrhage (SAH) is due to ruptured IA (Macdonald et al., 2017). Unruptured IAs (uIAs) are frequently asymptomatic, but IA rupture can result in aneurysmal subarachnoid hemorrhage (aSAH), which has a poor prognosis (30% of death, 30% of independence and 30% of dependence), with patients often suffering from a disability or even death (Chen et al., 2020; (Nieuwkamp et al., 2009). In light of the elevated rates of mortality and morbidity brought on by IA, it is crucial to explore the pathogenesis of intracranial aneurysm.

Polyunsaturated fatty acids (PUFA) are fatty acids that contain multiple double bonds in their molecular structure. According to the position and number of double bonds, polyunsaturated fatty acids can be further divided into two categories, omega-3 and omega-6. The long-chain omega-3 polyunsaturated fatty acid (omega-3) and docosahexaenoic acid (DHA) have been suggested to have cardioprotective (Burr et al., 1989; (Wang et al., 2006), anti-inflammatory (Massaro et al., 2006), immunoregulatory (Yang et al., 2013), antioxidant (Carrepeiro et al., 2011; (Bouzidi et al., 2010) and anti-tumor activities (Calviello et al., 2006). A recent study reported a significant decrease in red blood cell distribution width, pulse wave analysis and heart rate in patients with abdominal aortic aneurysm who were supplemented with 1.8 g of long-chain omega-3 polyunsaturated fatty acids for 12 weeks, suggesting an improving in intravascular inflammation and vascular stiffness (Meital et al., 2019; (Meital et al., 2020). Omega-3 and DHA down-regulate multiple aspects of the inflammatory process, such as oxidative stress and inflammation in macrophages (Meital et al., 2019). According to a recent retrospective study, high levels of free fatty acids on admission can provide some reference predictions for the poor prognosis of patients with aSAH at 3 months (Wang et al., 2021). Other studies demonstrated that arachidonic acid (AA) and AA-containing phospholipids were not detected in unruptured IA walls, whereas AA and AA-containing phospholipids were detected in significantly higher amounts in ruptured IA walls than in unruptured IA walls (Takeda et al., 2021).

Mendelian randomization (MR) is the use of genetic variation in non-experimental data to estimate the causal link between exposure and outcome, and it can reduce the impact of behavioral, social, psychological, and other factors (Davey Smith et al., 2014). And in recent years, many MR studies have emerged to provide clinical evidence (Hu et al., 2023; (Yuan et al., 2019). To investigate the etiology of cerebral aneurysms, it is essential to further clarification of the causal relationship between PUFAs and cerebral aneurysms through Mendelian randomization.

## Methods

### Study design

In this study, we performed a two-sample Mendelian randomization analysis to examine the causal effects of PUFA and cerebral aneurysms using genome wide association study (GWAS) summary statistics. This instrumental-variable analysis mimics RCT with respect to the random allocation of single nucleotide polymorphisms (SNPs) in offspring (independent of confounding factors such as sex and age). In addition, this MR design has to fulfill three assumptions: (i) genetic instruments predict the exposure of interest (P < 5×10–8); (ii) genetic instruments are independent of potential confounders; (iii) genetic instruments affects the outcome only via the risk factors (Boef et al., 2015). The MR analysis used summary GWAS data publicly available from GWASs and no individual level data used. Due to the fact that each of the original GWASs had obtained ethical approval and participant consent, they were not required.

### Data source and selection of genetic instruments

SNPs associated with 5 PUFAs were obtained from the open GWAS website, containing omega-3 fatty acids, omega-6 fatty acids, DHA, the ratio of omega-3 fatty acids to total fatty acids and the ratio of omega-6 fatty acids to omega-3 fatty acids. According to the GWAS ID, we named the ratio of omega-3 fatty acids to total fatty acids and ratio of omega-6 fatty acids to omega-3 fatty acids abbreviated as omega-3-pct and omega-6 by omega-3, respectively. They are derived from a large-scale UK Biobank GWAS of fatty acids with 114,999 individuals of European ancestry (Borges et al., 2022). Circulating omega-3 (i.e., DHA and total omega-3) and omega-6 (i.e., linoleic acid and total omega-6) fatty acid concentration were measured using a targeted high-throughput nuclear magnetic resonance metabolomics platform (Julkunen et al., 2021). The mean concentration of total omega-3 fatty acids, DHA and omega-6 fatty acids was 0.53 (SD 0.22) mmol/L, 0.23 (SD 0.08) mmol/L and 4.45 (SD 0.68) mmol/L. (Supplementary Table 1)

For the outcome, the genetic effect of the corresponding SNP on IA and aSAH were obtained from the most recent GWAS with a total of 65796 Europeans (6252 IA cases and 59544 controls) and 63740 Europeans (4196 aSAH cases and 59544 controls) after excluding UK Biobank (Bakker et al., 2020). And Summary statistics for uIA phenotypes were extracted from the FinnGen database (https://r8.finngen.fi/pheno/I9_ANEURYSM) with a total of 314556 Europeans (1788 uIA cases and 312768 controls). (Supplementary Table 1)

According to three essential model assumptions of MR. First, SNPs were associated with the appropriate exposure at the genome-wide significance threshold P < 5×10-8. Second, to quantify the strength of the genetic instruments, SNPs with an F statistic less than 10 were excluded. Third, we clustered SNPs in linkage disequilibrium (LD, R2≥0.001 and within 10 mb). Fourth, remove SNPs that are obscure, palindromic, and linked to recognized confounding variables (body mass index (McDowell et al., 2018), blood pressure (Sun et al., 2022), type 2 diabetes (Tian et al., 2022), high-density lipoprotein (Huang et al., 2018)). Fifth, If an instrumental SNP for the exposure was not available from the outcome data set, we replaced it with a suitable proxy SNP (r^2^>0.8 in the European 1000 Genomes Project reference panel using LDlink https://ldlink.nci.nih.gov/]) or removed it in the absence of such a proxy. SNPS located in the MHC region were removed. Finally, removing SNPs with potential pleiotropy and outlier through mendelian randomization pleiotropy residual sum and outlier (MR-PRESSO). Then, the heterogeneity between-SNP were tested using inverse variance weighting (IVW) and the MRLJEgger method based on the SNPs that were retained after pleiotropy correction. (Figure 1.) It can be observed that the final SNPs for each group is not identical. Details of the SNPs used as instrumental variables were displayed in Supplementary Tables 2 - 4.

**Figure 1.**
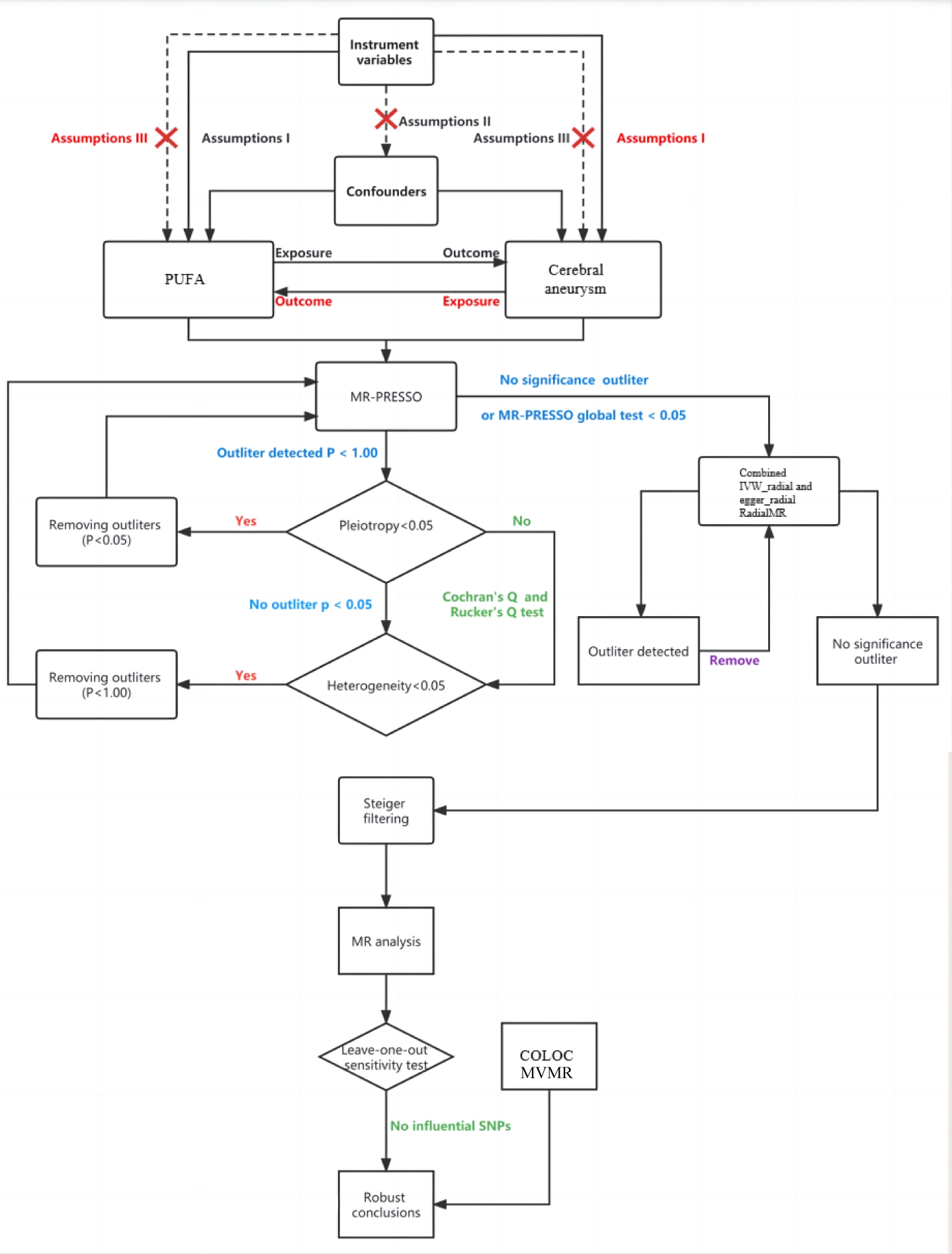
Flowchart showing Mendelian randomization. MR, Mendelian randomization; MR-PRESSO, Mendelian randomization pleiotropy residual sum and outlier; SNPs, single-nucleotide polymorphisms; MVMR, multivariable Mendelian randomization; IVW: inverse variance weighting; PUFA: polyunsaturated fatty acids; COLOC: colocalization.

### Mendelian randomization analyses

Eleven MR analytical approaches were used in this study to investigate the causal impacts of PUFA on the cerebral aneurysm. As the main analysis, we used standard IVW estimates, which are characterized by an analysis that does not take into account the presence of an intercept term and uses the inverse of the outcome variance (quadratic of the standard error) as a weight to provide a comprehensive estimate of the impact of the PUFA on the cerebral aneurysm. Furthermore, additional MR analyses, such as MR-Egger regression, weighted median, simple mode, weighted mode methods and so on were implemented as complements to the IVW, because these methods could provide more robust estimates over a wider range of scenarios. The MR-Egger method can provide causal estimates that are unaffected by breaches of standard IV assumptions and can detect whether standard IV assumptions are violated (Bowden et al., 2015). The weighted median method provides consistent estimates of effect, when at least half of the weighted variance provided by horizontal pleiotropy is valid (Bowden et al., 2016). We also performed reverse MR studies, where the cerebral aneurysms were treated as the exposure and PUFA was treated as the result, in order to thoroughly investigate the potential existence of reverse causation.

### Sensitivity analysis

For comparison with univariate MR results, to remove bias from closely related lipid traits, we performed multivariable Mendelian randomization-IVW (MVMR-IVW), adjusting for HDL cholesterol, LDL cholesterol, total cholesterol and triglycerides (Borges et al., 2022). To assess the robustness of the above results, a series of sensitivity analyses, including IVW, MR-Egger, MR-Egger intercept test, MR-PRESSO global test and “leave-one-out” were conducted. Furthermore, FADS2 and its SNPs within 500 kb of the FADS2 locus were removed based on the results of leave one out for the purpose of performing sensitivity analyses.

### Bayesian colocalization (COLOC)

Bayesian colocalization (COLOC) analysis assesses whether single-nucleotide variants linked to gene expression and phenotype at the same locus are shared causal variants, and thus, gene expression and phenotype are “colocalized.” This approach assumes that: (1) in each test region, there exists at most one causal SNP for either trait; (2) the probability that a SNP is causal is independent of the probability that any other SNP in the genome is causal; (3) all causal SNPs are genotyped or imputed and included in analysis. According to these assumptions, there are five mutually exclusive hypotheses for each test region: (1) there is no causal SNP for either trait (H0); (2) there is one causal SNP for trait 1 only (H1); (3) there is one causal SNP for trait 2 only (H2); (4) there are two distinct causal SNPs, one for each trait (H3); and (5) there is a causal SNP common to both traits (H4). A large PP for H4 (PP.H4 above 0.75) strongly supports shared causal variants affecting both gene expression and phenotype (Kurki et al., 2023).

### Statistical Analysis

For the MR analysis, IVs with F values over 10 were considered strong instruments that could alleviate bias from weak instruments (Pierce et al., 2011). The formula used to calculate F values is as follows: F = [(N-K-1) R2]/[k(1-R2)], where R2 represents the proportion of variance explained by the genetic variants, N represents the sample size, and k represents the number of included SNPs. R2 = ∑^k^ 2β^2^(1-EAF)EAF, where EAF is the effect allele frequency and β is the estimated effect on LTL (Park et al., 2010). Given the limited number of cases in the GWAS, we calculated the statistical power for MR analysis using the mRnd website (https://shiny.cnsgenomics.com/mRnd/). To account for multiple testing in our primary analyses, a Bonferroni-corrected threshold of P < 0.0033 (α=0.05/15 [5 exposures and 3 outcomes]) was applied. All statistical analyses performed in this investigation were carried out using the “TwoSampleMR”, “phenoscanner”, “RadialMR”, “MendelianRandomization” and “MRPRESSO” packages in R software (version 4.2.2).

## Results

According to the above methodology, we filtered out the corresponding SNPs. (Supplementary Table 2 – 4)

In the outcome of IA, IVW analysis indicated that omega-3 fatty acid (OR = 0.80, 95% CI: 0.69 - 0.91, P = 1.01ⅹ10^-3^), DHA (OR = 0.75, 95% CI: 0.63 - 0.87, P = 3.12ⅹ10^-4^) and omega-3-pct (OR = 0.8, 95% CI: 0.71 - 0.90, P = 2.34ⅹ10^-4^) decreased the risk for IA. And omega-6 by omega-3 increased the risk for IA (OR = 1.27, 95% CI: 1.12 – 1.44, P = 1.53ⅹ10^-4^). The results demonstrated consistency in the outcomes and directionality of the various MR methods employed. (Supplementary Figure 1) Nevertheless, there was no link found between omega-6 fatty acid and IA. (Figure 2) The results of the MVMR demonstrated that omega-3, DHA, and omega-3-pct were also effective in reducing the incidence of IA following appropriate adjustments for HDL cholesterol, LDL cholesterol, total cholesterol and triglycerides. Furthermore, an elevated ratio of omega-6 by omega-3 was associated with an increased risk of developing IA. (Figure 5)

**Figure 2.**
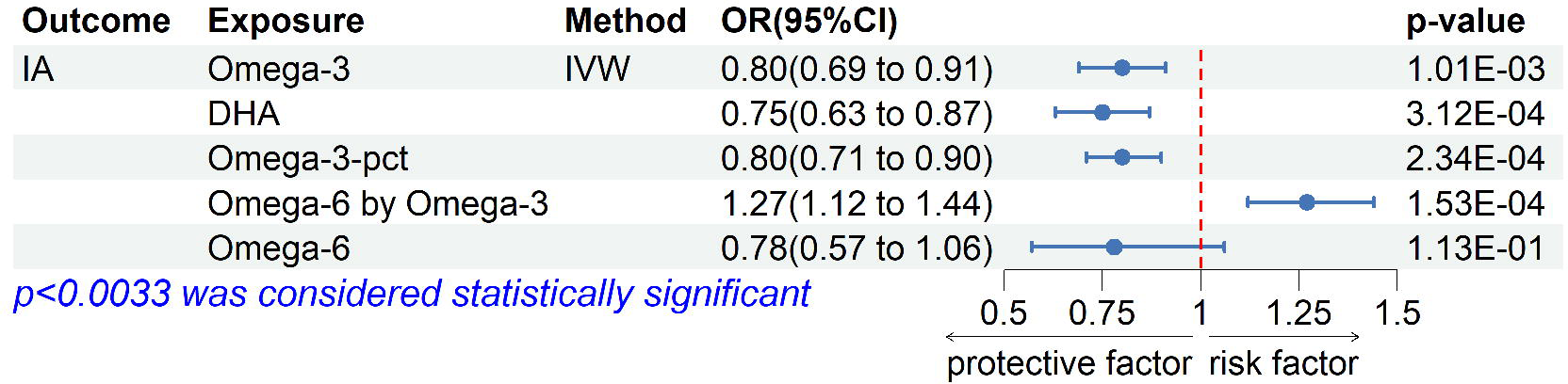
Mendelian randomization analysis of the association between PUFA and IA. PUFA: polyunsaturated fatty acids; IA: intracranial aneurysm;OR: odds ratio; CI: confidence interval; MR, Mendelian randomization; IVW: inverse variance weighting; DHA: docosahexaenoic acid.

In the outcome of aSAH, omega-3 fatty acid (OR = 0.71, 95% CI: 0.61 - 0.84, P = 3.73ⅹ10^-5^), DHA (OR = 0.67, 95% CI: 0.55 - 0.80, P = 2.32ⅹ10^-5^) and omega-3-pct (OR = 0.76, 95% CI: 0.66 - 0.87, P = 1.16ⅹ10^-4^) decreased the risk for aSAH. And omega-6 by omega-3 increased the risk for aSAH (OR = 1.35, 95% CI: 1.17 – 1.56, P = 5.78ⅹ10^-5^). The results and directionality of other MR methods revealed consistency. (Supplementary Figure 2) However, there was no causal relationship between omega-6 fatty acid and aSAH. (Figure 3.) After adjustment for HDL cholesterol, LDL cholesterol, total cholesterol and triglycerides, the results of the MVMR demonstrated that omega-3, DHA and omega-3-pct were also effective in reducing the incidence of IA. (Figure 6)

**Figure 3.**
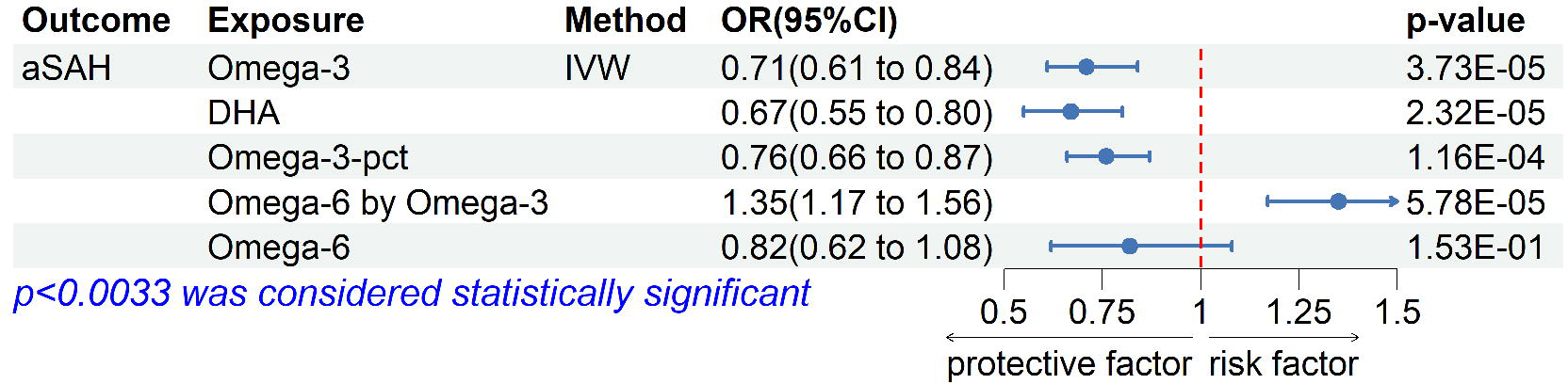
Mendelian randomization analysis of the association between PUFA and aSAH. PUFA: polyunsaturated fatty acids; aSAH: aneurysmal subarachnoid hemorrhage; OR: odds ratio; CI: confidence interval; MR, Mendelian randomization; IVW, inverse variance weighting; DHA: docosahexaenoic acid.

**Figure 4.**
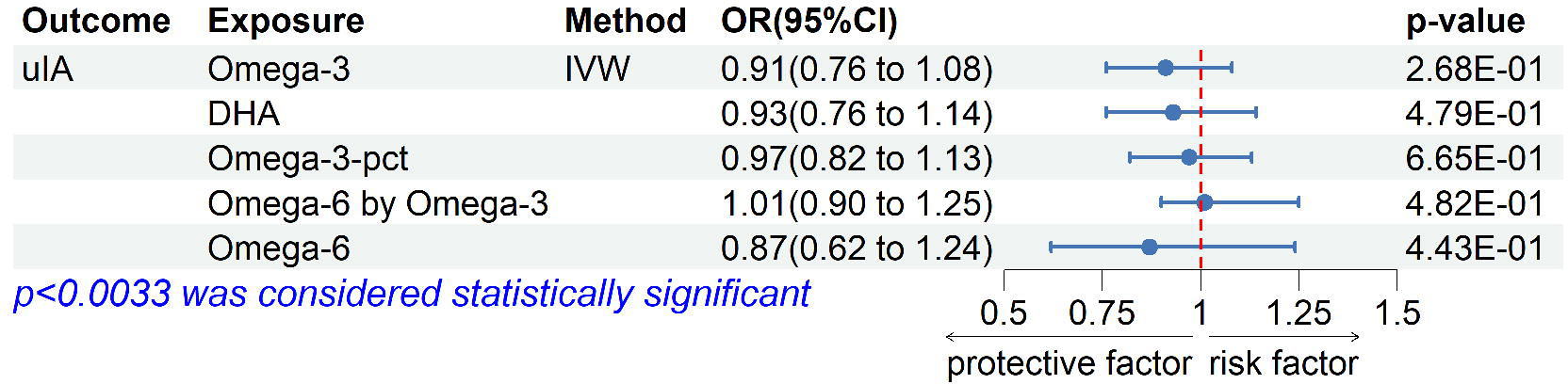
Mendelian randomization analysis of the association between PUFA and uIA. PUFA: polyunsaturated fatty acids; uIA: unruptured intracranial aneurysm; OR: odds ratio; CI: confidence interval; MR, Mendelian randomization; IVW, inverse variance weighting; DHA: docosahexaenoic acid.

**Figure 5.**
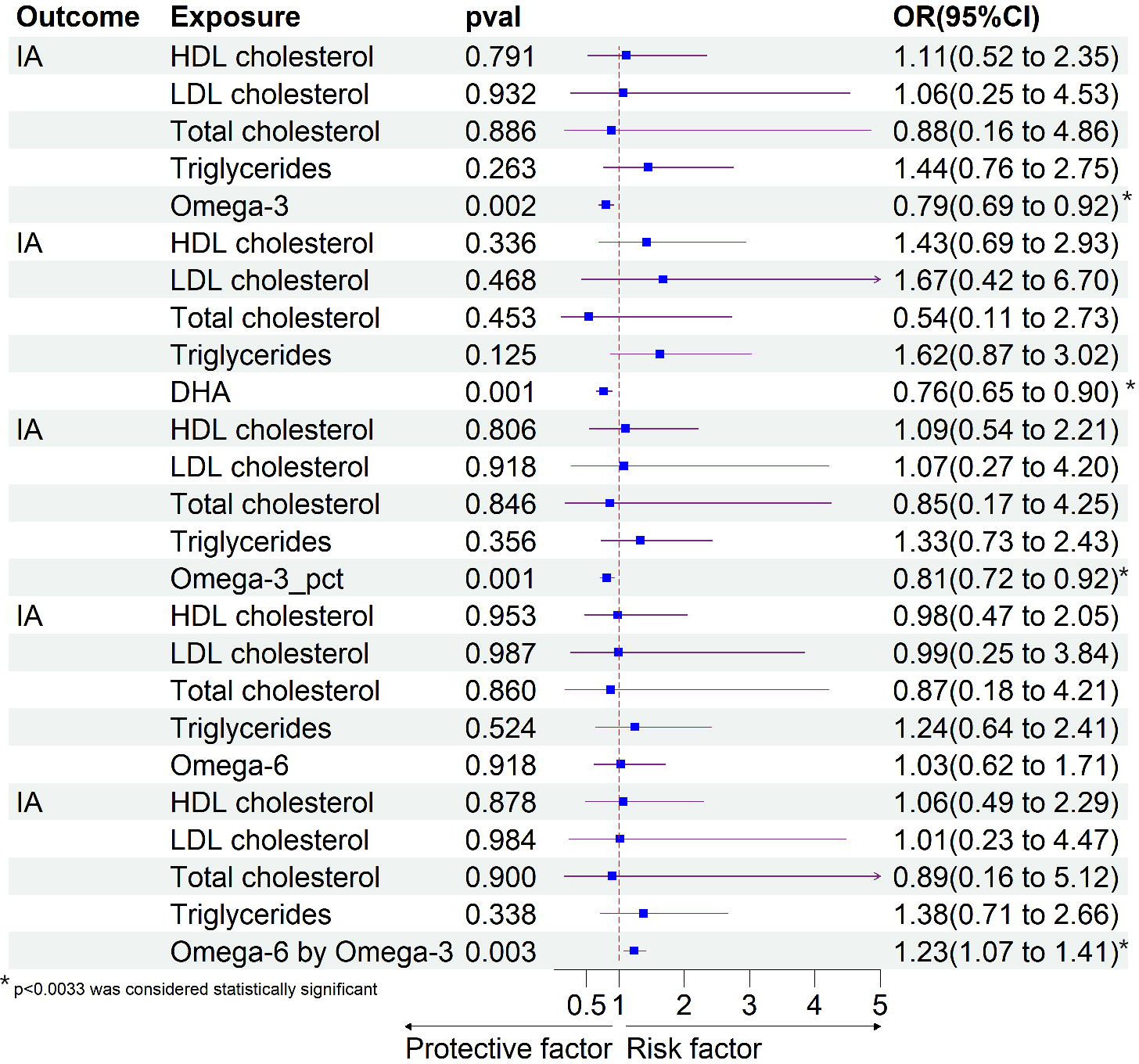
Multivariable Mendelian randomization analysis of the association between PUFA and IA. IA: intracranial aneurysm; DHA: docosahexaenoic acid; LDL: low density lipoprotein; HDL: high density lipoprotein.

**Figure 6.**
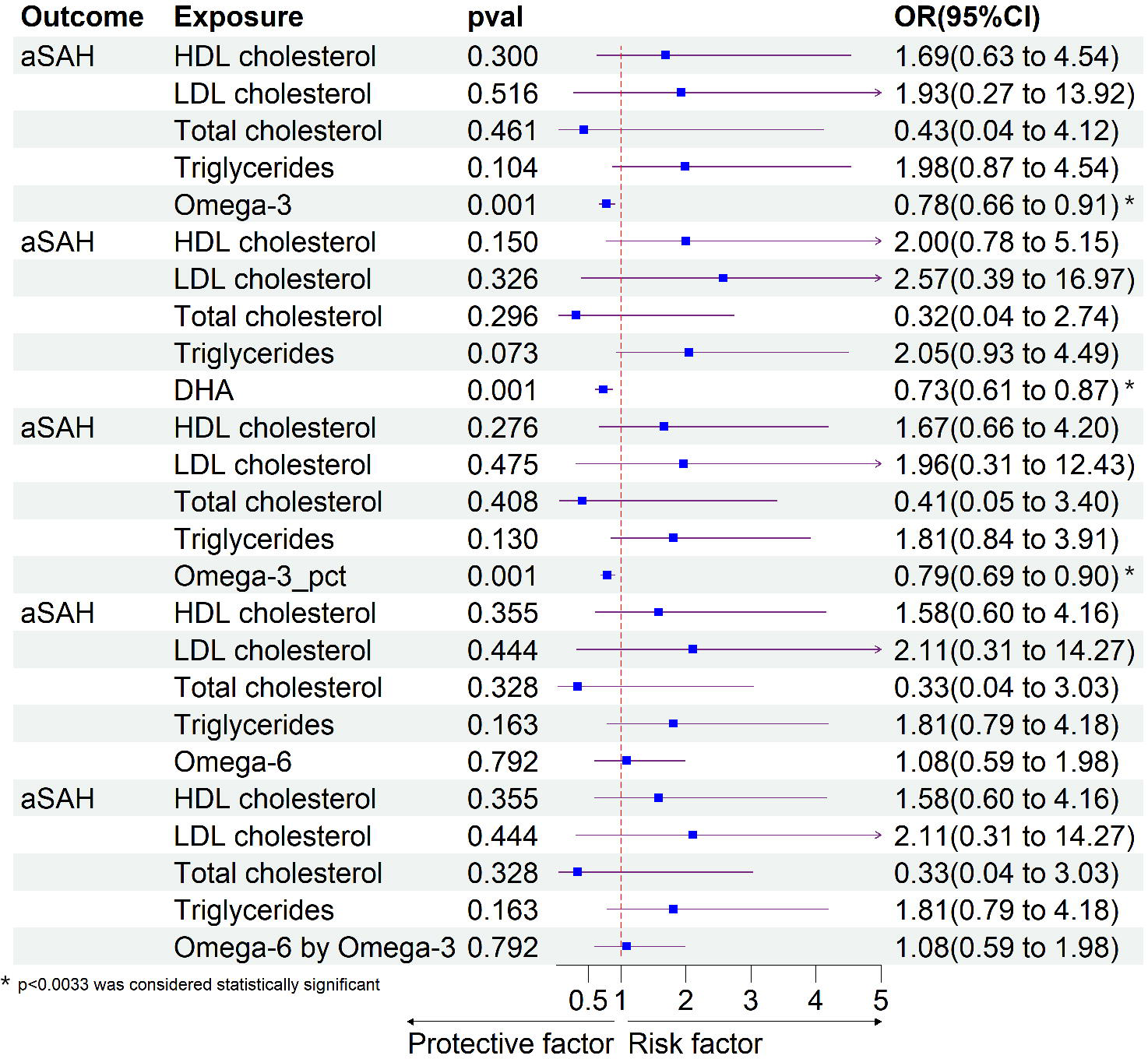
Multivariable Mendelian randomization analysis of the association between PUFA and aSAH. aSAH: aneurysmal subarachnoid hemorrhage; DHA: docosahexaenoic acid; LDL: low density lipoprotein; HDL: high density lipoprotein.

In the outcome of uIA, there was no causal relationship between all of the above five PUFAs and uIA. (Figure 4.) In the result of reverse MR, MR analysis of all groups failed sterling the filiting test. Adjusted MVMR still shows negative results. (Figure 7)

**Figure 7.**
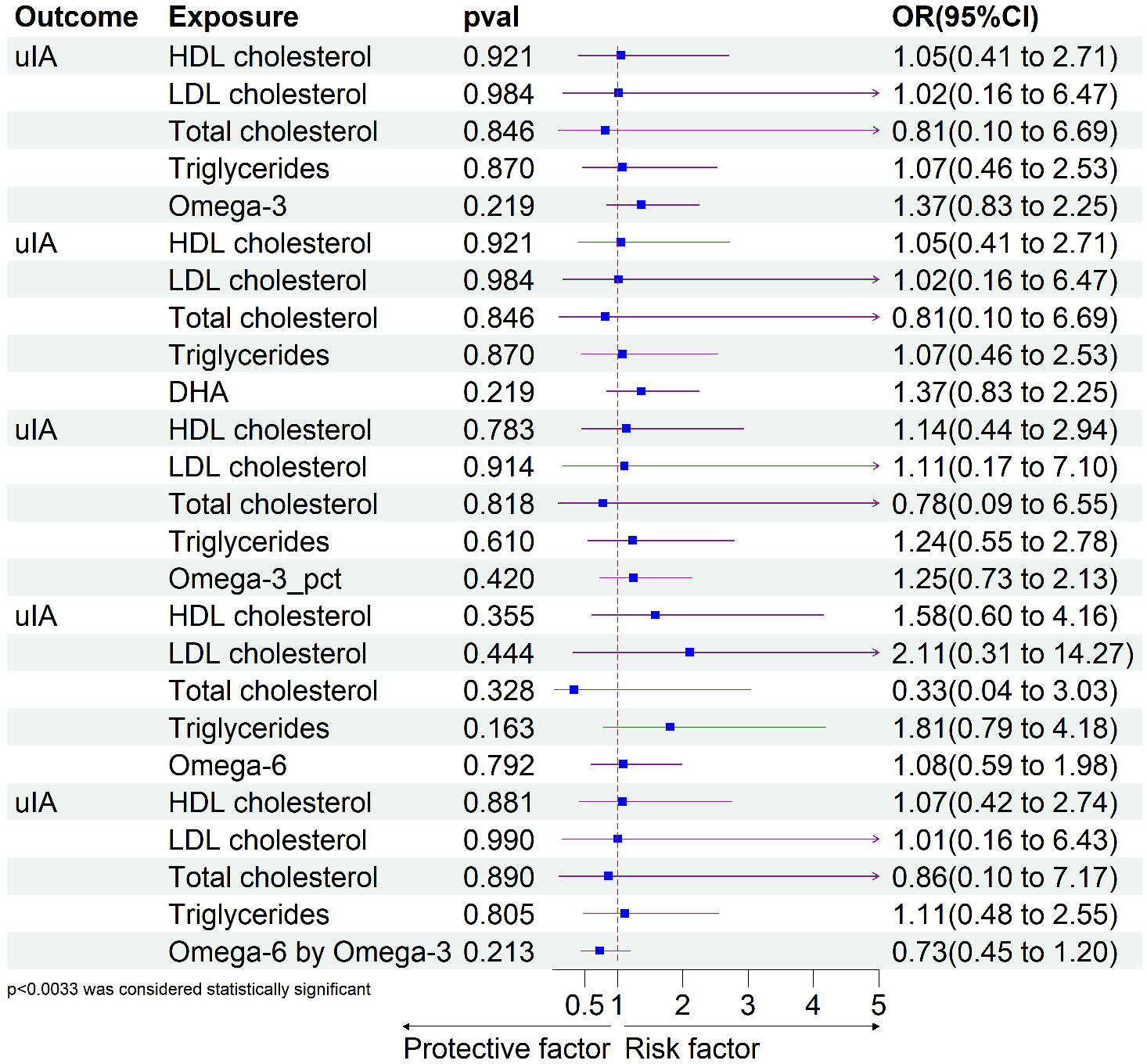
Multivariable Mendelian randomization analysis of the association between PUFA and uIA. uIA: unruptured intracranial aneurysm; DHA: docosahexaenoic acid; LDL: low density lipoprotein; HDL: high density lipoprotein.

To assess the robustness of the above results, a series of sensitivity analyses, including MR-Egger, IVW, MR-Egger intercept test, “leave-one-out” method and MR-PRESSO global test, were conducted (Supplementary Table 5 and Supplementary Figure 3-5). All P-values of the MR-Egger intercept tests, Q test and MR-PRESSO were > 0.05, indicating that no horizontal pleiotropy and heterogeneity existed. All steiger tests indicated correct directionality. The leave-one-out plot demonstrates that there is a potentially influential SNP (rs174564) driving the causal link between PUFA and cerebral aneurysm. We conducted further sensitivity analyses after removing rs174564 and its SNPs within 500 kb of the FADS2 locus. The results demonstrated that the p-values were all greater than 0.0033, indicating that SNP: rs174564 was a significant factor driving the causal association between PUFAs and CA. (Figure 8)

**Figure 8.**
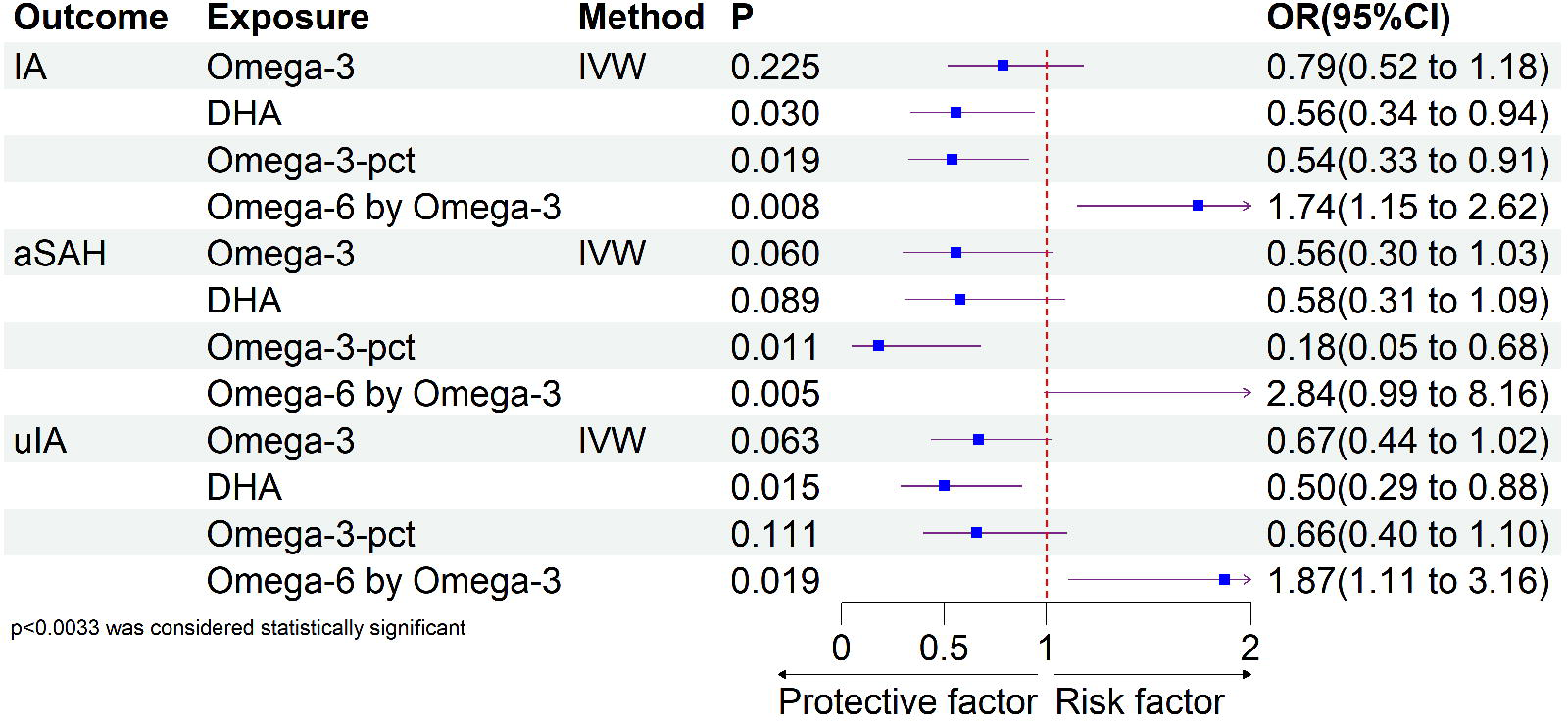
The sensitivity analysis of the association between PUFA and CA after remove FADS2 locus. IA: intracranial aneurysm; aSAH: aneurysmal subarachnoid hemorrhage; uIA: unruptured intracranial aneurysm; DHA: docosahexaenoic acid; IVW: inverse variance weighting

There is consistent evidence that PUFAs have a beneficial causal effect on cerebral aneurysm. In order to determine an MR estimate is not confounded by LD, we used COLOC to identify shared causal SNP between PUFAs and cerebral aneurysms. In COLOC analysis, we further calculated the posterior probability of each SNP being causal to both of the traits (PPH4). We identified shared causal SNP of omega-3 fatty acid and IA, aSAH in gene fatty acid desaturase 2 (FADS2), respectively. Similarly, in DHA, omega-3-pct and omega-6 by omega-3, these three fatty acids and IA, aSAH also obtain the same result. (Table 1 and Supplementary Table 6_1-6_8)

## Discussion

PUFA supplementation is controversial for the prevention of cerebral aneurysm disease, while cerebral aneurysm is a heterogeneous disease with various pathogenic mechanisms. Here, we implemented multiple rigorous MR methods to appraise the possible causal association of PUFA with cerebral aneurysm and its different states. We demonstrated that the omega-3 fatty acids, DHA and omega-3-pct causally decreased the risk for IA and aSAH. And omega-6 by omega-3 causally increased the risk of IA and aSAH. In addition to IVW, consistency was seen in the outcomes and directionality of different MR methods. The results of the MVMR and sensitivity analysis provide further evidence of the causal association between PUFAs and IA, aSAH. We identified shared causal SNPs of omega-3 fatty acid, DHA, omega-3-pct, omega-6 by omega-3 and IA in genes *FADS2*, respectively. The same results were acquired from these four PUFAs and aSAH. The results of reverse MR and COLOC jointly increase the validity of our findings.

This could be connected to PUFAs, which can decrease the aneurysm’s inflammatory process. The pathophysiology of IA involves inflammatory processes (Ge et al., 2022), which may include degeneration of the endothelium and vasa vasorum, as well as fragmentation of the internal elastic lamina (Nakatomi et al., 2000; (Meng et al., 2014). As an underlying mechanism, the prostaglandin E2 in macrophages was considered by Aoki et al. as a factor regulating such chronic inflammation involved in the progression and rupture of IAs (Aoki et al., 2017). However, multiple elements of the inflammatory process, including oxidative stress and inflammation in macrophages, can be downregulated by omega-3 and DHA (Meital et al., 2019). In the abdominal aortic aneurysm (AAA), supplementation with the omega-3 PUFA, DHA, suppressed production of 8-isoprostane, the gold standard biomarker of oxidative stress and a specific marker of lipid peroxidation (Kadiiska et al., 2005). The results concord with data from both experimental animal studies (Saraswathi et al., 2007; (Zúñiga et al., 2010) and human clinical trials (Mori et al., 2003; (Barden et al., 2015) that suggest omega-3 PUFAs decrease 8-isoprostane levels and ease oxidative stress. The findings from a randomized controlled trial suggest that omega-3 PUFAs improve erythrocyte fatty acid profile and ameliorate factors associated with inflammation in AAA patients (Meital et al., 2019). For aSAH, a prospective study demonstrated that administration of omega-3 fatty acids after aneurysmal SAH may reduce the frequency of cerebral vasospasm through the effects of anti-inflammatory and reduce both platelet aggregation and plasma triglycerides, and have neuroprotective effects and may improve clinical outcomes (Nakagawa et al., 2017). At the same time, it may have a preventive effect on the complication of intracranial hemorrhage after aneurysm surgery (Saito et al., 2017). A multi-center study with a sizable sample size is still required to clarify the trend of the effect of omega-3 fatty acids on aSAH, even though the favorable outcomes of these two studies have a certain reference value. Furthermore, Ririko et al. findings that AA and AA-containing phospholipids were not detected in the unruptured IA while they were detected in a significantly larger amount in the ruptured IA and suggest that the stability of the turnover of AA and linoleic acid in human unruptured IA walls is sustained (Takeda et al., 2021). Several earlier studies employing rat model found that the thickened aneurysmal wall and various brain areas after subarachnoid hemorrhage had higher amounts of free fatty acids, including AA (Gewirtz et al., 1999; (Ikedo et al., 2018). This is in line with results of our MR analyses between omega-6 fatty acid and aSAH.

Besides, in the development of cardiovascular disease, genetically higher plasma α-linolenic and linoleic levels are inversely associated with large-artery stroke and venous thromboembolism, whereas AA level have a favorable correlation with these cardiovascular diseases (Yuan et al., 2019). This may be associated with the Δ5-desaturase and Δ6-desaturase, which are the primary rate-limiting enzyme for the synthesis of inborn long-chain PUFAs and is encoded by the fatty acid desaturase 1 (*FADS1*) gene and fatty acid desaturase 2 (*FADS2*) gene (Simopoulos, 2010). Even the aortic valve’s fatty acid composition might be impacted by *FADS1* (Plunde et al., 2020). In our COLOC analysis, we identified shared causal SNP of omega-3 fatty acid and IA, aSAH in gene fatty acid desaturase 2 (*FADS2*), respectively. The same result was also found in DHA, omega-3-pct and omega-6 by omega-3. Furthermore, sensitivity analyses of removing the FADS2 locus yielded analogous results, thereby corroborating the result that FADS2 drives the causal relationship. It is obvious that omega-3 fatty acids play a vital role in vascular-related diseases. Conversely, it’s incorrect to assume that all forms of omega-6 fatty acids would worsen blood vessel damage. More thorough large-sample study is still required for additional validation.

However, several limitations should be taken into account in our study. First, given that the participants in this study were form European population, the results of this study cannot be smoothly extrapolated to other ethnic groups with diverse lifestyles and cultural backgrounds. Hence, further multi-ancestry studies are needed to evaluate whether our findings can be generalized to individuals of other ancestries. Second, since MR analyses extrapolated causal hypotheses by exploiting the random allocation of genetic variants, it was difficult to completely distinguish between mediation and pleiotropy using MR approaches. The generous variants in our genome probably affect one or more phenotypes, even though we made an effort to remove the impacts of pleiotropic and heterogeneous factors and confounders, some effects may still be there. Third, not all SNPs were examined, as some were removed because of LD (and no proxy SNPs were found), which may have impacted the results. Fourth PUFAs were measured from nonfasting peripheral plasma specimens, not aneurysm cavity blood and aneurysm wall tissue Fifth, there is currently a lack of GWAS datasets of cerebral aneurysms from different cerebrovascular sites, and exploring the relationship between genetically predicted PUFAs and the occurrence of specific types of cerebral aneurysms remains a challenge in current MR analyses.

## Conclusion

In view of the extremely poor prognosis of cerebral aneurysms and the high mortality rate of secondary rupture, our research focuses on the causal relationship between PUFAs and cerebral aneurysms to improve our understanding of the pathogenic mechanism of various PUFAs in cerebral aneurysms. And provide new insights into the diagnosis and treatment of brain aneurysms in the future.

## Data Availability

The GWAS databases we use are all from publicly published data, each of the original GWASs had obtained ethical approval and participant consent

## Acknowledgements

The authors want to acknowledge all of the participants and investigators for contributing and sharing summary-level data on GWAS.

## Consent for Publication

All participants provided informed consent before enrollment in our study.

## Author contributions

Study design: Weijie Yu, Yunyun Mei, Liwei Zhou and Zhanxiang Wang. Acquisition of datasets: Weijie Yu, Yunyun Mei, Chongfei Li, Xiaoyan Chen, Zhenwei Lu, Qionghui Huang and Zhangyu Li. Data analysis: Weijie Yu, Yunyun Mei and Chongfei Li. Manuscript drafting: Weijie Yu and Yunyun Mei. Manuscript revision: Liwei Zhou and Zhanxiang Wang. Final approval of manuscript: all authors.

## Ethical Approval

Due to the fact that each of the original GWASs had obtained ethical approval and participant consent, they were not required.

## Data availability

The original contributions presented in the study are included in the article/supplementary material. Further inquiries can be directed to the corresponding author.

