## Supplementary material for "Predicting the causal relationship between polyunsaturated fatty acids and cerebral aneurysm risk from a Mendelian randomization study": Supplymentary figure

**Supply figures**

Supplementary figure 1. The detailed results of MR between PUFA and IA

Supplementary figure 2. The detailed results of MR between PUFA and aSAH

Supplementary figure 3. The detailed results of causal direction and sensitivity analysis between PUFA and IA.

Supplementary figure 4. The detailed results of causal direction and sensitivity analysis between PUFA and aSAH.

Supplementary figure 5. The detailed results of causal direction and sensitivity analysis between PUFA and uIA.

Supplementary figure 1-1. The detailed results of MR between omega-3 [fatty](javascript:;) [acid](javascript:;) and IA

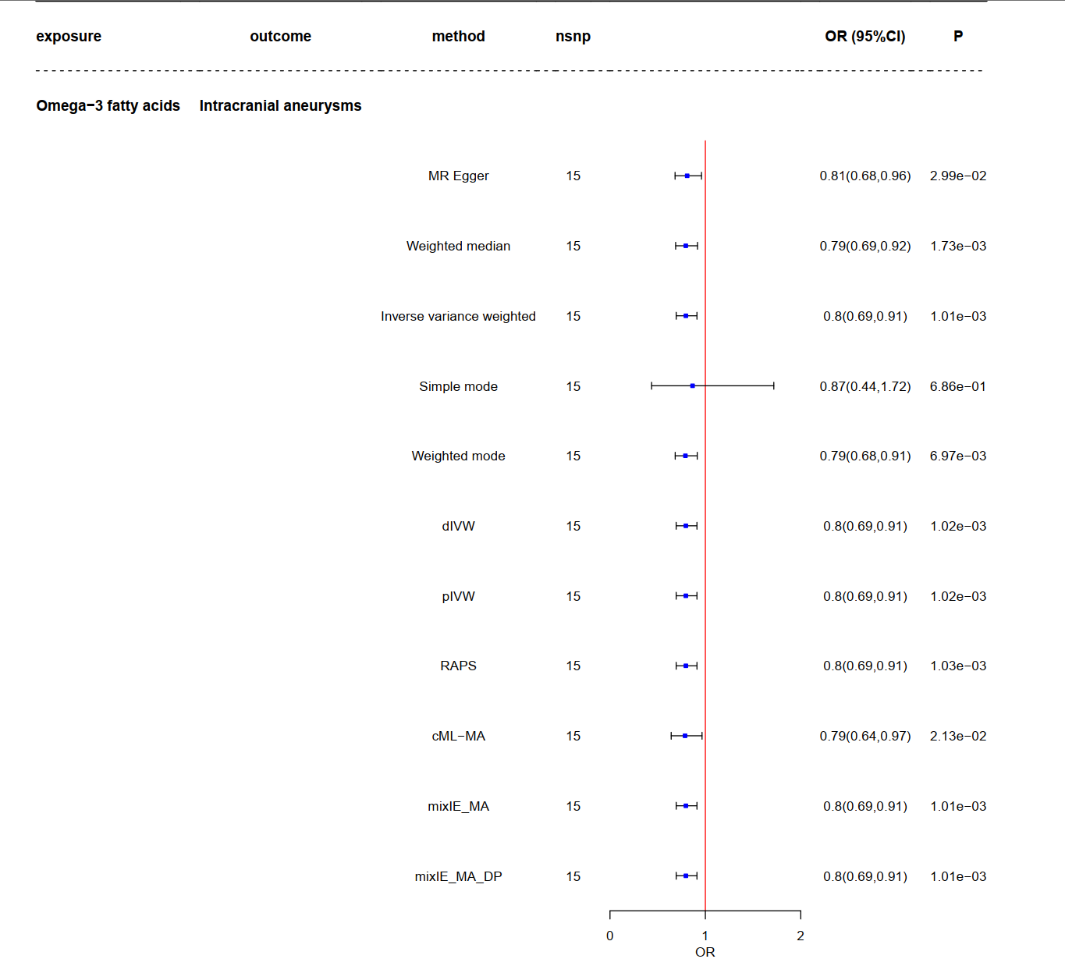

Supplementary figure 1-2. The detailed results of MR between DHA [acid](javascript:;) and IA

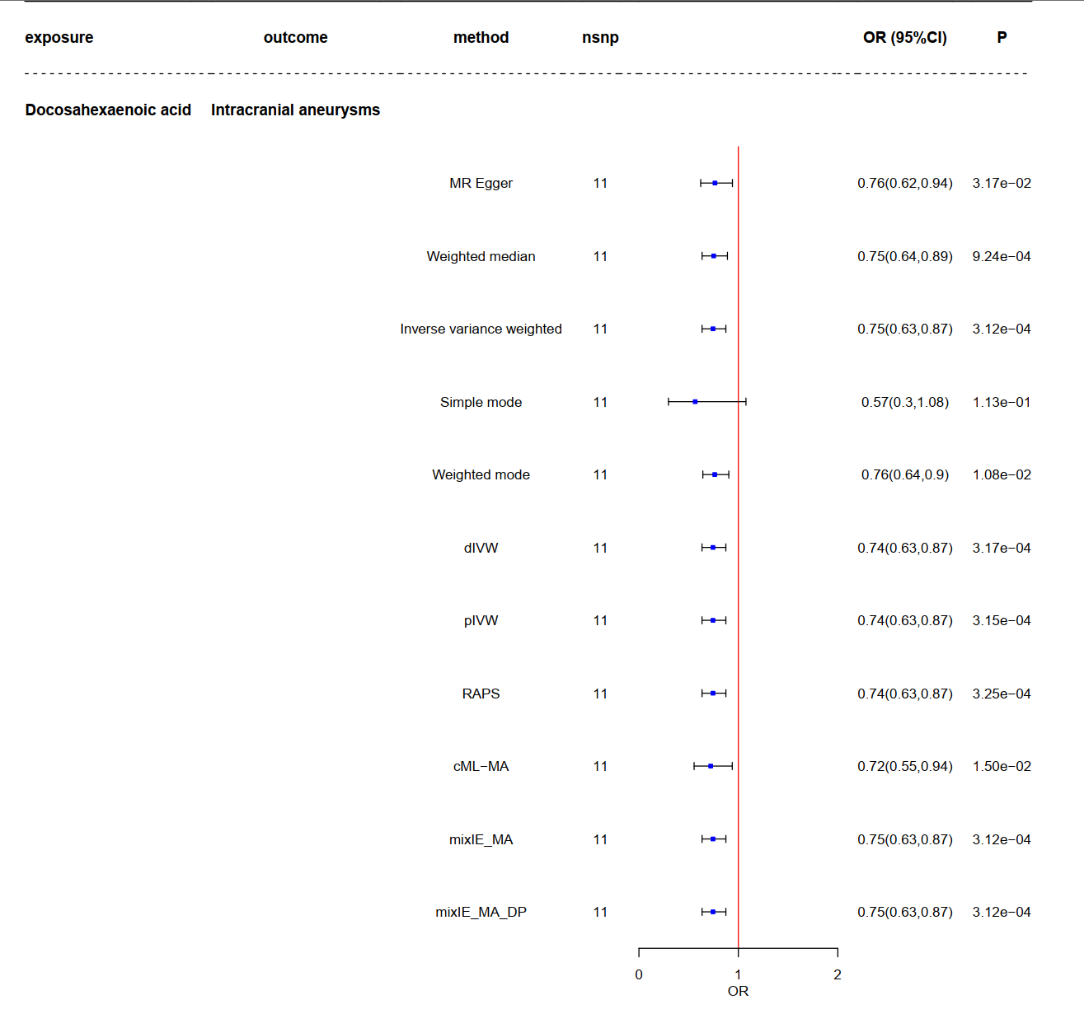

Supplementary figure 1-3. The detailed results of MR between omega-3-pct and IA

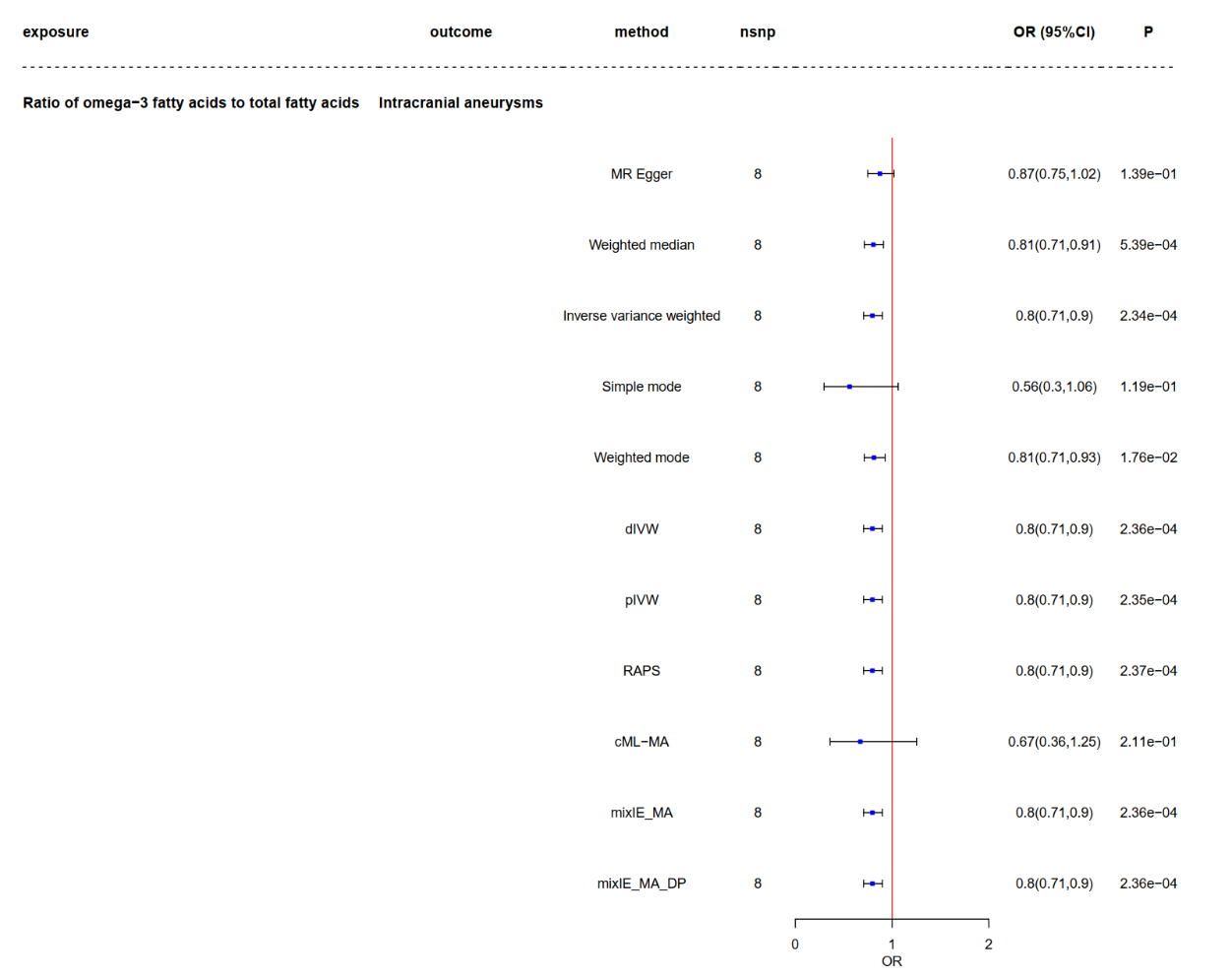

Supplementary figure 1-4. The detailed results of MR between omega-6 by omega-3 and IA

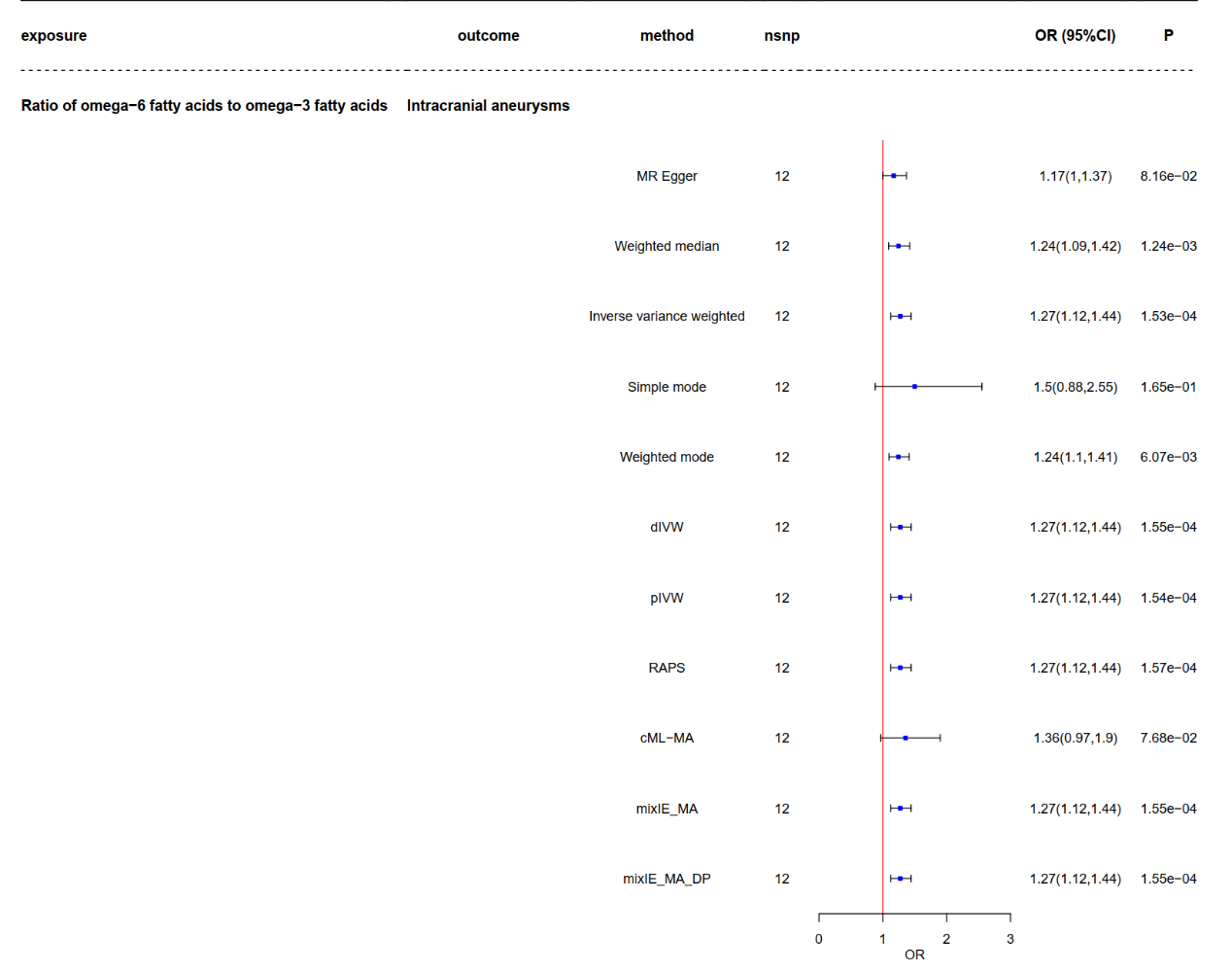

Supplementary figure 1-5. The detailed results of MR between omega-6 and IA

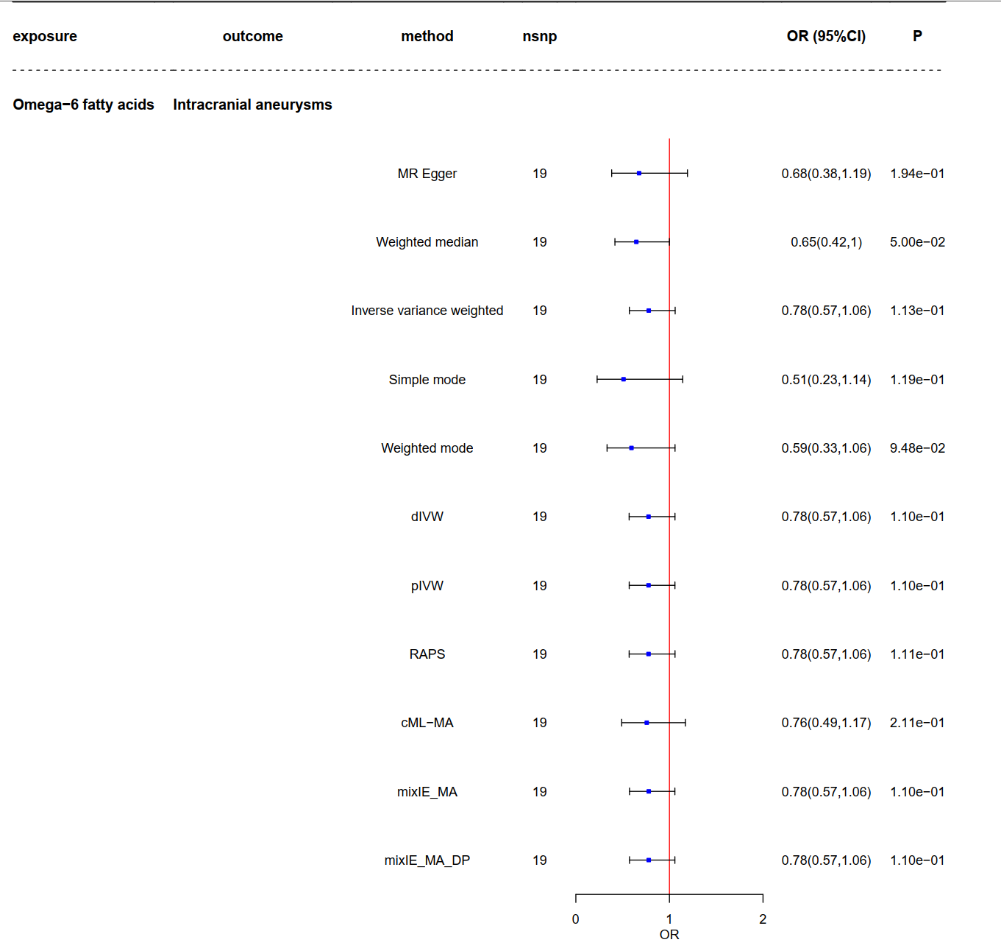

Supplementary figure 2-1. The detailed results of MR between omega-3 and aSAH

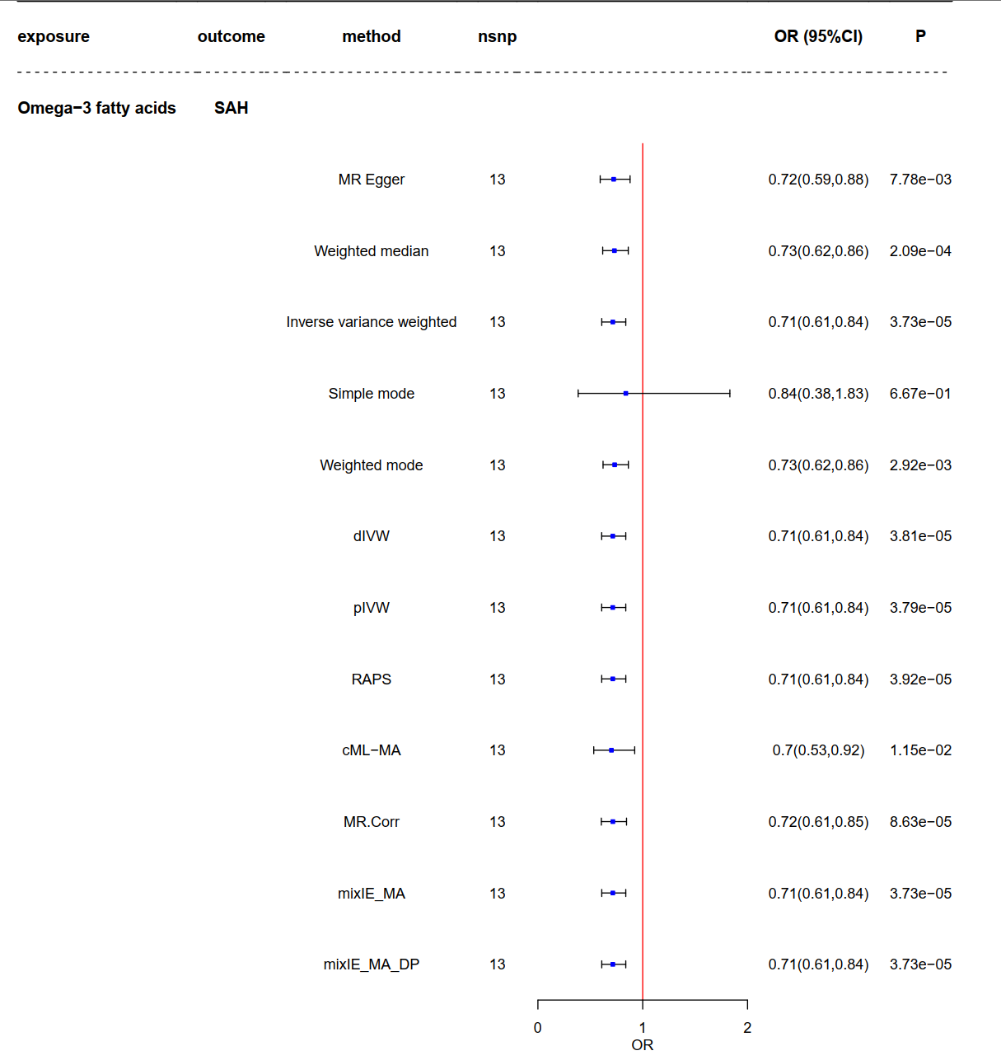

Supplementary figure 2-2. The detailed results of MR between DHA and aSAH

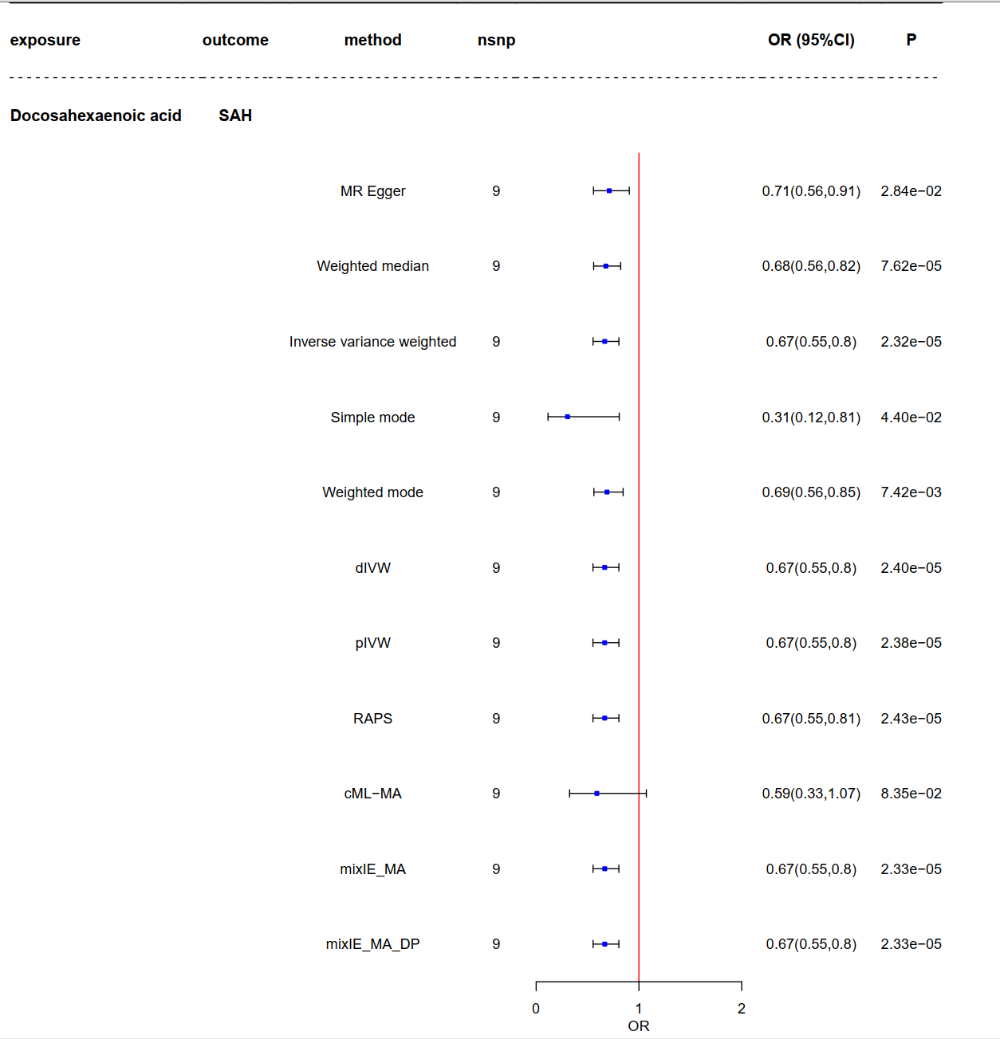

Supplementary figure 2-3. The detailed results of MR between omega-3-pct and aSAH

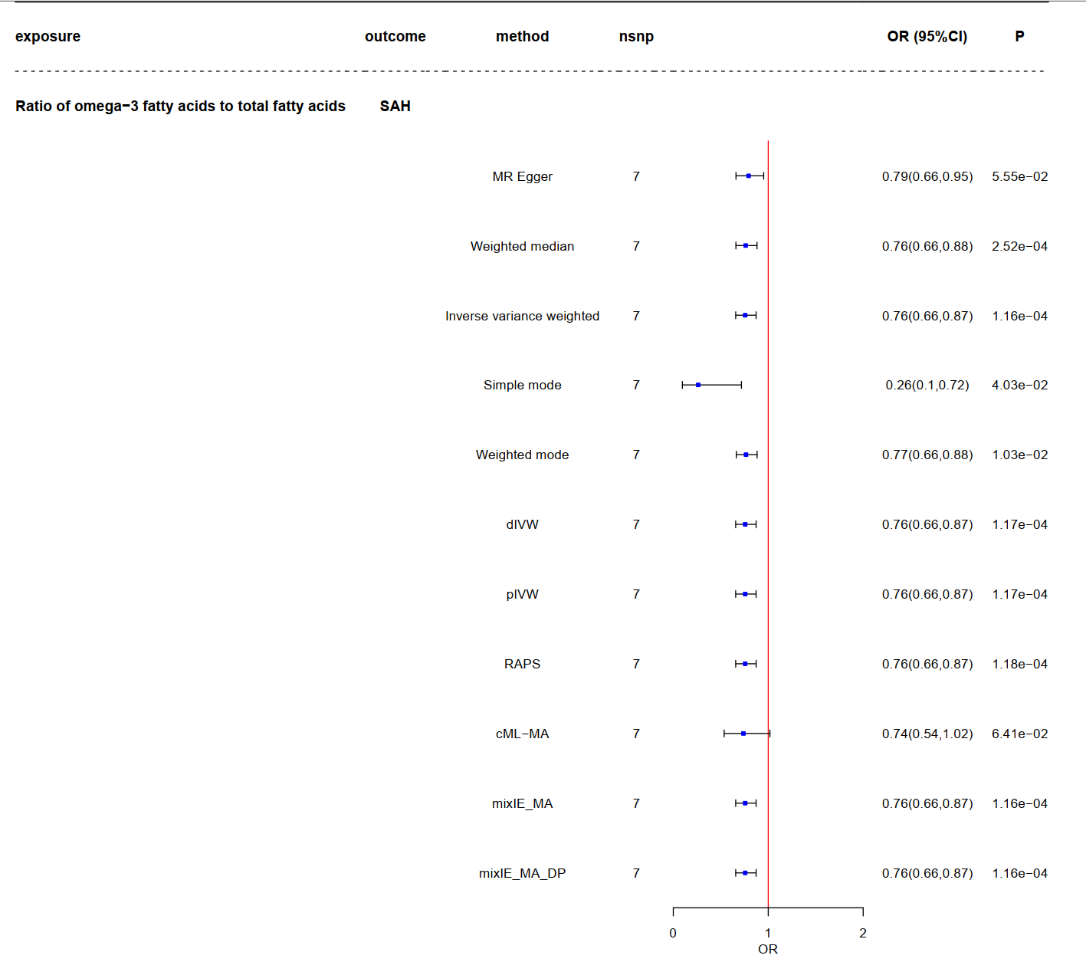

Supplementary figure 2-4. The detailed results of MR between omega-6 by omega-3 and aSAH

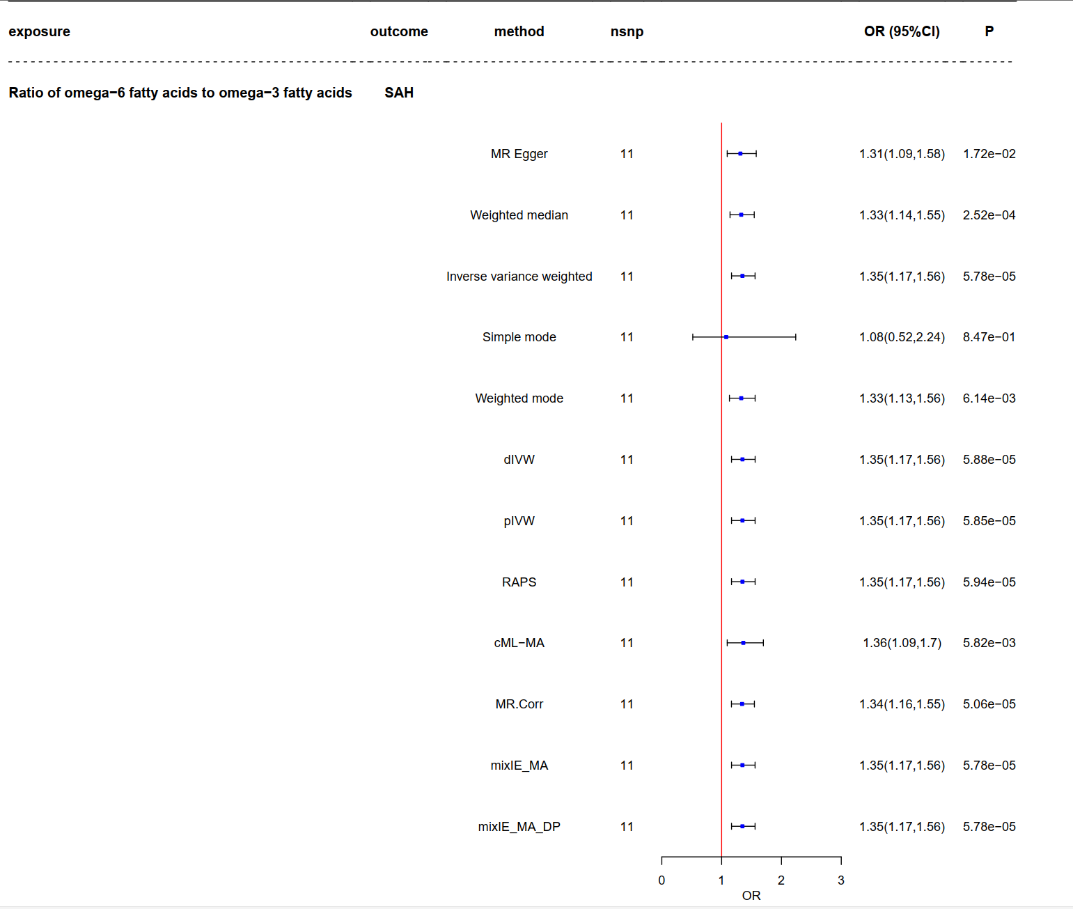

Supplementary figure 2-5. The detailed results of MR between omega-6 and aSAH

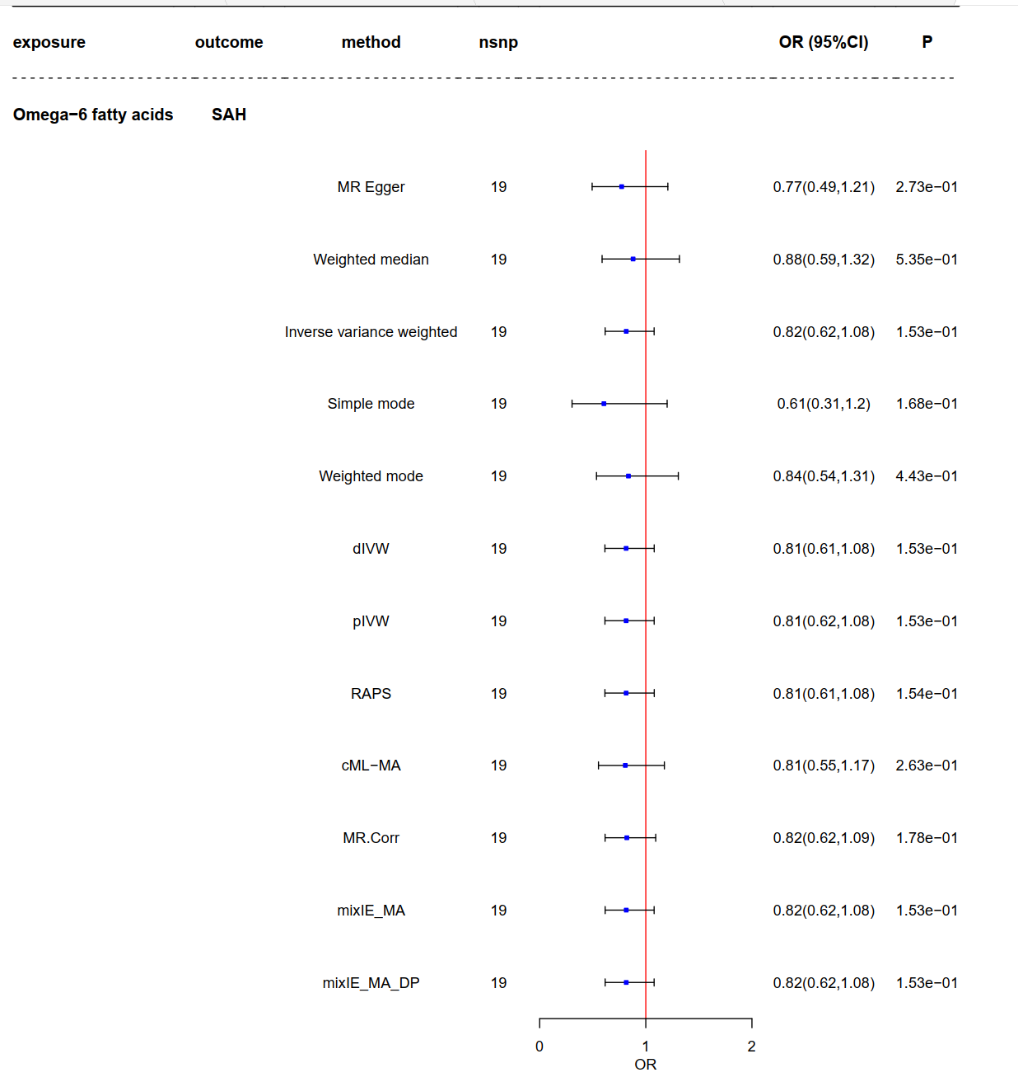

Supplementary figure 3-1. The detailed results of causal direction and sensitivity analysis between omega-3 and IA. A: scatter plot; B: funnel plot; C: forest plot; D: leave-one-out analysis.

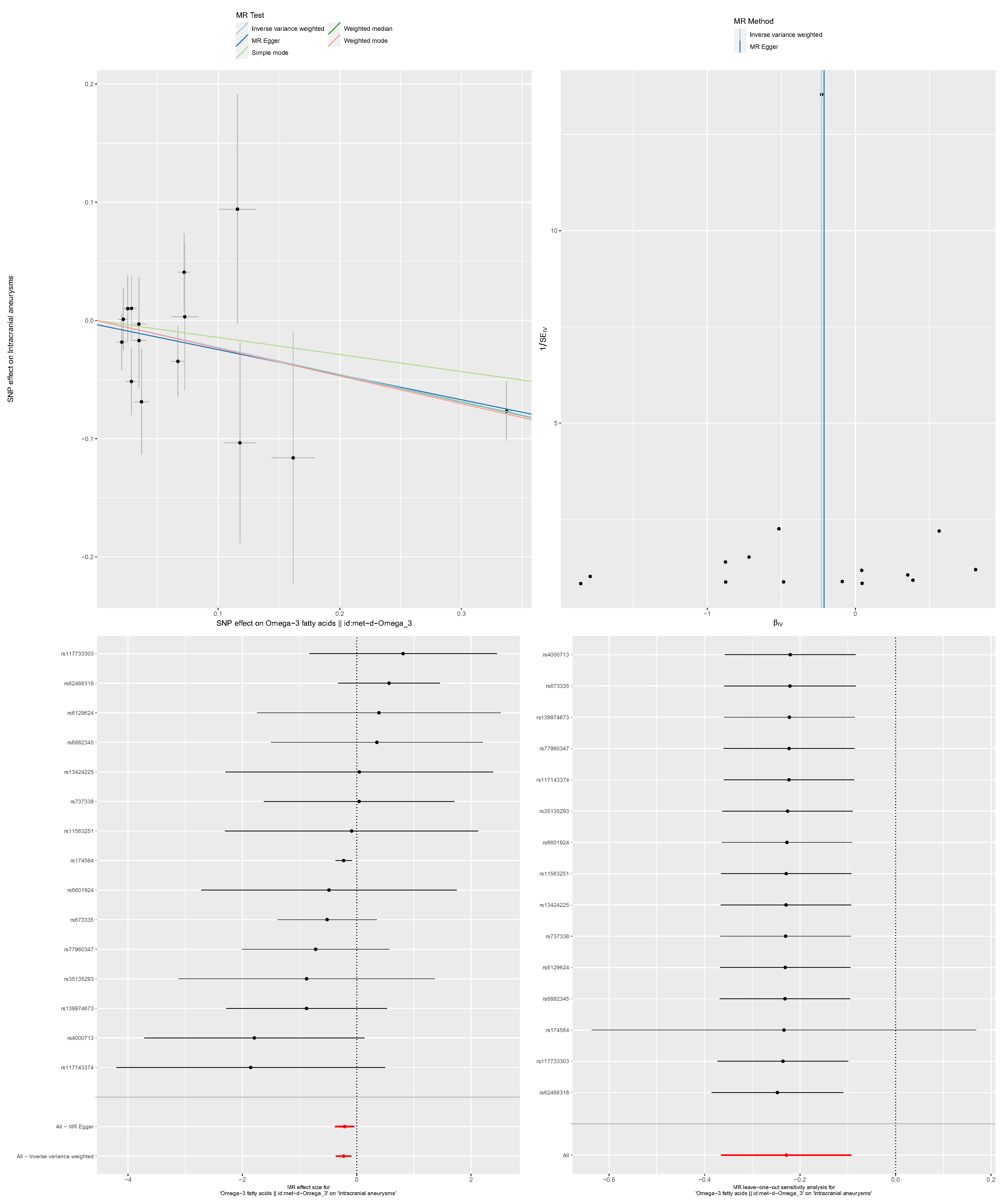

Supplementary figure 3-2. The detailed results of causal direction and sensitivity analysis between DHA and IA. A: scatter plot; B: funnel plot; C: forest plot; D: leave-one-out analysis.

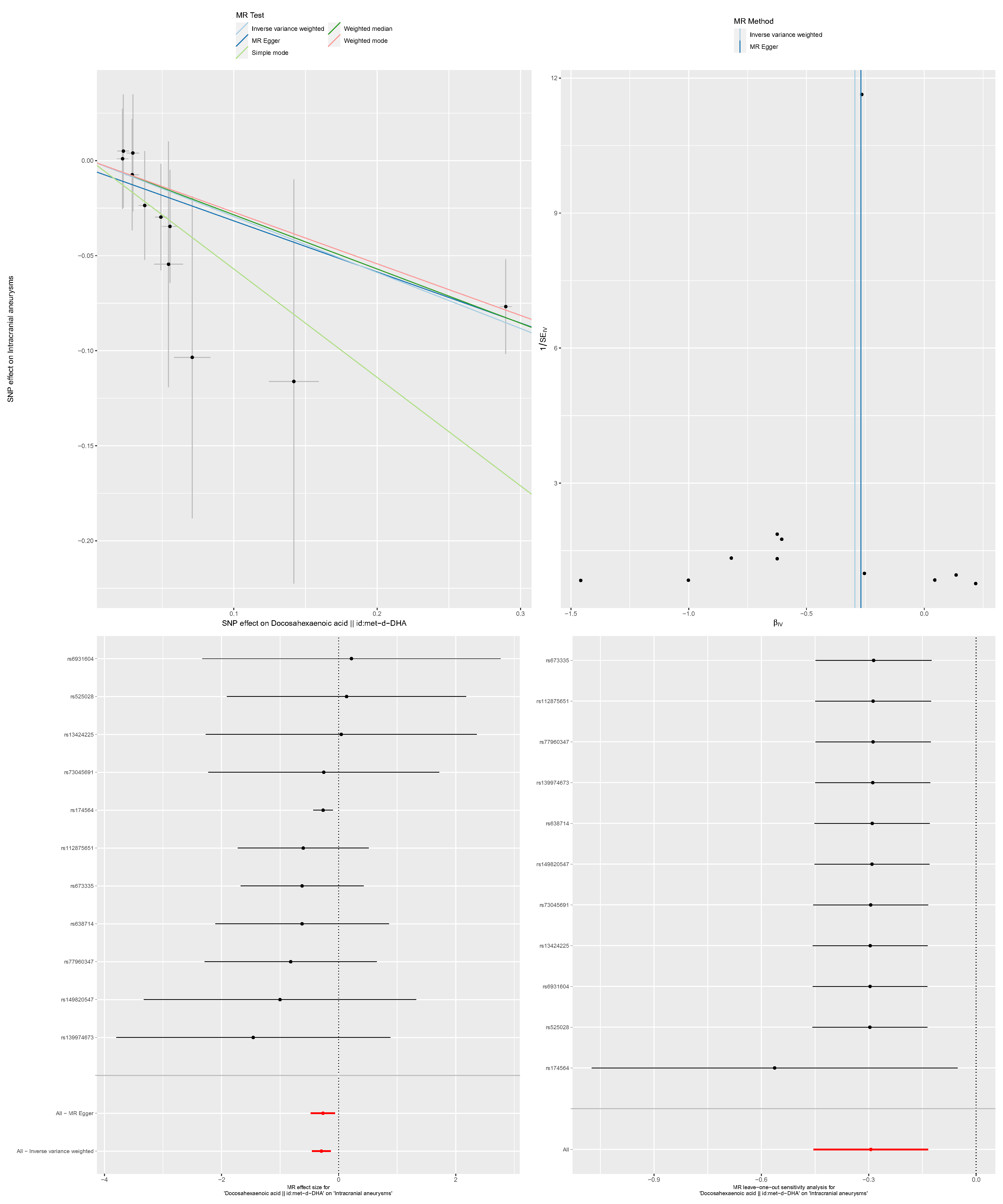

Supply figure 3-3. The detailed results of causal direction and sensitivity analysis between omega-3-pct and IA. A: scatter plot; B: funnel plot; C: forest plot; D: leave-one-out analysis.

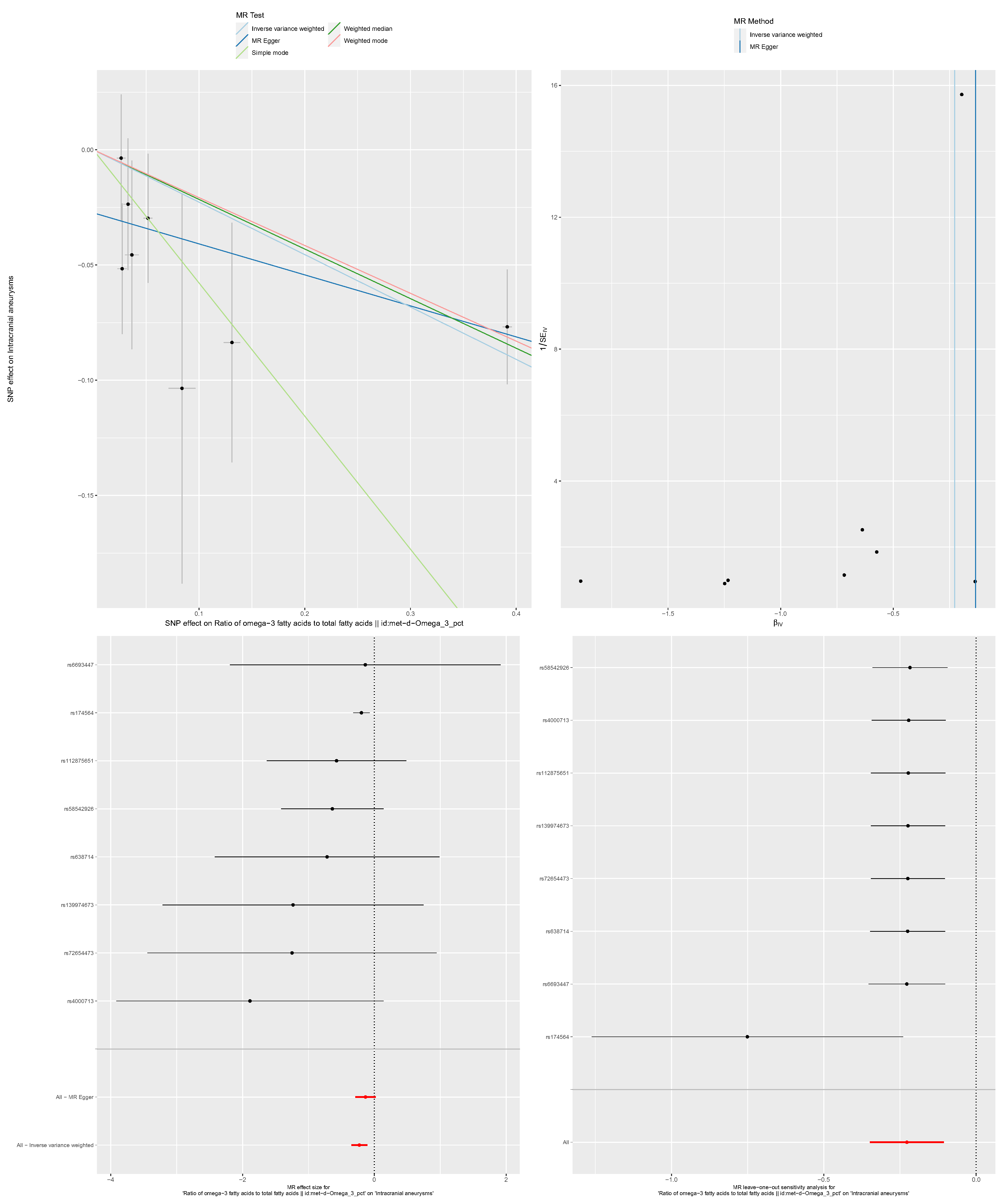

Supply figure 3-4. The detailed results of causal direction and sensitivity analysis between omega-6 by omega-3 and IA. A: scatter plot; B: funnel plot; C: forest plot; D: leave-one-out analysis.

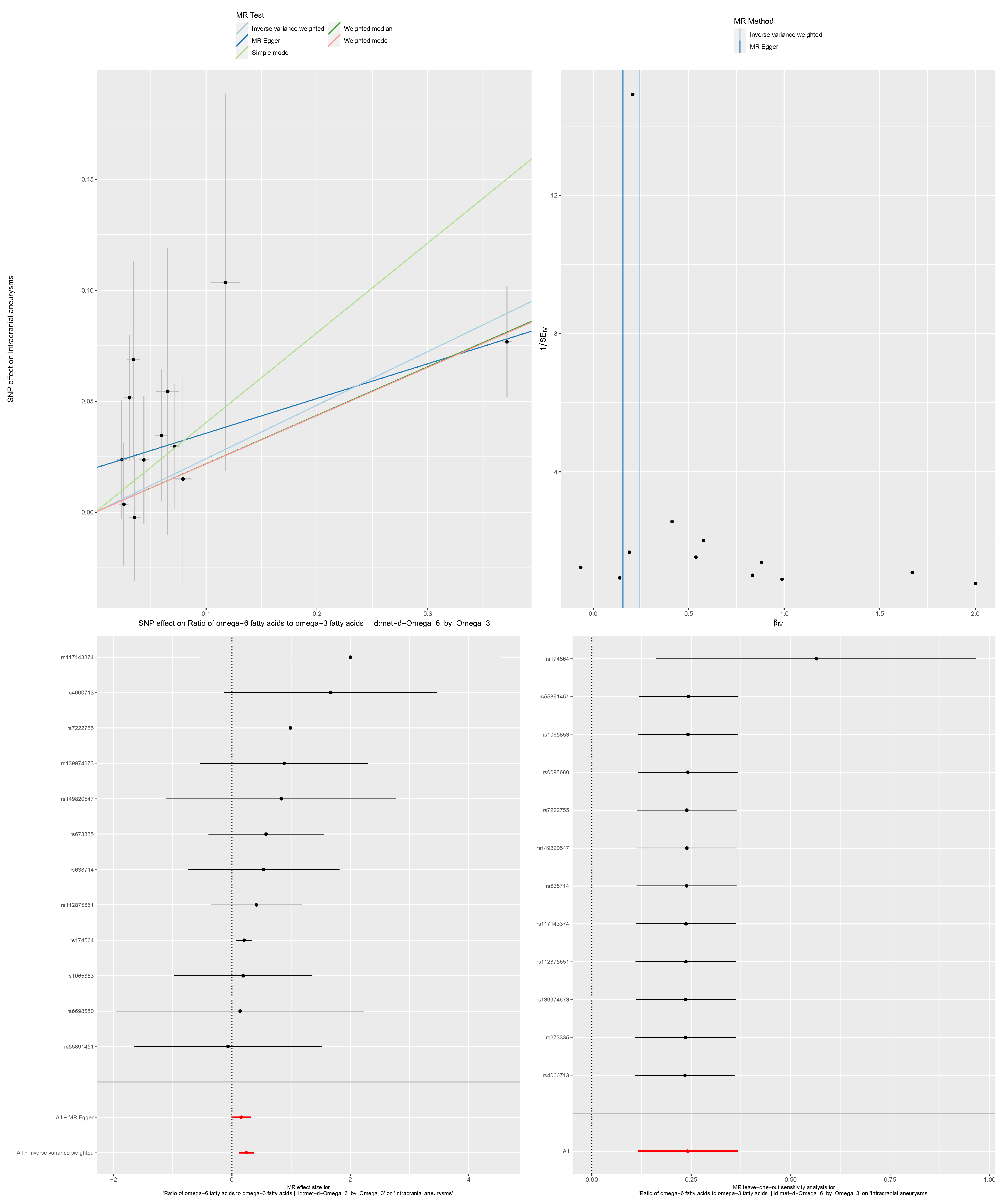

Supply figure 3-5. The detailed results of causal direction and sensitivity analysis between omega-6 and IA. A: scatter plot; B: funnel plot; C: forest plot; D: leave-one-out analysis.

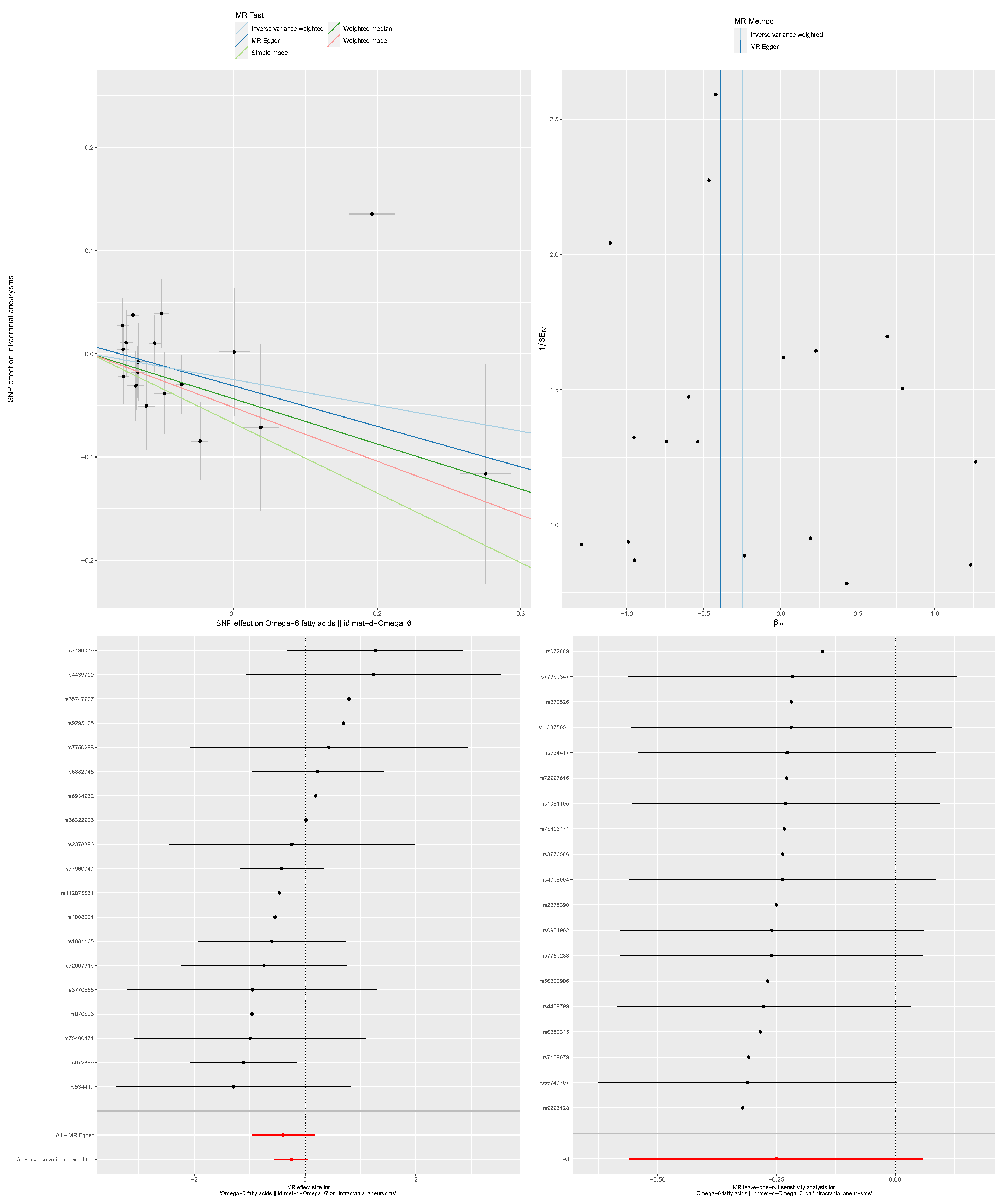

Supply figure 4-1. The detailed results of causal direction and sensitivity analysis between omega-3 and aSAH. A: scatter plot; B: funnel plot; C: forest plot; D: leave-one-out analysis.

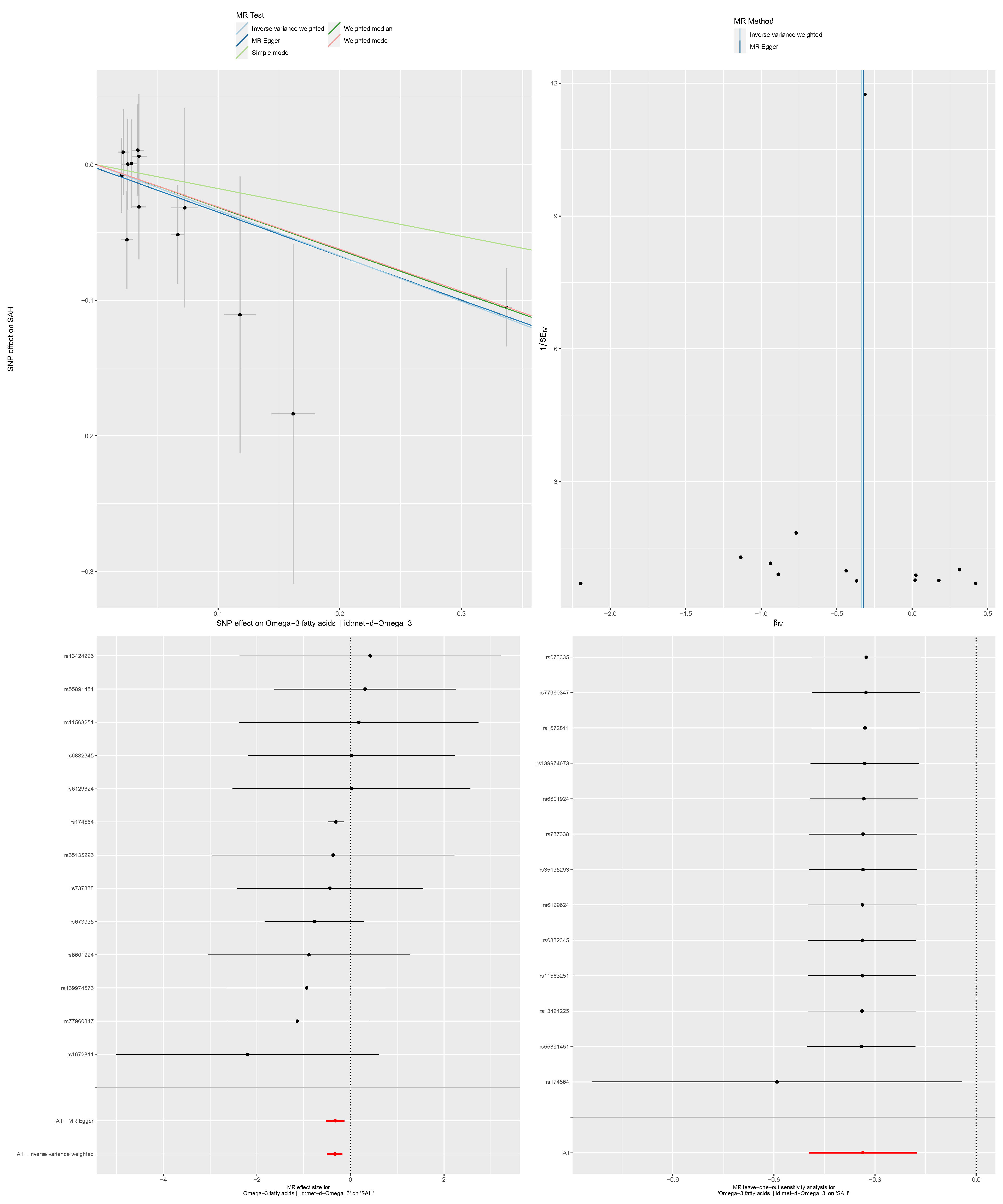

Supply figure 4-2. The detailed results of causal direction and sensitivity analysis between DHA and aSAH. A: scatter plot; B: funnel plot; C: forest plot; D: leave-one-out analysis.

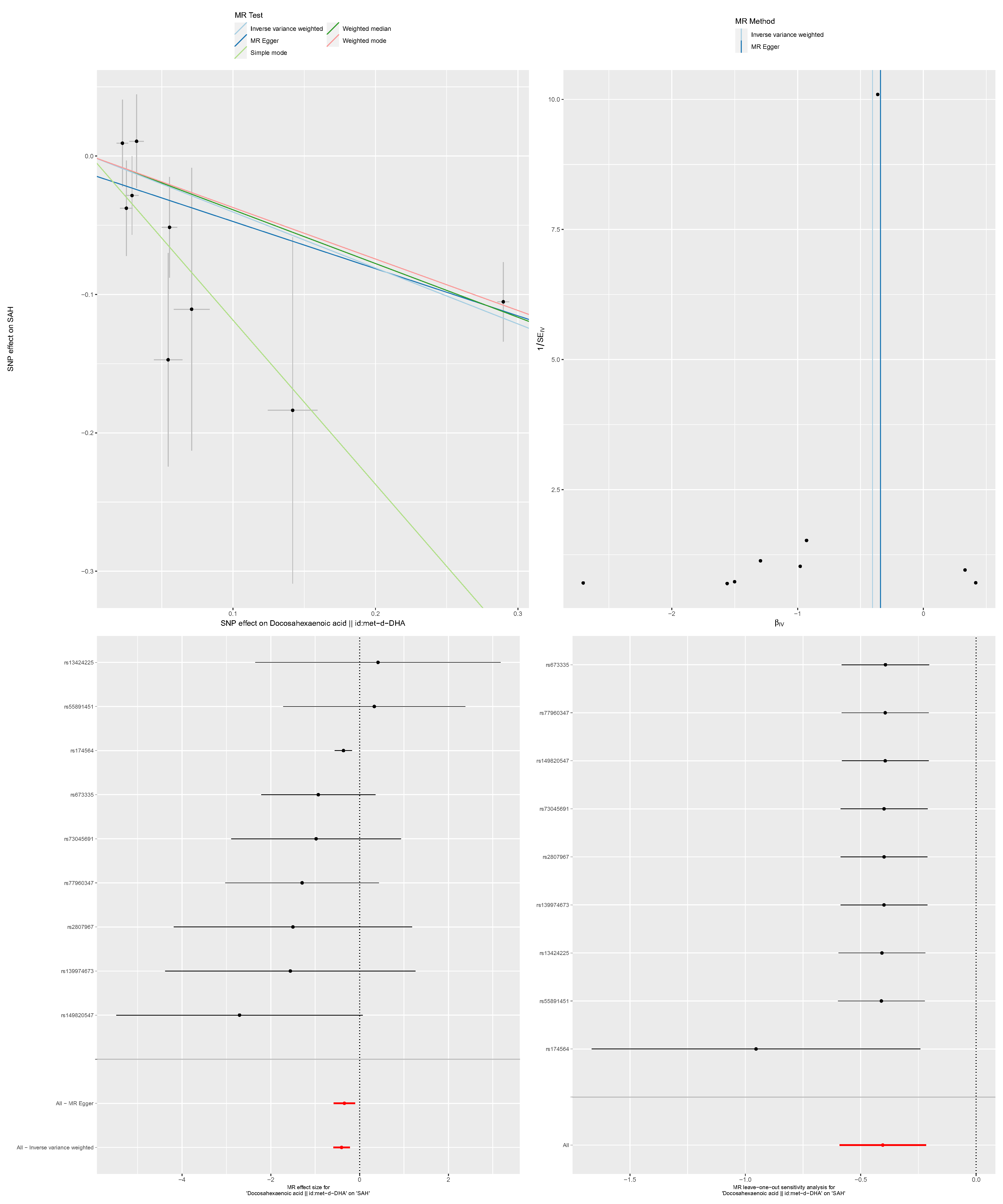

Supply figure 4-3. The detailed results of causal direction and sensitivity analysis between omega-3-pct and aSAH. A: scatter plot; B: funnel plot; C: forest plot; D: leave-one-out analysis.

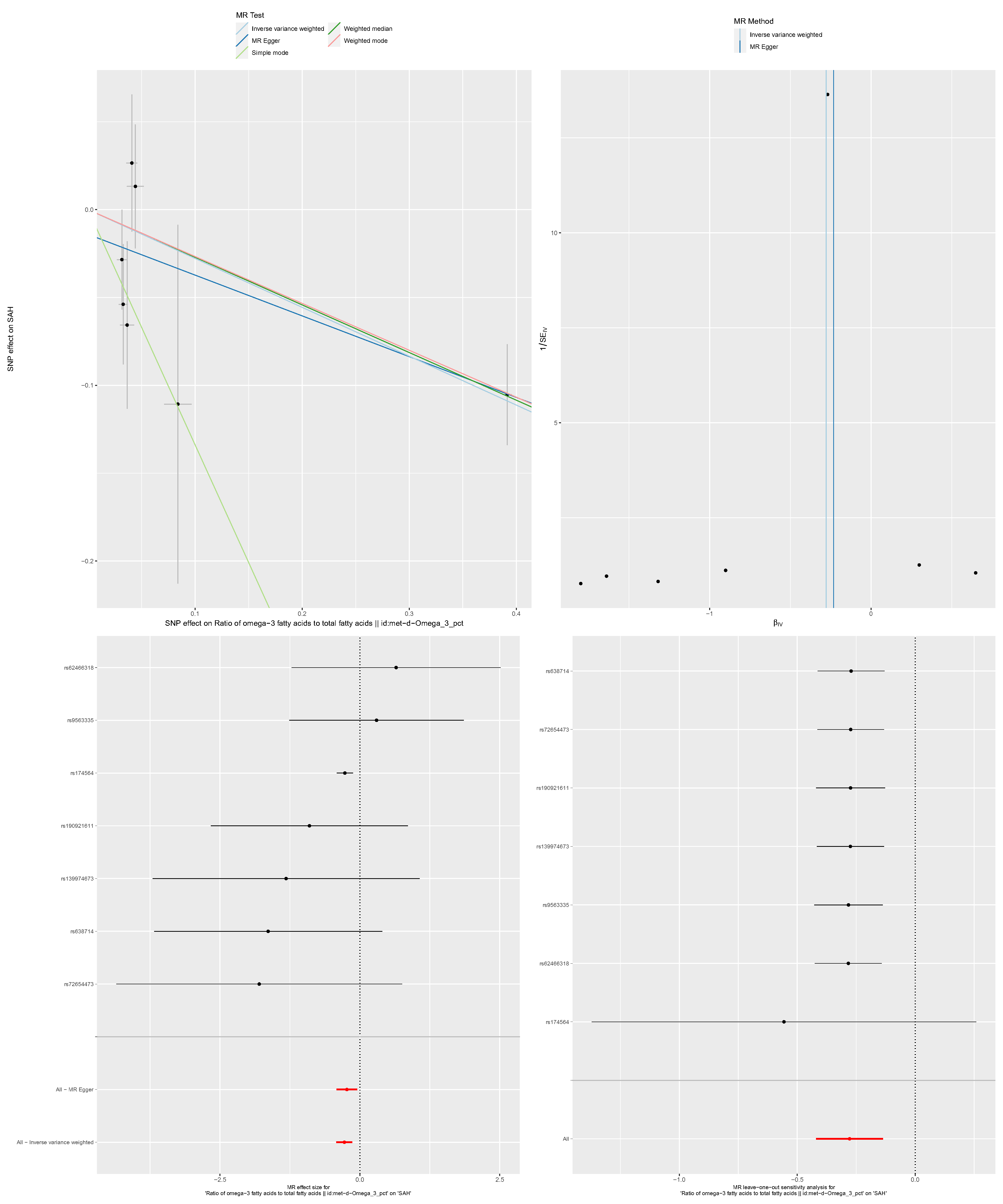

Supply figure 4-4. The detailed results of causal direction and sensitivity analysis between omega-6 by omega-3 and aSAH. A: scatter plot; B: funnel plot; C: forest plot; D: leave-one-out analysis.

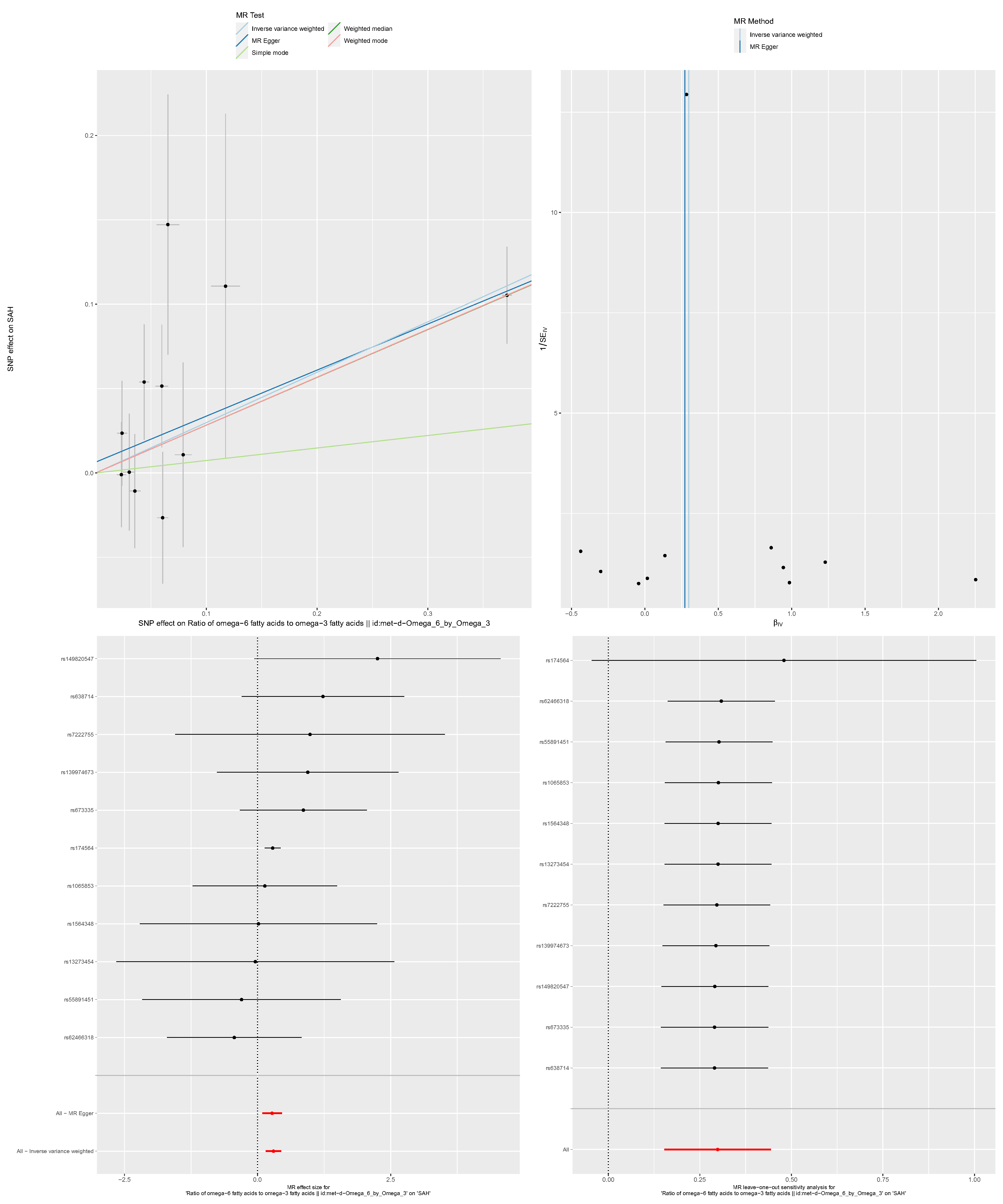

Supply figure 4-5. The detailed results of causal direction and sensitivity analysis between omega-6 and aSAH. A: scatter plot; B: funnel plot; C: forest plot; D: leave-one-out analysis.

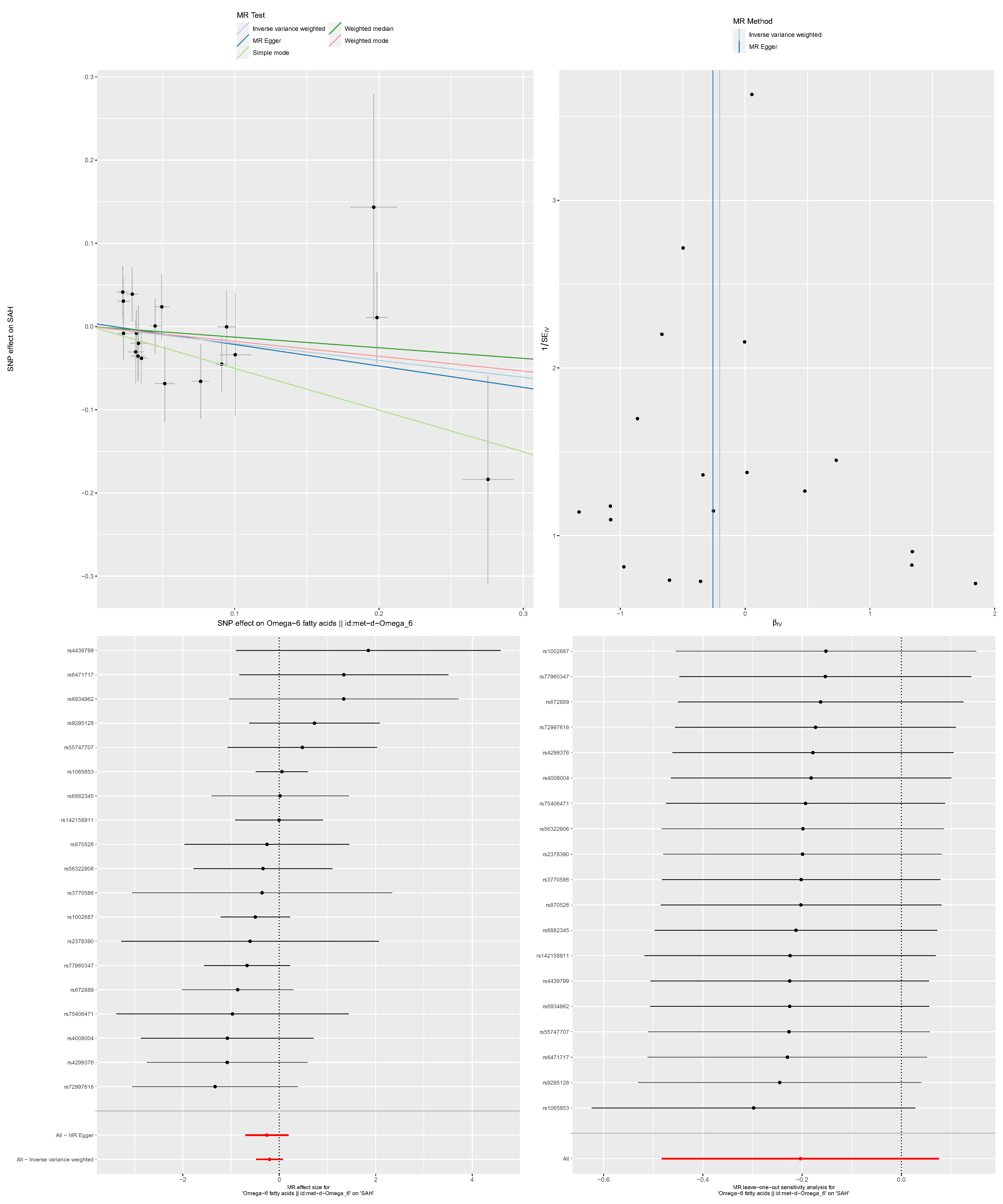

Supply figure 5-1. The detailed results of causal direction and sensitivity analysis between omega-3 and uIA. A: scatter plot; B: funnel plot; C: forest plot; D: leave-one-out analysis.

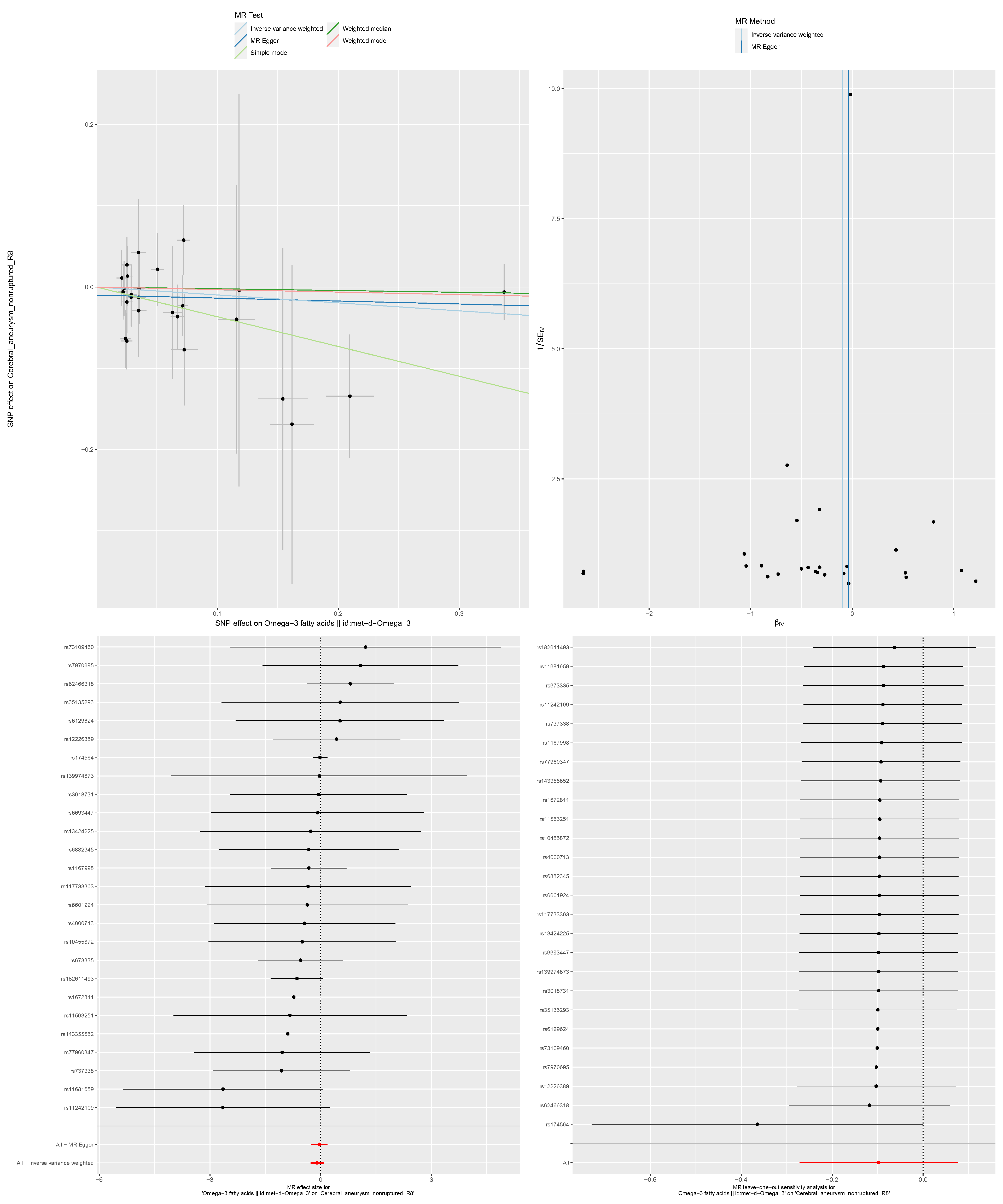

Supply figure 5-2. The detailed results of causal direction and sensitivity analysis between DHA and uIA. A: scatter plot; B: funnel plot; C: forest plot; D: leave-one-out analysis.

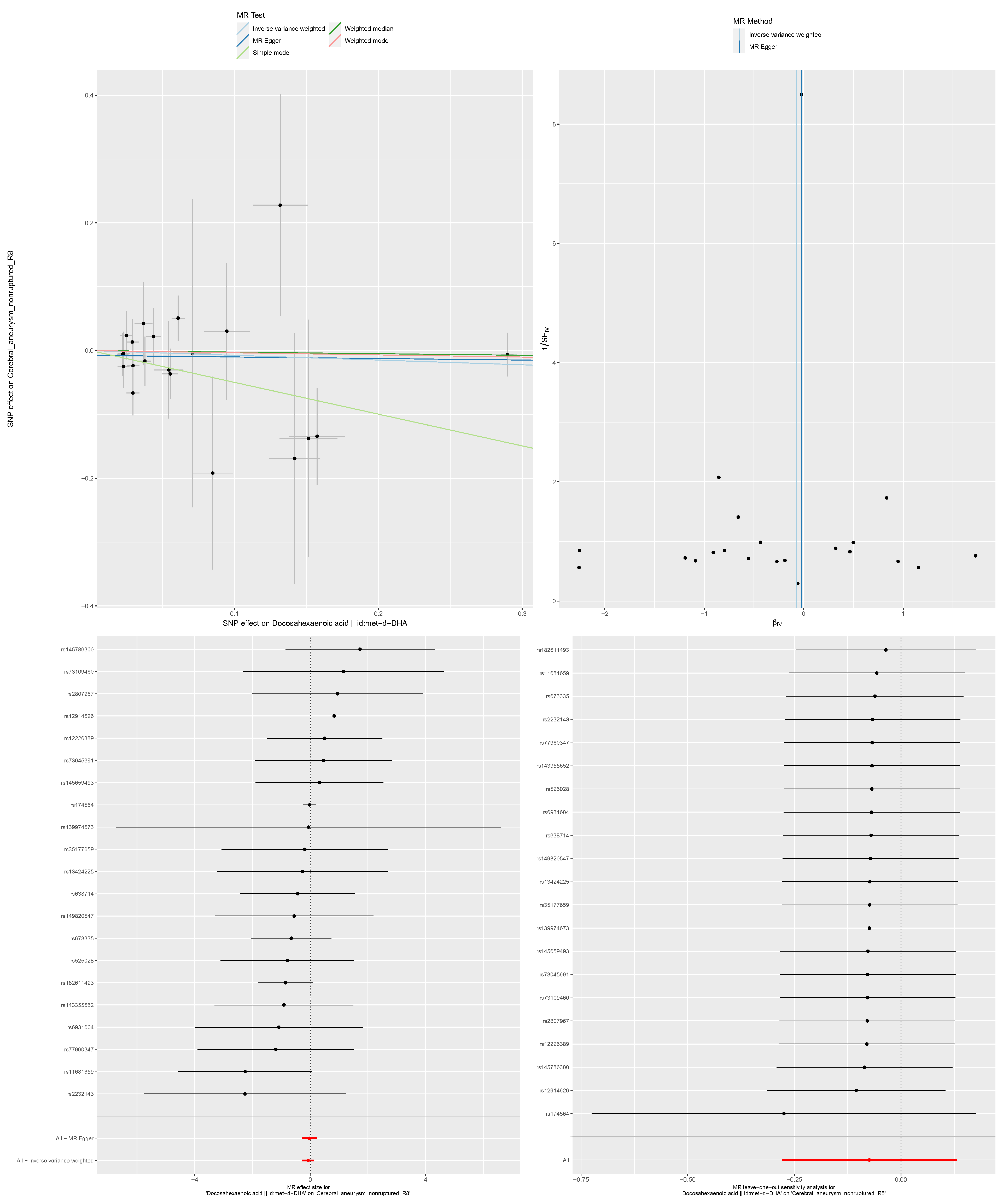

Supply figure 5-3. The detailed results of causal direction and sensitivity analysis between omega-3-pct and uIA. A: scatter plot; B: funnel plot; C: forest plot; D: leave-one-out analysis.

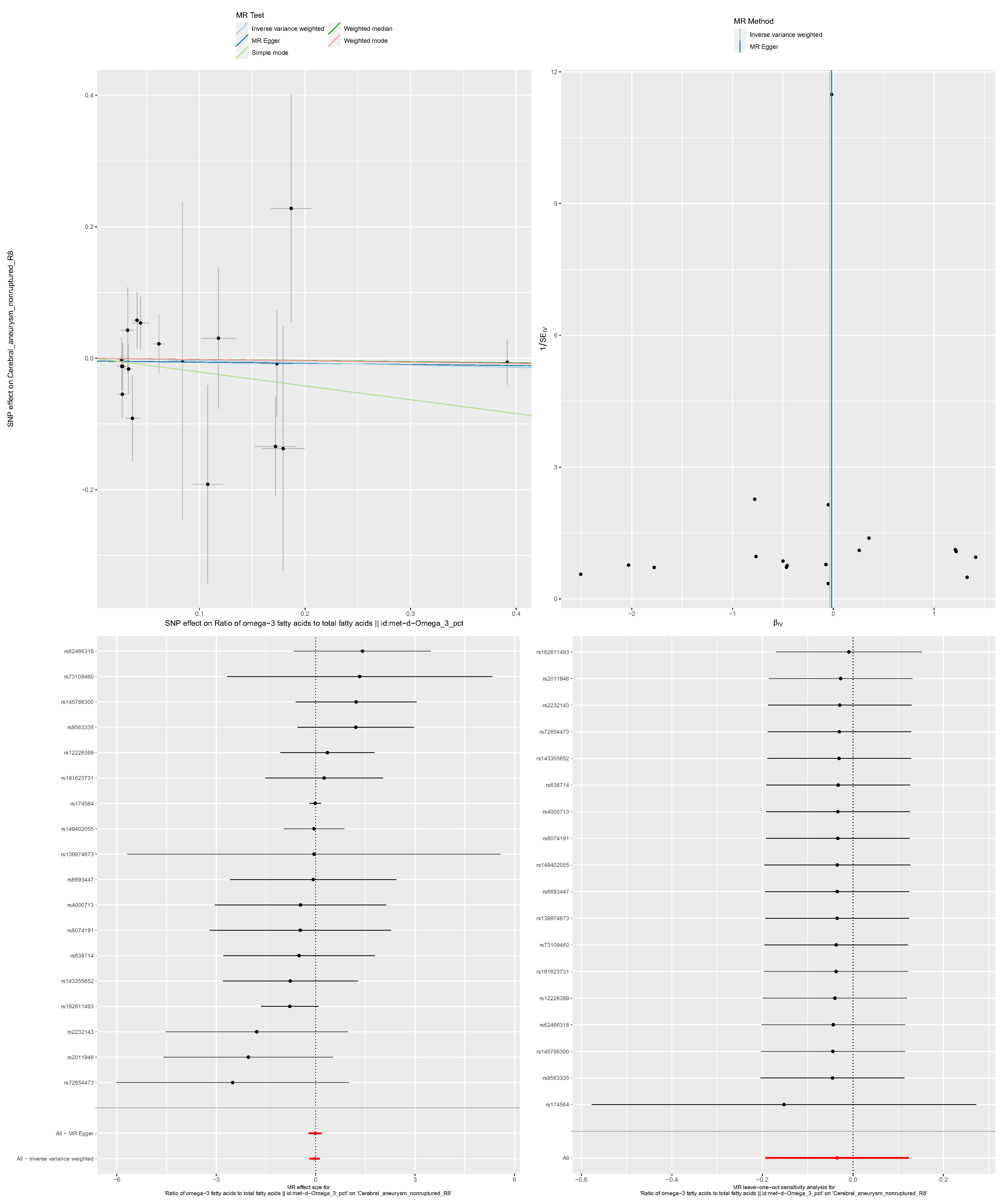

Supply figure 5-4. The detailed results of causal direction and sensitivity analysis between omega-6 by omega-3 and uIA. A: scatter plot; B: funnel plot; C: forest plot; D: leave-one-out analysis.

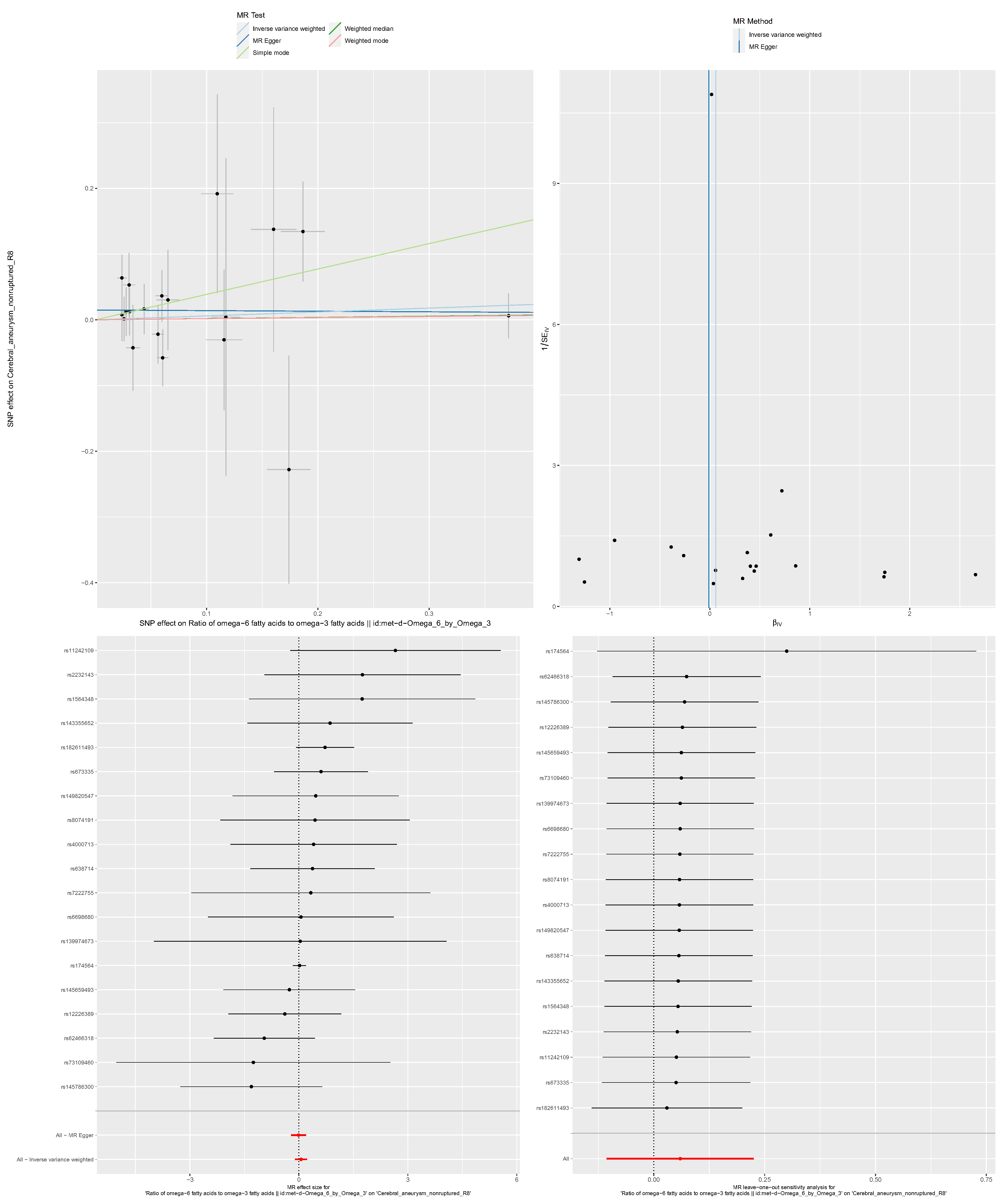

Supply figure 5-5. The detailed results of causal direction and sensitivity analysis between omega-6 and uIA. A: scatter plot; B: funnel plot; C: forest plot; D: leave-one-out analysis.

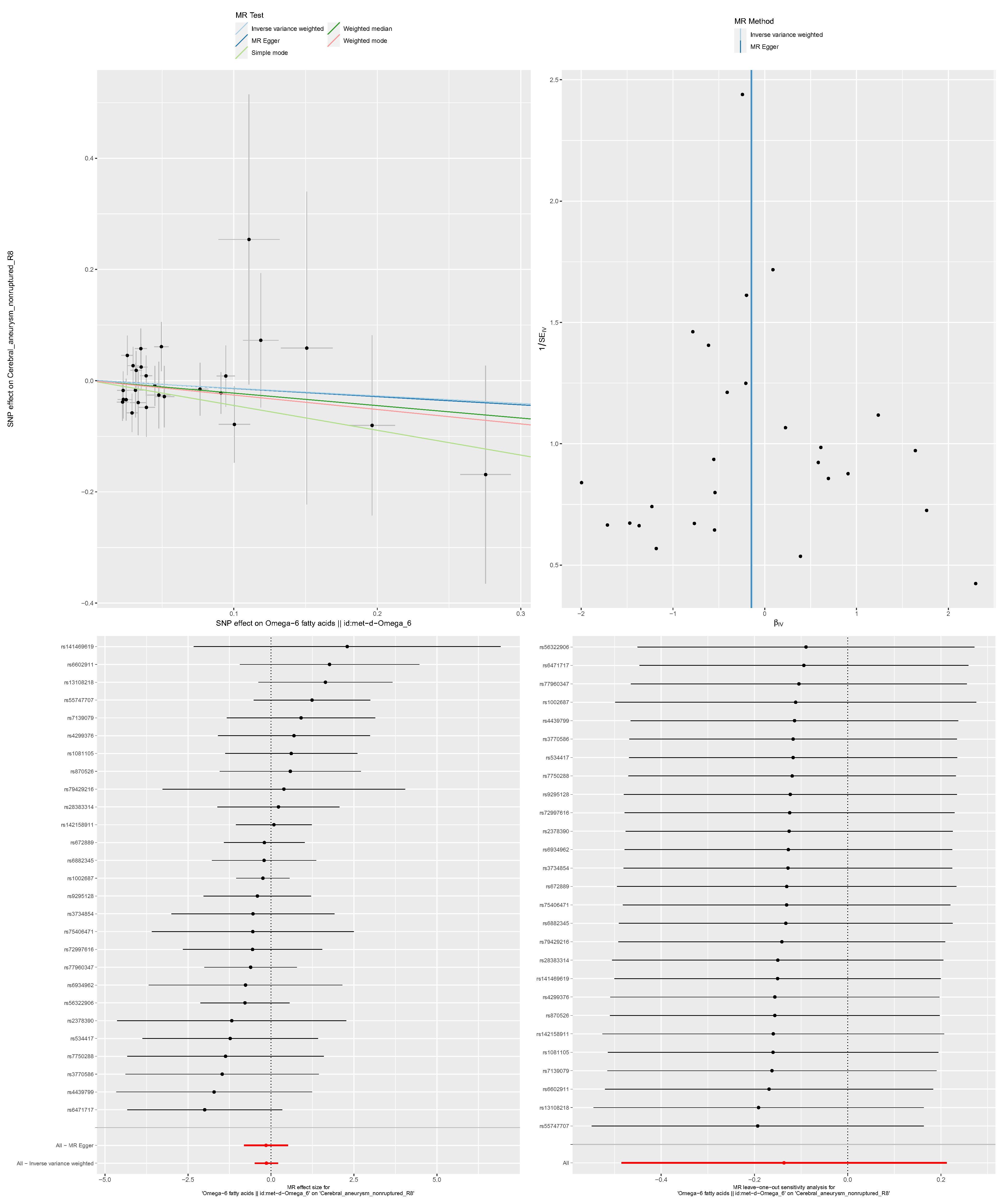
