## Supplementary material for "Predicting the causal relationship between polyunsaturated fatty acids and cerebral aneurysm risk from a Mendelian randomization study": Supplymentary table

**Supplementary table**

1 The detailed information of all GWAS databases in this study.

2	Characteristics of instrumental variables (IVs) associated with intracranial aneurysm in the PUFAs (Include 2_1-5)

3	Characteristics of instrumental variables (IVs) associated with subarachnoid hemorrhage in the PUFAs (Include 3_1-5)

4	Characteristics of instrumental variables (IVs) associated with unruptured intracranial aneurysm in the PUFAs (Include 4_1-5)

5	Sensitivity analysis of forward causal effect between fatty acid and the risk of aneurysm.

6	The result of Bayesian colocalization analysis between fatty acid and aneurysm (Include 6_1-8)

| Supplementary Table 1. The detailed information of all GWAS databases in this study. | | | | | | | |
| --- | --- | --- | --- | --- | --- | --- | --- |
| **Trait** | **Sample size** | **Case** | **Control** | **Population** | **Study** | **GWAS ID** | **Data sources** |
| Omega-3 fatty acids | 114999 | - | - | European | UKBB | met-d-Omega_3 | <https://gwas.mrcieu.ac.uk/datasets/met-d-Omega_3/> |
|  |  |  |  |  |  |  | doi: 10.1186/s12916-022-02399-w |
| Ratio of omega-3 fatty acids to total fatty acids | 114999 | - | - | European | UKBB | met-d-Omega_3_pct | https://gwas.mrcieu.ac.uk/datasets/met-d-Omega_3_pct/ |
|  |  |  |  |  |  |  | doi: 10.1186/s12916-022-02399-w |
| Ratio of omega-6 fatty acids to omega-3 fatty acids | 114999 | - | - | European | UKBB | met-d-Omega_6_by_Omega_3 | https://gwas.mrcieu.ac.uk/datasets/met-d-Omega_6_by_Omega_3/ |
|  |  |  |  |  |  |  | doi: 10.1186/s12916-022-02399-w |
| Docosahexaenoic acid | 114999 | - | - | European | UKBB | met-d-DHA | https://gwas.mrcieu.ac.uk/datasets/met-d-DHA/ |
|  |  |  |  |  |  |  | doi: 10.1186/s12916-022-02399-w |
| Omega-6 fatty acids | 114999 | - | - | European | UKBB | met-d-Omega_6 | <https://gwas.mrcieu.ac.uk/datasets/met-d-Omega_6/> |
|  |  |  |  |  |  |  | doi: 10.1186/s12916-022-02399-w |
| Intracranial aneurysm | 65796 | 6252 | 59544 | European | excludingUKBB | - | [http://www.cerebrovascularportal.org.](http://www.cerebrovascularportal.org./) |
|  |  |  |  |  |  |  | DOI：10.1038/s41588-020-00725-7 |
| subarachnoid hemorrhage | 63740 | 4196 | 59544 | European | excludingUKBB | - | [http://www.cerebrovascularportal.org.](http://www.cerebrovascularportal.org./) |
|  |  |  |  |  |  |  | DOI：10.1038/s41588-020-00725-7 |
| Unruptured intracranial aneurysm | 314556 | 1788 | 312768 | European | Finngen | - | https://r8.finngen.fi/pheno/I9_ANEURYSM |
| HDL cholesterol | 187167 | - | - | European | GLGC | ieu-a-299 | https://www.nature.com/articles/ng.2797 |
|  |  |  |  |  |  |  | DOI: 10.1038/ng.2797 |
| LDL cholesterol | 173082 | - | - | European | GLGC | ieu-a-300 | https://www.nature.com/articles/ng.2797 |
|  |  |  |  |  |  |  | DOI: 10.1038/ng.2797 |
| Total cholesterol | 187365 | - | - | European | GLGC | ieu-a-301 | https://www.nature.com/articles/ng.2797 |
|  |  |  |  |  |  |  | DOI: 10.1038/ng.2797 |
| Triglycerides | 177861 | - | - | European | GLGC | ieu-a-302 | https://www.nature.com/articles/ng.2797 |
|  |  |  |  |  |  |  | DOI: 10.1038/ng.2797 |

Supplementary Table 2_1. Characteristics of instrumental variables (IVs) associated with intracranial aneurysm in the omega-3

| **SNP** | **Nearest gene** | **Chromosome** | **Position (37/38)** | **Effect Allele** | **Other Allele** | **EAF** | **Beta** | **SE** | **P-value** | **Remove** |
| --- | --- | --- | --- | --- | --- | --- | --- | --- | --- | --- |
| rs112875651 | — | 8 | 126506694 | G | A | 0.608 | 0.087 | 0.004 | 3.50E-98 | Remove (confounders) |
| rs11563251 | UGT1A1, UGT1A3, UGT1A4, UGT1A5,  UGT1A6, UGT1A7, UGT1A8, UGT1A9, UGT1A10 | 2 | 234679384 | T | C | 0.111 | 0.035 | 0.006 | 3.20E-08 |  |
| rs1167998 | DOCK7 | 1 | 62931632 | A | C | 0.645 | 0.071 | 0.004 | 3.60E-66 | Remove (confounders) |
| rs117143374 | BRWD1, PSMG1 | 21 | 40555561 | T | C | 0.858 | 0.037 | 0.006 | 2.20E-10 |  |
| rs117733303 | LPAL2 | 6 | 160922870 | A | G | 0.981 | 0.116 | 0.015 | 1.40E-15 |  |
| rs1260326 | GCKR | 2 | 27730940 | T | C | 0.396 | 0.082 | 0.004 | 8.40E-88 | Remove (confounders) |
| rs13424225 | — | 2 | 241214158 | T | G | 0.450 | 0.022 | 0.004 | 2.20E-08 |  |
| rs139974673 | CATSPER2P1 | 15 | 44027885 | C | T | 0.026 | 0.118 | 0.013 | 2.30E-21 |  |
| rs174564 | FADS2 | 11 | 61588305 | A | G | 0.653 | 0.337 | 0.004 | 1.00E-200 | Remove (FADS2 )* |
| rs261290 | — | 15 | 58678720 | T | C | 0.345 | 0.114 | 0.004 | 3.90E-161 | Remove (confounders) |
| rs35135293 | — | 2 | 20363666 | C | T | 0.483 | 0.021 | 0.004 | 3.90E-08 |  |
| rs4000713 | MIR148A | 7 | 25990597 | G | A | 0.705 | 0.029 | 0.004 | 1.00E-11 |  |
| rs58542926 | TM6SF2 | 19 | 19379549 | C | T | 0.926 | 0.172 | 0.008 | 1.40E-113 | Remove (confounders) |
| rs6129624 | — | 20 | 39167592 | G | A | 0.665 | 0.026 | 0.004 | 5.10E-10 |  |
| rs62466318 | — | 7 | 73042085 | C | T | 0.796 | 0.072 | 0.005 | 1.20E-45 |  |
| rs629301 | CELSR2 | 1 | 109818306 | T | G | 0.778 | 0.038 | 0.005 | 1.30E-14 | Remove (confounders) |
| rs633695 | LIPC, LOC101928694 | 15 | 58725839 | G | A | 0.292 | 0.084 | 0.004 | 9.10E-80 | Remove (confounders) |
| rs6601924 | AKR1C4 | 10 | 5247302 | C | T | 0.846 | 0.035 | 0.006 | 8.50E-10 |  |
| rs673335 | — | 11 | 75450576 | T | C | 0.840 | 0.067 | 0.006 | 1.10E-34 |  |
| rs6882345 | — | 5 | 156397673 | A | G | 0.633 | 0.029 | 0.004 | 1.90E-13 |  |
| rs737338 | C19orf80, DOCK6 | 19 | 11347657 | C | T | 0.965 | 0.073 | 0.011 | 3.50E-11 |  |
| rs77960347 | LIPG | 18 | 47109955 | G | A | 0.013 | 0.162 | 0.018 | 7.20E-22 |  |
| rs7819706 | — | 8 | 19844415 | A | G | 0.882 | 0.040 | 0.006 | 1.80E-10 | Remove (confounders) |
| rs7924036 | JMJD1C | 10 | 65191645 | T | G | 0.504 | 0.023 | 0.004 | 5.50E-10 | Remove (confounders) |
| rs7970695 | HNF1A | 12 | 121423376 | G | A | 0.379 | 0.025 | 0.004 | 1.20E-10 | Remove (radial MR) |
| rs9304381 | — | 18 | 47158234 | T | C | 0.818 | 0.053 | 0.005 | 5.20E-24 | Remove (confounders) |

*Sensitivity analysis after the removal of the FADS2 and its SNPs within 500 kb of the FADS2 locus.

Supplementary Table 2_2. Characteristics of instrumental variables (IVs) associated with intracranial aneurysm in the omega-3-pct

| **SNP** | **Nearest gene** | **Chromosome** | **Position (37/38)** | **Effect Allele** | **Other Allele** | **EAF** | **Beta** | **SE** | **P-value** | **Remove** |
| --- | --- | --- | --- | --- | --- | --- | --- | --- | --- | --- |
| rs112875651 | — | 8 | 126506694 | G | A | 0.608 | 0.052 | 0.004 | 5.70E-35 |  |
| rs1260326 | GCKR | 2 | 27730940 | T | C | 0.396 | 0.038 | 0.004 | 2.30E-19 | Remove (confounders) |
| rs139974673 | CATSPER2P1 | 15 | 44027885 | C | T | 0.026 | 0.084 | 0.013 | 1.30E-11 |  |
| rs1560390 | — | 15 | 58580781 | T | C | 0.780 | 0.032 | 0.005 | 6.90E-12 | Remove (confounders) |
| rs174564 | FADS2 | 11 | 61588305 | A | G | 0.653 | 0.392 | 0.004 | 1.00E-200 | Remove (FADS2 )* |
| rs2394976 | — | 6 | 31311912 | G | T | 0.838 | 0.031 | 0.006 | 5.40E-09 | Remove (radial MR) |
| rs261291 | — | 15 | 58680178 | C | T | 0.356 | 0.081 | 0.004 | 1.80E-83 | Remove (confounders) |
| rs272888 | LOC553103, SLC22A4 | 5 | 131665423 | C | T | 0.707 | 0.029 | 0.004 | 1.80E-11 | Remove (radial MR) |
| rs4000713 | MIR148A | 7 | 25990597 | G | A | 0.705 | 0.027 | 0.004 | 1.60E-10 |  |
| rs58542926 | TM6SF2 | 19 | 19379549 | C | T | 0.926 | 0.131 | 0.008 | 1.20E-67 |  |
| rs62466318 | — | 7 | 73042085 | C | T | 0.796 | 0.041 | 0.005 | 3.10E-16 | Remove (radial MR) |
| rs638714 | USP1 | 1 | 62906489 | G | T | 0.654 | 0.033 | 0.004 | 4.90E-15 |  |
| rs6693447 | RER1 | 1 | 2330190 | G | T | 0.462 | 0.026 | 0.004 | 8.30E-12 |  |
| rs72654473 | APOE | 19 | 45414399 | A | C | 0.108 | 0.037 | 0.007 | 6.10E-09 |  |
| rs7444298 | LOC102546226, TMEM161B-AS1 | 5 | 87730027 | G | A | 0.237 | 0.027 | 0.005 | 2.40E-08 | Remove (confounders) |
| rs7924036 | JMJD1C | 10 | 65191645 | T | G | 0.504 | 0.038 | 0.004 | 4.00E-23 | Remove (confounders) |
| rs9947684 | — | 18 | 47166694 | G | A | 0.654 | 0.029 | 0.004 | 2.80E-12 | Remove (confounders) |

*Sensitivity analysis after the removal of the FADS2 and its SNPs within 500 kb of the FADS2 locus.

Supplementary Table 2_3. Characteristics of instrumental variables (IVs) associated with intracranial aneurysm in the DHA

| **SNP** | **Nearest gene** | **Chromosome** | **Position (37/38)** | **Effect Allele** | **Other Allele** | **EAF** | **Beta** | **SE** | **P-value** | **Remove** |
| --- | --- | --- | --- | --- | --- | --- | --- | --- | --- | --- |
| rs11122450 | GALNT2 | 1 | 230301811 | G | T | 0.611 | 0.023 | 0.004 | 3.40E-08 | Remove (confounders) |
| rs112875651 | — | 8 | 126506694 | G | A | 0.608 | 0.049 | 0.004 | 1.60E-32 |  |
| rs1260326 | GCKR | 2 | 27730940 | T | C | 0.396 | 0.047 | 0.004 | 8.70E-30 | Remove (confounders) |
| rs13424225 | — | 2 | 241214158 | T | G | 0.450 | 0.022 | 0.004 | 1.10E-08 |  |
| rs139974673 | CATSPER2P1 | 15 | 44027885 | C | T | 0.026 | 0.071 | 0.013 | 8.80E-09 |  |
| rs149820547 | SCGB2A1 | 11 | 61983775 | T | G | 0.958 | 0.054 | 0.010 | 4.80E-08 | Remove (FADS2 locus)* |
| rs1560390 | — | 15 | 58580781 | T | C | 0.780 | 0.050 | 0.005 | 2.50E-26 | Remove (confounders) |
| rs174564 | FADS2 | 11 | 61588305 | A | G | 0.653 | 0.290 | 0.004 | 1.00E-200 | Remove (FADS2 )* |
| rs2278426 | C19orf80, DOCK6 | 19 | 11350488 | C | T | 0.965 | 0.060 | 0.011 | 3.70E-08 | Remove (confounders) |
| rs261291 | — | 15 | 58680178 | C | T | 0.356 | 0.115 | 0.004 | 5.40E-172 | Remove (confounders) |
| rs273912 | LOC553103, SLC22A4 | 5 | 131661349 | T | G | 0.707 | 0.026 | 0.004 | 6.60E-10 | Remove (radial MR) |
| rs325 | LPL | 8 | 19819328 | C | T | 0.100 | 0.039 | 0.007 | 3.80E-09 | Remove (confounders) |
| rs3764261 | CETP | 16 | 56993324 | A | C | 0.324 | 0.039 | 0.004 | 1.90E-21 | Remove (confounders) |
| rs525028 | APOA1, APOC3 | 11 | 116705516 | G | A | 0.291 | 0.030 | 0.004 | 2.70E-12 |  |
| rs58542926 | TM6SF2 | 19 | 19379549 | C | T | 0.926 | 0.119 | 0.008 | 1.40E-57 | Remove (confounders) |
| rs629301 | CELSR2 | 1 | 109818306 | T | G | 0.778 | 0.044 | 0.005 | 1.40E-19 | Remove (confounders) |
| rs638714 | USP1 | 1 | 62906489 | G | T | 0.654 | 0.038 | 0.004 | 5.40E-20 |  |
| rs673335 | — | 11 | 75450576 | T | C | 0.840 | 0.055 | 0.005 | 8.10E-25 |  |
| rs6931604 | — | 6 | 98578215 | T | C | 0.600 | 0.023 | 0.004 | 2.70E-08 |  |
| rs73045691 | — | 19 | 45440529 | A | G | 0.304 | 0.029 | 0.005 | 3.60E-10 |  |
| rs7412 | APOE | 19 | 45412079 | C | T | 0.919 | 0.077 | 0.007 | 5.40E-26 | Remove (confounders) |
| rs77960347 | LIPG | 18 | 47109955 | G | A | 0.013 | 0.142 | 0.017 | 2.50E-17 |  |
| rs7924036 | JMJD1C | 10 | 65191645 | T | G | 0.504 | 0.036 | 0.004 | 4.80E-21 | Remove (confounders) |
| rs9304381 | — | 18 | 47158234 | T | C | 0.818 | 0.050 | 0.005 | 5.80E-23 | Remove (confounders) |

*Sensitivity analysis after the removal of the FADS2 and its SNPs within 500 kb of the FADS2 locus.

Supplementary Table 2_4. Characteristics of instrumental variables (IVs) associated with intracranial aneurysm in the omega-6

| **SNP** | **Nearest gene** | **Chromosome** | **Position (37/38)** | **Effect Allele** | **Other Allele** | **EAF** | **Beta** | **SE** | **P-value** | **Remove** |
| --- | --- | --- | --- | --- | --- | --- | --- | --- | --- | --- |
| rs1002687 | DOCK7 | 1 | 62963737 | A | G | 0.645 | 0.091 | 0.004 | 1.00E-107 | Remove (confounders) |
| rs1065853 | APOC1, APOE, TOMM40 | 19 | 45413233 | G | T | 0.919 | 0.199 | 0.007 | 3.60E-160 | Remove (confounders) |
| rs1081105 | APOC1, APOE, TOMM40 | 19 | 45412955 | C | A | 0.028 | 0.119 | 0.012 | 1.80E-22 |  |
| rs112875651 | — | 8 | 126506694 | G | A | 0.608 | 0.064 | 0.004 | 2.20E-53 |  |
| rs11789603 | ABCA1 | 9 | 107647019 | T | C | 0.109 | 0.048 | 0.006 | 9.70E-14 | Remove (confounders) |
| rs1260326 | GCKR | 2 | 27730940 | T | C | 0.396 | 0.064 | 0.004 | 3.90E-55 | Remove (confounders) |
| rs12740374 | CELSR2, PSRC1 | 1 | 109817590 | G | T | 0.779 | 0.057 | 0.005 | 1.50E-32 | Remove (confounders) |
| rs142158911 | LDLR | 19 | 11190534 | G | A | 0.883 | 0.094 | 0.006 | 5.20E-52 | Remove (confounders) |
| rs1800961 | HNF4A, MIR3646 | 20 | 43042364 | C | T | 0.970 | 0.074 | 0.012 | 3.30E-10 | Remove (confounders) |
| rs183130 | CETP | 16 | 56991363 | T | C | 0.324 | 0.062 | 0.004 | 1.40E-48 | Remove (confounders) |
| rs2378390 | ERGIC3, FER1L4 | 20 | 34150207 | G | A | 0.859 | 0.033 | 0.006 | 3.20E-09 |  |
| rs261290 | — | 15 | 58678720 | T | C | 0.345 | 0.097 | 0.004 | 1.00E-116 | Remove (confounders) |
| rs2737245 | TRPS1 | 8 | 116658583 | G | T | 0.721 | 0.027 | 0.005 | 1.40E-09 | Remove (confounders) |
| rs2740488 | ABCA1 | 9 | 107661742 | A | C | 0.735 | 0.050 | 0.005 | 5.40E-28 | Remove (confounders) |
| rs3770586 | ABCB11 | 2 | 169828995 | C | T | 0.516 | 0.023 | 0.004 | 7.10E-09 |  |
| rs4008004 | TTC39B | 9 | 15300968 | A | C | 0.222 | 0.033 | 0.005 | 8.40E-12 |  |
| rs4299376 | ABCG5, ABCG8 | 2 | 44072576 | G | T | 0.324 | 0.035 | 0.004 | 1.10E-16 | Remove (confounders) |
| rs4439799 | TBKBP1 | 17 | 45781599 | T | C | 0.502 | 0.022 | 0.004 | 1.30E-08 |  |
| rs534417 | ASAP3 | 1 | 23784965 | G | A | 0.875 | 0.039 | 0.006 | 9.30E-11 |  |
| rs55747707 | MLXIPL | 7 | 73037366 | G | A | 0.796 | 0.049 | 0.005 | 1.70E-22 |  |
| rs56322906 | C19orf80, DOCK6 | 19 | 11346155 | G | A | 0.965 | 0.100 | 0.011 | 1.20E-19 |  |
| rs5754102 | UBE2L3 | 22 | 21916272 | C | A | 0.817 | 0.032 | 0.005 | 9.90E-10 | Remove (confounders) |
| rs58542926 | HAPLN4, SUGP1, TM6SF2 | 19 | 19379549 | C | T | 0.926 | 0.128 | 0.008 | 2.50E-65 | Remove (confounders) |
| rs633695 | LIPC, LOC101928694 | 15 | 58725839 | G | A | 0.292 | 0.073 | 0.004 | 1.30E-59 | Remove (confounders) |
| rs6471717 | — | 8 | 59377357 | G | A | 0.337 | 0.029 | 0.004 | 4.00E-12 | Remove (confounders) |
| rs6547409 | — | 2 | 21190209 | C | T | 0.949 | 0.081 | 0.009 | 2.40E-20 | Remove (confounders) |
| rs672889 | — | 2 | 21319016 | G | T | 0.860 | 0.076 | 0.006 | 1.30E-41 |  |
| rs6882345 | TIMD4 | 5 | 156397673 | A | G | 0.633 | 0.045 | 0.004 | 1.20E-27 |  |
| rs6934962 | FRK | 6 | 116322349 | T | C | 0.400 | 0.023 | 0.004 | 2.20E-08 |  |
| rs7139079 | HNF1A, HNF1A-AS1 | 12 | 121415293 | G | A | 0.407 | 0.030 | 0.004 | 3.30E-13 |  |
| rs72997616 | DGAT2, LOC283214 | 11 | 75474195 | C | A | 0.906 | 0.052 | 0.007 | 1.60E-13 |  |
| rs75406471 | AKR1C4 | 10 | 5257647 | G | A | 0.845 | 0.031 | 0.006 | 2.70E-08 |  |
| rs7750288 | IGF2R | 6 | 160400147 | G | A | 0.285 | 0.025 | 0.004 | 1.30E-08 |  |
| rs77960347 | LIPG | 18 | 47109955 | G | A | 0.013 | 0.276 | 0.018 | 2.80E-56 |  |
| rs870526 | — | 2 | 20369562 | C | T | 0.479 | 0.032 | 0.004 | 7.70E-16 |  |
| rs9295128 | — | 6 | 160751531 | G | T | 0.983 | 0.196 | 0.016 | 3.40E-36 |  |
| rs9304381 | — | 18 | 47158234 | T | C | 0.818 | 0.070 | 0.005 | 7.20E-42 | Remove (confounders) |

Supplementary Table 2_5. Characteristics of instrumental variables (IVs) associated with intracranial aneurysm in the Omega-6-by-Omega-3

| **SNP** | **Nearest gene** | **Chromosome** | **Position (37/38)** | **Effect Allele** | **Other Allele** | **EAF** | **Beta** | **SE** | **P-value** | **Remove** |
| --- | --- | --- | --- | --- | --- | --- | --- | --- | --- | --- |
| rs1065853 | APOE | 19 | 45413233 | G | T | 0.919 | 0.079 | 0.008 | 1.20E-26 |  |
| rs10733306 | — | 9 | 16048248 | T | C | 0.460 | 0.023 | 0.004 | 3.20E-08 | Remove (confounders) |
| rs112875651 | — | 8 | 126506694 | A | G | 0.392 | 0.072 | 0.004 | 6.50E-65 |  |
| rs117143374 | BRWD1, PSMG1 | 21 | 40555561 | C | T | 0.142 | 0.034 | 0.006 | 6.00E-09 |  |
| rs1260326 | GCKR | 2 | 27730940 | C | T | 0.604 | 0.065 | 0.004 | 4.20E-55 | Remove (confounders) |
| rs139974673 | CATSPER2P1 | 15 | 44027885 | T | C | 0.974 | 0.117 | 0.013 | 9.10E-21 |  |
| rs149820547 | SCGB2A1 | 11 | 61983775 | G | T | 0.042 | 0.065 | 0.010 | 1.20E-10 | Remove (FADS2 locus)* |
| rs1560390 | — | 15 | 58580781 | C | T | 0.220 | 0.035 | 0.005 | 2.80E-13 | Remove (confounders) |
| rs1564348 | SLC22A1 | 6 | 160578860 | C | T | 0.170 | 0.030 | 0.005 | 1.40E-08 | Remove (confounders) |
| rs16940904 | KANSL1 | 17 | 44186063 | T | C | 0.227 | 0.041 | 0.005 | 2.30E-18 | Remove (radial MR) |
| rs174564 | FADS2 | 11 | 61588305 | G | A | 0.347 | 0.371 | 0.004 | 1.00E-200 | Remove (FADS2)* |
| rs261291 | — | 15 | 58680178 | T | C | 0.644 | 0.089 | 0.004 | 9.90E-99 | Remove (confounders) |
| rs4000713 | MIR148A | 7 | 25990597 | A | G | 0.295 | 0.031 | 0.004 | 6.10E-13 |  |
| rs55891451 | CYP2C9 | 10 | 96728169 | A | C | 0.798 | 0.036 | 0.005 | 1.20E-12 |  |
| rs58542926 | TM6SF2 | 19 | 19379549 | T | C | 0.074 | 0.143 | 0.008 | 3.40E-79 | Remove (confounders) |
| rs62466318 | — | 7 | 73042085 | T | C | 0.204 | 0.061 | 0.005 | 4.40E-33 | Remove (radial MR) |
| rs638714 | USP1 | 1 | 62906489 | T | G | 0.346 | 0.044 | 0.004 | 3.70E-25 |  |
| rs6698680 | RER1 | 1 | 2329661 | A | G | 0.538 | 0.026 | 0.004 | 2.10E-11 |  |
| rs673335 | — | 11 | 75450576 | C | T | 0.160 | 0.060 | 0.006 | 7.40E-28 |  |
| rs7222755 | TRIM65 | 17 | 73888423 | G | A | 0.291 | 0.024 | 0.005 | 4.10E-08 |  |
| rs9947684 | — | 18 | 47166694 | A | G | 0.346 | 0.028 | 0.004 | 5.70E-12 | Remove (confounders) |

*Sensitivity analysis after the removal of the FADS2 and its SNPs within 500 kb of the FADS2 locus.

Supplementary Table 3_1. Characteristics of instrumental variables (IVs) associated with subarachnoid hemorrhage in the Omega-3

| **SNP** | **Nearest gene** | **Chromosome** | **Position (37/38)** | **Effect Allele** | **Other Allele** | **EAF** | **Beta** | **SE** | **P-value** | **Remove** |
| --- | --- | --- | --- | --- | --- | --- | --- | --- | --- | --- |
| rs10455872 | LPA | 6 | 161010118 | A | G | 0.921 | 0.063 | 0.008 | 2.80E-17 | Remove (confounders) |
| rs11242109 | LOC553103, SLC22A4 | 5 | 131677047 | T | G | 0.479 | 0.024 | 0.004 | 2.40E-09 | Remove (radial MR) |
| rs11563251 | UGT1A1, UGT1A3, UGT1A4, UGT1A5, UGT1A6,  UGT1A7, UGT1A8, UGT1A9, UGT1A10 | 2 | 234679384 | T | C | 0.111 | 0.035 | 0.006 | 3.20E-08 |  |
| rs1167998 | DOCK7 | 1 | 62931632 | A | C | 0.645 | 0.071 | 0.004 | 3.60E-66 | Remove (confounders) |
| rs117733303 | LPAL2 | 6 | 160922870 | A | G | 0.981 | 0.116 | 0.015 | 1.40E-15 | Remove (confounders) |
| rs1260326 | GCKR | 2 | 27730940 | T | C | 0.396 | 0.082 | 0.004 | 8.40E-88 | Remove (confounders) |
| rs13424225 | — | 2 | 241214158 | T | G | 0.450 | 0.022 | 0.004 | 2.20E-08 |  |
| rs139974673 | CATSPER2P1 | 15 | 44027885 | C | T | 0.026 | 0.118 | 0.013 | 2.30E-21 |  |
| rs1672811 | MPV17L | 16 | 15501099 | C | T | 0.748 | 0.025 | 0.005 | 3.00E-08 |  |
| rs174564 | FADS2 | 11 | 61588305 | A | G | 0.653 | 0.337 | 0.004 | 1.00E-200 | Remove (FADS2)* |
| rs261290 | — | 15 | 58678720 | T | C | 0.345 | 0.114 | 0.004 | 3.90E-161 | Remove (confounders) |
| rs35135293 | — | 2 | 20363666 | C | T | 0.483 | 0.021 | 0.004 | 3.90E-08 |  |
| rs4000713 | MIR148A | 7 | 25990597 | G | A | 0.705 | 0.029 | 0.004 | 1.00E-11 | Remove (radial MR) |
| rs55891451 | CYP2C9 | 10 | 96728169 | C | A | 0.202 | 0.034 | 0.005 | 4.60E-12 |  |
| rs58542926 | TM6SF2 | 19 | 19379549 | C | T | 0.926 | 0.172 | 0.008 | 1.40E-113 | Remove (confounders) |
| rs6129624 | — | 20 | 39167592 | G | A | 0.665 | 0.026 | 0.004 | 5.10E-10 |  |
| rs62466318 | — | 7 | 73042085 | C | T | 0.796 | 0.072 | 0.005 | 1.20E-45 | Remove (confounders) |
| rs629301 | CELSR2 | 1 | 109818306 | T | G | 0.778 | 0.038 | 0.005 | 1.30E-14 | Remove (confounders) |
| rs633695 | LIPC, LOC101928694 | 15 | 58725839 | G | A | 0.292 | 0.084 | 0.004 | 9.10E-80 | Remove (confounders) |
| rs6601924 | AKR1C4 | 10 | 5247302 | C | T | 0.846 | 0.035 | 0.006 | 8.50E-10 |  |
| rs673335 | — | 11 | 75450576 | T | C | 0.840 | 0.067 | 0.006 | 1.10E-34 |  |
| rs6882345 | — | 5 | 156397673 | A | G | 0.633 | 0.029 | 0.004 | 1.90E-13 |  |
| rs737338 | C19orf80, DOCK6 | 19 | 11347657 | C | T | 0.965 | 0.073 | 0.011 | 3.50E-11 |  |
| rs77960347 | LIPG | 18 | 47109955 | G | A | 0.013 | 0.162 | 0.018 | 7.20E-22 |  |
| rs7819706 | — | 8 | 19844415 | A | G | 0.882 | 0.040 | 0.006 | 1.80E-10 | Remove (confounders) |
| rs7924036 | JMJD1C | 10 | 65191645 | T | G | 0.504 | 0.023 | 0.004 | 5.50E-10 | Remove (confounders) |
| rs7970695 | HNF1A | 12 | 121423376 | G | A | 0.379 | 0.025 | 0.004 | 1.20E-10 | Remove (confounders) |
| rs9304381 | — | 18 | 47158234 | T | C | 0.818 | 0.053 | 0.005 | 5.20E-24 | Remove (confounders) |

*Sensitivity analysis after the removal of the FADS2 and its SNPs within 500 kb of the FADS2 locus.

Supplementary Table 3_2. Characteristics of instrumental variables (IVs) associated with subarachnoid hemorrhage in the Omega-3-pct

| **SNP** | **Nearest gene** | **Chromosome** | **Position (37/38)** | **Effect Allele** | **Other Allele** | **EAF** | **Beta** | **SE** | **P-value** | **Remove** |
| --- | --- | --- | --- | --- | --- | --- | --- | --- | --- | --- |
| rs1260326 | GCKR | 2 | 27730940 | T | C | 0.396 | 0.038 | 0.004 | 2.30E-19 | Remove (confounders) |
| rs139974673 | CATSPER2P1 | 15 | 44027885 | C | T | 0.026 | 0.084 | 0.013 | 1.30E-11 |  |
| rs1560390 | — | 15 | 58580781 | T | C | 0.780 | 0.032 | 0.005 | 6.90E-12 | Remove (confounders) |
| rs174564 | FADS2 | 11 | 61588305 | A | G | 0.653 | 0.392 | 0.004 | 1.00E-200 | Remove (FADS2)* |
| rs190921611 | — | 19 | 45441475 | A | G | 0.311 | 0.032 | 0.005 | 2.00E-11 |  |
| rs261291 | — | 15 | 58680178 | C | T | 0.356 | 0.081 | 0.004 | 1.80E-83 | Remove (confounders) |
| rs272888 | LOC553103, SLC22A4 | 5 | 131665423 | C | T | 0.707 | 0.029 | 0.004 | 1.80E-11 | Remove (radial MR) |
| rs4000713 | MIR148A | 7 | 25990597 | G | A | 0.705 | 0.027 | 0.004 | 1.60E-10 | Remove (radial MR) |
| rs58542926 | TM6SF2 | 19 | 19379549 | C | T | 0.926 | 0.131 | 0.008 | 1.20E-67 | Remove (confounders) |
| rs62466318 | — | 7 | 73042085 | C | T | 0.796 | 0.041 | 0.005 | 3.10E-16 |  |
| rs638714 | USP1 | 1 | 62906489 | G | T | 0.654 | 0.033 | 0.004 | 4.90E-15 |  |
| rs72654473 | APOE | 19 | 45414399 | A | C | 0.108 | 0.037 | 0.007 | 6.10E-09 |  |
| rs7444298 | LOC102546226, TMEM161B-AS1 | 5 | 87730027 | G | A | 0.237 | 0.027 | 0.005 | 2.40E-08 | Remove (confounders) |
| rs7924036 | JMJD1C | 10 | 65191645 | T | G | 0.504 | 0.038 | 0.004 | 4.00E-23 | Remove (confounders) |
| rs9563335 | — | 13 | 56069705 | A | G | 0.138 | 0.044 | 0.008 | 5.50E-09 |  |
| rs9947684 | — | 18 | 47166694 | G | A | 0.654 | 0.029 | 0.004 | 2.80E-12 | Remove (confounders) |

*Sensitivity analysis after the removal of the FADS2 and its SNPs within 500 kb of the FADS2 locus.

Supplementary Table 3_3. Characteristics of instrumental variables (IVs) associated with subarachnoid hemorrhage in the DHA

| **SNP** | **Nearest gene** | **Chromosome** | **Position (37/38)** | **Effect Allele** | **Other Allele** | **EAF** | **Beta** | **SE** | **P-value** | **Remove** |
| --- | --- | --- | --- | --- | --- | --- | --- | --- | --- | --- |
| rs11122450 | GALNT2 | 1 | 230301811 | G | T | 0.611 | 0.023 | 0.004 | 3.40E-08 | Remove (confounders) |
| rs1260326 | GCKR | 2 | 27730940 | T | C | 0.396 | 0.047 | 0.004 | 8.70E-30 | Remove (confounders) |
| rs13424225 | — | 2 | 241214158 | T | G | 0.450 | 0.022 | 0.004 | 1.10E-08 |  |
| rs139974673 | CATSPER2P1 | 15 | 44027885 | C | T | 0.026 | 0.071 | 0.013 | 8.80E-09 |  |
| rs149820547 | SCGB2A1 | 11 | 61983775 | T | G | 0.958 | 0.054 | 0.010 | 4.80E-08 | Remove (FADS2 locus)* |
| rs1560390 | — | 15 | 58580781 | T | C | 0.780 | 0.050 | 0.005 | 2.50E-26 | Remove (confounders) |
| rs174564 | FADS2 | 11 | 61588305 | A | G | 0.653 | 0.290 | 0.004 | 1.00E-200 | Remove (FADS2 )* |
| rs2278426 | C19orf80, DOCK6 | 19 | 11350488 | C | T | 0.965 | 0.060 | 0.011 | 3.70E-08 | Remove (confounders) |
| rs261291 | — | 15 | 58680178 | C | T | 0.356 | 0.115 | 0.004 | 5.40E-172 | Remove (confounders) |
| rs273912 | LOC553103, SLC22A4 | 5 | 131661349 | T | G | 0.707 | 0.026 | 0.004 | 6.60E-10 | Remove (radial MR) |
| rs2807967 | DNAJC1 | 10 | 22251380 | C | T | 0.718 | 0.025 | 0.004 | 1.20E-08 |  |
| rs325 | LPL | 8 | 19819328 | C | T | 0.100 | 0.039 | 0.007 | 3.80E-09 | Remove (confounders) |
| rs3764261 | CETP | 16 | 56993324 | A | C | 0.324 | 0.039 | 0.004 | 1.90E-21 | Remove (confounders) |
| rs55891451 | CYP2C9 | 10 | 96728169 | C | A | 0.202 | 0.032 | 0.005 | 5.60E-11 |  |
| rs58542926 | TM6SF2 | 19 | 19379549 | C | T | 0.926 | 0.119 | 0.008 | 1.40E-57 | Remove (confounders) |
| rs629301 | CELSR2 | 1 | 109818306 | T | G | 0.778 | 0.044 | 0.005 | 1.40E-19 | Remove (confounders) |
| rs638714 | USP1 | 1 | 62906489 | G | T | 0.654 | 0.038 | 0.004 | 5.40E-20 | Remove (confounders) |
| rs673335 | — | 11 | 75450576 | T | C | 0.840 | 0.055 | 0.005 | 8.10E-25 |  |
| rs73045691 | — | 19 | 45440529 | A | G | 0.304 | 0.029 | 0.005 | 3.60E-10 |  |
| rs7412 | APOE | 19 | 45412079 | C | T | 0.919 | 0.077 | 0.007 | 5.40E-26 | Remove (confounders) |
| rs77960347 | LIPG | 18 | 47109955 | G | A | 0.013 | 0.142 | 0.017 | 2.50E-17 |  |
| rs7924036 | JMJD1C | 10 | 65191645 | T | G | 0.504 | 0.036 | 0.004 | 4.80E-21 | Remove (confounders) |
| rs9304381 | — | 18 | 47158234 | T | C | 0.818 | 0.050 | 0.005 | 5.80E-23 | Remove (confounders) |

*Sensitivity analysis after the removal of the FADS2 and its SNPs within 500 kb of the FADS2 locus.

Supplementary Table 3_4. Characteristics of instrumental variables (IVs) associated with subarachnoid hemorrhage in the Omega-6

| **SNP** | **Nearest gene** | **Chromosome** | **Position (37/38)** | **Effect Allele** | **Other Allele** | **EAF** | **Beta** | **SE** | **P-value** | **Remove** |
| --- | --- | --- | --- | --- | --- | --- | --- | --- | --- | --- |
| rs1002687 | DOCK7 | 1 | 62963737 | A | G | 0.645 | 0.091 | 0.004 | 1.00E-107 |  |
| rs1065853 | APOC1, APOE, TOMM40 | 19 | 45413233 | G | T | 0.919 | 0.199 | 0.007 | 3.60E-160 |  |
| rs11789603 | ABCA1 | 9 | 107647019 | T | C | 0.109 | 0.048 | 0.006 | 9.70E-14 | Remove (confounders) |
| rs1260326 | GCKR | 2 | 27730940 | T | C | 0.396 | 0.064 | 0.004 | 3.90E-55 | Remove (confounders) |
| rs12740374 | CELSR2, PSRC1 | 1 | 109817590 | G | T | 0.779 | 0.057 | 0.005 | 1.50E-32 | Remove (confounders) |
| rs142158911 | LDLR | 19 | 11190534 | G | A | 0.883 | 0.094 | 0.006 | 5.20E-52 |  |
| rs1800961 | HNF4A, MIR3646 | 20 | 43042364 | C | T | 0.970 | 0.074 | 0.012 | 3.30E-10 | Remove (confounders) |
| rs183130 | CETP | 16 | 56991363 | T | C | 0.324 | 0.062 | 0.004 | 1.40E-48 | Remove (confounders) |
| rs2378390 | ERGIC3, FER1L4 | 20 | 34150207 | G | A | 0.859 | 0.033 | 0.006 | 3.20E-09 |  |
| rs261290 | — | 15 | 58678720 | T | C | 0.345 | 0.097 | 0.004 | 1.00E-116 | Remove (confounders) |
| rs2737245 | TRPS1 | 8 | 116658583 | G | T | 0.721 | 0.027 | 0.005 | 1.40E-09 | Remove (confounders) |
| rs2740488 | ABCA1 | 9 | 107661742 | A | C | 0.735 | 0.050 | 0.005 | 5.40E-28 | Remove (confounders) |
| rs3770586 | ABCB11 | 2 | 169828995 | C | T | 0.516 | 0.023 | 0.004 | 7.10E-09 |  |
| rs4008004 | TTC39B | 9 | 15300968 | A | C | 0.222 | 0.033 | 0.005 | 8.40E-12 |  |
| rs4299376 | ABCG5, ABCG8 | 2 | 44072576 | G | T | 0.324 | 0.035 | 0.004 | 1.10E-16 |  |
| rs4439799 | TBKBP1 | 17 | 45781599 | T | C | 0.502 | 0.022 | 0.004 | 1.30E-08 |  |
| rs55747707 | MLXIPL | 7 | 73037366 | G | A | 0.796 | 0.049 | 0.005 | 1.70E-22 |  |
| rs56322906 | C19orf80, DOCK6 | 19 | 11346155 | G | A | 0.965 | 0.100 | 0.011 | 1.20E-19 |  |
| rs5754102 | UBE2L3 | 22 | 21916272 | C | A | 0.817 | 0.032 | 0.005 | 9.90E-10 | Remove (confounders) |
| rs58542926 | HAPLN4, SUGP1, TM6SF2 | 19 | 19379549 | C | T | 0.926 | 0.128 | 0.008 | 2.50E-65 | Remove (confounders) |
| rs633695 | LIPC, LOC101928694 | 15 | 58725839 | G | A | 0.292 | 0.073 | 0.004 | 1.30E-59 | Remove (confounders) |
| rs6471717 | — | 8 | 59377357 | G | A | 0.337 | 0.029 | 0.004 | 4.00E-12 |  |
| rs6547409 | — | 2 | 21190209 | C | T | 0.949 | 0.081 | 0.009 | 2.40E-20 | Remove (confounders) |
| rs672889 | — | 2 | 21319016 | G | T | 0.860 | 0.076 | 0.006 | 1.30E-41 |  |
| rs6882345 | TIMD4 | 5 | 156397673 | A | G | 0.633 | 0.045 | 0.004 | 1.20E-27 |  |
| rs6934962 | FRK | 6 | 116322349 | T | C | 0.400 | 0.023 | 0.004 | 2.20E-08 |  |
| rs7139079 | HNF1A, HNF1A-AS1 | 12 | 121415293 | G | A | 0.407 | 0.030 | 0.004 | 3.30E-13 | Remove (radial MR) |
| rs72997616 | DGAT2, LOC283214 | 11 | 75474195 | C | A | 0.906 | 0.052 | 0.007 | 1.60E-13 |  |
| rs75406471 | AKR1C4 | 10 | 5257647 | G | A | 0.845 | 0.031 | 0.006 | 2.70E-08 |  |
| rs77960347 | LIPG | 18 | 47109955 | G | A | 0.013 | 0.276 | 0.018 | 2.80E-56 |  |
| rs870526 | — | 2 | 20369562 | C | T | 0.479 | 0.032 | 0.004 | 7.70E-16 |  |
| rs9295128 | — | 6 | 160751531 | G | T | 0.983 | 0.196 | 0.016 | 3.40E-36 |  |
| rs9304381 | — | 18 | 47158234 | T | C | 0.818 | 0.070 | 0.005 | 7.20E-42 | Remove (confounders) |

Supplementary Table 3_5. Characteristics of instrumental variables (IVs) associated with subarachnoid hemorrhage in the Omega-6-by-Omega-3

| **SNP** | **Nearest gene** | **Chromosome** | **Position (37/38)** | **Effect Allele** | **Other Allele** | **EAF** | **Beta** | **SE** | **P-value** | **Remove** |
| --- | --- | --- | --- | --- | --- | --- | --- | --- | --- | --- |
| rs1065853 | APOE | 19 | 45413233 | G | T | 0.919 | 0.079 | 0.008 | 1.20E-26 |  |
| rs10733306 | — | 9 | 16048248 | T | C | 0.460 | 0.023 | 0.004 | 3.20E-08 | Remove (confounders) |
| rs11242109 | LOC553103, SLC22A4 | 5 | 131677047 | G | T | 0.521 | 0.024 | 0.004 | 1.40E-09 | Remove (radial MR) |
| rs1260326 | GCKR | 2 | 27730940 | C | T | 0.604 | 0.065 | 0.004 | 4.20E-55 | Remove (confounders) |
| rs13273454 | — | 8 | 19940058 | T | C | 0.471 | 0.023 | 0.004 | 5.80E-09 |  |
| rs139974673 | CATSPER2P1 | 15 | 44027885 | T | C | 0.974 | 0.117 | 0.013 | 9.10E-21 |  |
| rs149820547 | SCGB2A1 | 11 | 61983775 | G | T | 0.042 | 0.065 | 0.010 | 1.20E-10 | Remove (FADS2 locus)* |
| rs1560390 | — | 15 | 58580781 | C | T | 0.220 | 0.035 | 0.005 | 2.80E-13 | Remove (confounders) |
| rs1564348 | SLC22A1 | 6 | 160578860 | C | T | 0.170 | 0.030 | 0.005 | 1.40E-08 |  |
| rs174564 | FADS2 | 11 | 61588305 | G | A | 0.347 | 0.371 | 0.004 | 1.00E-200 | Remove (FADS2 )* |
| rs261291 | — | 15 | 58680178 | T | C | 0.644 | 0.089 | 0.004 | 9.90E-99 | Remove (confounders) |
| rs4000713 | MIR148A | 7 | 25990597 | A | G | 0.295 | 0.031 | 0.004 | 6.10E-13 | Remove (radial MR) |
| rs55891451 | CYP2C9 | 10 | 96728169 | A | C | 0.798 | 0.036 | 0.005 | 1.20E-12 |  |
| rs58542926 | TM6SF2 | 19 | 19379549 | T | C | 0.074 | 0.143 | 0.008 | 3.40E-79 | Remove (confounders) |
| rs62466318 | — | 7 | 73042085 | T | C | 0.204 | 0.061 | 0.005 | 4.40E-33 |  |
| rs638714 | USP1 | 1 | 62906489 | T | G | 0.346 | 0.044 | 0.004 | 3.70E-25 |  |
| rs673335 | — | 11 | 75450576 | C | T | 0.160 | 0.060 | 0.006 | 7.40E-28 |  |
| rs7222755 | TRIM65 | 17 | 73888423 | G | A | 0.291 | 0.024 | 0.005 | 4.10E-08 |  |
| rs9947684 | — | 18 | 47166694 | A | G | 0.346 | 0.028 | 0.004 | 5.70E-12 | Remove (confounders) |

*Sensitivity analysis after the removal of the FADS2 and its SNPs within 500 kb of the FADS2 locus.

Supplementary Table 4_1. Characteristics of instrumental variables (IVs) associated with unruptured intracranial aneurysm in the Omega-3

| **SNP** | **Nearest gene** | **Chromosome** | **Position (37/38)** | **Effect Allele** | **Other Allele** | **EAF** | **Beta** | **SE** | **P-value** | **Remove** |
| --- | --- | --- | --- | --- | --- | --- | --- | --- | --- | --- |
| rs10455872 | LPA | 6 | 161010118 | A | G | 0.921 | 0.063 | 0.008 | 2.80E-17 |  |
| rs11242109 | LOC553103, SLC22A4 | 5 | 131677047 | T | G | 0.479 | 0.024 | 0.004 | 2.40E-09 |  |
| rs1132899 | APOC2, APOC4, APOC4-APOC2 | 19 | 45448036 | C | T | 0.510 | 0.027 | 0.004 | 8.60E-11 | Remove (confounders) |
| rs11563251 | UGT1A1, UGT1A3, UGT1A4, UGT1A5, UGT1A6,  UGT1A7, UGT1A8, UGT1A9, UGT1A10 | 2 | 234679384 | T | C | 0.111 | 0.035 | 0.006 | 3.20E-08 |  |
| rs1167998 | DOCK7 | 1 | 62931632 | A | C | 0.645 | 0.071 | 0.004 | 3.60E-66 |  |
| rs11681659 | — | 2 | 136820960 | C | T | 0.284 | 0.025 | 0.004 | 2.00E-08 |  |
| rs117143374 | BRWD1, PSMG1 | 21 | 40555561 | T | C | 0.858 | 0.037 | 0.006 | 2.20E-10 | Remove (radial MR) |
| rs117733303 | LPAL2 | 6 | 160922870 | A | G | 0.981 | 0.116 | 0.015 | 1.40E-15 |  |
| rs12226389 | — | 11 | 61823630 | T | C | 0.814 | 0.051 | 0.005 | 1.10E-22 | Remove (FADS locus)* |
| rs1260326 | GCKR | 2 | 27730940 | T | C | 0.396 | 0.082 | 0.004 | 8.40E-88 | Remove (confounders) |
| rs13424225 | — | 2 | 241214158 | T | G | 0.450 | 0.022 | 0.004 | 2.20E-08 |  |
| rs139974673 | CATSPER2P1 | 15 | 44027885 | C | T | 0.026 | 0.118 | 0.013 | 2.30E-21 |  |
| rs143355652 | DAGLA | 11 | 61453822 | C | T | 0.990 | 0.154 | 0.020 | 9.40E-14 | Remove (FADS2 locus)* |
| rs1672811 | MPV17L | 16 | 15501099 | C | T | 0.748 | 0.025 | 0.005 | 3.00E-08 |  |
| rs16940904 | KANSL1 | 17 | 44186063 | C | T | 0.773 | 0.036 | 0.005 | 3.90E-14 | Remove (radial MR) |
| rs174564 | FADS2 | 11 | 61588305 | A | G | 0.653 | 0.337 | 0.004 | 1.00E-200 | Remove (FADS2)* |
| rs182611493 | MAU2 | 19 | 19458388 | A | G | 0.987 | 0.210 | 0.020 | 1.10E-27 |  |
| rs2394976 | — | 6 | 31311912 | G | T | 0.838 | 0.046 | 0.006 | 1.20E-15 | Remove (radial MR) |
| rs261290 | — | 15 | 58678720 | T | C | 0.345 | 0.114 | 0.004 | 3.90E-161 | Remove (confounders) |
| rs3018731 | PPP1R32 | 11 | 61248776 | A | G | 0.282 | 0.035 | 0.005 | 2.00E-14 | Remove (FADS2 locus)* |
| rs34663616 | — | 15 | 58569330 | A | C | 0.138 | 0.036 | 0.006 | 4.40E-10 | Remove (radial MR) |
| rs35135293 | — | 2 | 20363666 | C | T | 0.483 | 0.021 | 0.004 | 3.90E-08 |  |
| rs4000713 | MIR148A | 7 | 25990597 | G | A | 0.705 | 0.029 | 0.004 | 1.00E-11 |  |
| rs58542926 | TM6SF2 | 19 | 19379549 | C | T | 0.926 | 0.172 | 0.008 | 1.40E-113 | Remove (confounders) |
| rs6129624 | — | 20 | 39167592 | G | A | 0.665 | 0.026 | 0.004 | 5.10E-10 |  |
| rs62466318 | — | 7 | 73042085 | C | T | 0.796 | 0.072 | 0.005 | 1.20E-45 |  |
| rs629301 | CELSR2 | 1 | 109818306 | T | G | 0.778 | 0.038 | 0.005 | 1.30E-14 | Remove (confounders) |
| rs633695 | LIPC, LOC101928694 | 15 | 58725839 | G | A | 0.292 | 0.084 | 0.004 | 9.10E-80 | Remove (confounders) |
| rs6601924 | AKR1C4 | 10 | 5247302 | C | T | 0.846 | 0.035 | 0.006 | 8.50E-10 |  |
| rs6693447 | RER1 | 1 | 2330190 | G | T | 0.462 | 0.023 | 0.004 | 4.80E-09 |  |
| rs673335 | — | 11 | 75450576 | T | C | 0.840 | 0.067 | 0.006 | 1.10E-34 |  |
| rs6882345 | — | 5 | 156397673 | A | G | 0.633 | 0.029 | 0.004 | 1.90E-13 |  |
| rs73109460 | ZMIZ2 | 7 | 44785800 | G | A | 0.876 | 0.035 | 0.006 | 9.20E-10 |  |
| rs737338 | C19orf80, DOCK6 | 19 | 11347657 | C | T | 0.965 | 0.073 | 0.011 | 3.50E-11 |  |
| rs77960347 | LIPG | 18 | 47109955 | G | A | 0.013 | 0.162 | 0.018 | 7.20E-22 |  |
| rs7819706 | — | 8 | 19844415 | A | G | 0.882 | 0.040 | 0.006 | 1.80E-10 | Remove (confounders) |
| rs7924036 | JMJD1C | 10 | 65191645 | T | G | 0.504 | 0.023 | 0.004 | 5.50E-10 | Remove (confounders) |
| rs7970695 | HNF1A | 12 | 121423376 | G | A | 0.379 | 0.025 | 0.004 | 1.20E-10 |  |
| rs9304381 | — | 18 | 47158234 | T | C | 0.818 | 0.053 | 0.005 | 5.20E-24 | Remove (confounders) |
| rs9987289 | LOC157273 | 8 | 9183358 | G | A | 0.909 | 0.057 | 0.007 | 3.20E-16 | Remove (confounders) |

*Sensitivity analysis after the removal of the FADS2 and its SNPs within 500 kb of the FADS2 locus.

Supplementary Table 4_2. Characteristics of instrumental variables (IVs) associated with unruptured intracranial aneurysm in the Omega-3-pct

| **SNP** | **Nearest gene** | **Chromosome** | **Position (37/38)** | **Effect Allele** | **Other Allele** | **EAF** | **Beta** | **SE** | **P-value** | **Remove** |
| --- | --- | --- | --- | --- | --- | --- | --- | --- | --- | --- |
| rs11632618 | LIPC, LOC101928694 | 15 | 58724706 | A | G | 0.070 | 0.078 | 0.008 | 1.10E-22 | Remove (confounders) |
| rs12226389 | — | 11 | 61823630 | T | C | 0.814 | 0.062 | 0.005 | 2.20E-32 | Remove (FADS2 locus)* |
| rs1260326 | GCKR | 2 | 27730940 | T | C | 0.396 | 0.038 | 0.004 | 2.30E-19 | Remove (confounders) |
| rs139974673 | CATSPER2P1 | 15 | 44027885 | C | T | 0.026 | 0.084 | 0.013 | 1.30E-11 |  |
| rs143355652 | DAGLA | 11 | 61453822 | C | T | 0.990 | 0.179 | 0.020 | 1.20E-18 | Remove (FADS2 locus)* |
| rs145786300 | RPLP0P2 | 11 | 61406089 | G | A | 0.988 | 0.187 | 0.019 | 1.30E-22 | Remove (FADS2 locus)* |
| rs149402055 | CPT1A | 11 | 68525539 | C | T | 0.980 | 0.173 | 0.015 | 9.20E-30 |  |
| rs1560390 | — | 15 | 58580781 | T | C | 0.780 | 0.032 | 0.005 | 6.90E-12 | Remove (confounders) |
| rs16940904 | KANSL1 | 17 | 44186063 | C | T | 0.773 | 0.042 | 0.005 | 2.40E-19 | Remove (radial MR) |
| rs174564 | FADS2 | 11 | 61588305 | A | G | 0.653 | 0.392 | 0.004 | 1.00E-200 | Remove (FADS2)* |
| rs182611493 | MAU2 | 19 | 19458388 | A | G | 0.987 | 0.172 | 0.020 | 8.90E-19 |  |
| rs191623731 | — | 11 | 61844663 | G | T | 0.016 | 0.118 | 0.016 | 9.80E-14 | Remove (FADS2 locus)* |
| rs2011946 | — | 2 | 136817616 | C | A | 0.265 | 0.027 | 0.005 | 2.60E-09 |  |
| rs2232143 | VPS37C | 11 | 60899701 | C | T | 0.022 | 0.108 | 0.014 | 1.30E-13 |  |
| rs2394976 | — | 6 | 31311912 | G | T | 0.838 | 0.031 | 0.006 | 5.40E-09 | Remove (radial MR) |
| rs261291 | — | 15 | 58680178 | C | T | 0.356 | 0.081 | 0.004 | 1.80E-83 | Remove (confounders) |
| rs272888 | LOC553103, SLC22A4 | 5 | 131665423 | C | T | 0.707 | 0.029 | 0.004 | 1.80E-11 | Remove (radial MR) |
| rs4000713 | MIR148A | 7 | 25990597 | G | A | 0.705 | 0.027 | 0.004 | 1.60E-10 |  |
| rs58542926 | TM6SF2 | 19 | 19379549 | C | T | 0.926 | 0.131 | 0.008 | 1.20E-67 | Remove (confounders) |
| rs62466318 | — | 7 | 73042085 | C | T | 0.796 | 0.041 | 0.005 | 3.10E-16 |  |
| rs638714 | USP1 | 1 | 62906489 | G | T | 0.654 | 0.033 | 0.004 | 4.90E-15 |  |
| rs6693447 | RER1 | 1 | 2330190 | G | T | 0.462 | 0.026 | 0.004 | 8.30E-12 |  |
| rs72654473 | APOE | 19 | 45414399 | A | C | 0.108 | 0.037 | 0.007 | 6.10E-09 |  |
| rs73109460 | ZMIZ2 | 7 | 44785800 | G | A | 0.876 | 0.032 | 0.006 | 7.80E-09 |  |
| rs7444298 | LOC102546226, TMEM161B-AS1 | 5 | 87730027 | G | A | 0.237 | 0.027 | 0.005 | 2.40E-08 | Remove (confounders) |
| rs7924036 | JMJD1C | 10 | 65191645 | T | G | 0.504 | 0.038 | 0.004 | 4.00E-23 | Remove (confounders) |
| rs8074191 | PEMT | 17 | 17407191 | T | C | 0.244 | 0.027 | 0.005 | 2.30E-08 |  |
| rs9563335 | — | 13 | 56069705 | A | G | 0.138 | 0.044 | 0.008 | 5.50E-09 |  |
| rs9947684 | — | 18 | 47166694 | G | A | 0.654 | 0.029 | 0.004 | 2.80E-12 | Remove (confounders) |

*Sensitivity analysis after the removal of the FADS2 and its SNPs within 500 kb of the FADS2 locus.

Supplementary Table 4_3. Characteristics of instrumental variables (IVs) associated with unruptured intracranial aneurysm in the DHA

| **SNP** | **Nearest gene** | **Chromosome** | **Position (37/38)** | **Effect Allele** | **Other Allele** | **EAF** | **Beta** | **SE** | **P-value** | **Remove** |
| --- | --- | --- | --- | --- | --- | --- | --- | --- | --- | --- |
| rs11122450 | GALNT2 | 1 | 230301811 | G | T | 0.611 | 0.023 | 0.004 | 3.40E-08 | Remove (confounders) |
| rs11681659 | — | 2 | 136820960 | C | T | 0.284 | 0.029 | 0.004 | 5.30E-12 |  |
| rs12226389 | — | 11 | 61823630 | T | C | 0.814 | 0.044 | 0.005 | 3.80E-18 | Remove (FADS2 locus)* |
| rs1260326 | GCKR | 2 | 27730940 | T | C | 0.396 | 0.047 | 0.004 | 8.70E-30 | Remove (confounders) |
| rs12914626 | LIPC, LOC101928694 | 15 | 58738423 | C | T | 0.298 | 0.061 | 0.004 | 7.50E-45 |  |
| rs13424225 | — | 2 | 241214158 | T | G | 0.450 | 0.022 | 0.004 | 1.10E-08 |  |
| rs139974673 | CATSPER2P1 | 15 | 44027885 | C | T | 0.026 | 0.071 | 0.013 | 8.80E-09 |  |
| rs143355652 | DAGLA | 11 | 61453822 | C | T | 0.990 | 0.151 | 0.020 | 6.60E-14 | Remove (FADS2 locus)* |
| rs145659493 | — | 11 | 61850279 | A | C | 0.016 | 0.095 | 0.016 | 1.70E-09 | Remove (FADS2 locus)* |
| rs145786300 | RPLP0P2 | 11 | 61406089 | G | A | 0.988 | 0.132 | 0.019 | 4.00E-12 | Remove (FADS2 locus)* |
| rs149820547 | SCGB2A1 | 11 | 61983775 | T | G | 0.958 | 0.054 | 0.010 | 4.80E-08 | Remove (FADS2 locus)* |
| rs1560390 | — | 15 | 58580781 | T | C | 0.780 | 0.050 | 0.005 | 2.50E-26 | Remove (confounders) |
| rs16940904 | KANSL1 | 17 | 44186063 | C | T | 0.773 | 0.040 | 0.005 | 2.10E-17 | Remove (radial MR) |
| rs174564 | FADS2 | 11 | 61588305 | A | G | 0.653 | 0.290 | 0.004 | 1.00E-200 | Remove (FADS2)* |
| rs182611493 | MAU2 | 19 | 19458388 | A | G | 0.987 | 0.157 | 0.019 | 1.60E-16 |  |
| rs2232143 | VPS37C | 11 | 60899701 | C | T | 0.022 | 0.085 | 0.014 | 6.60E-09 |  |
| rs2278426 | C19orf80, DOCK6 | 19 | 11350488 | C | T | 0.965 | 0.060 | 0.011 | 3.70E-08 | Remove (confounders) |
| rs2394976 | — | 6 | 31311912 | G | T | 0.838 | 0.034 | 0.005 | 5.60E-11 | Remove (radial MR) |
| rs261291 | — | 15 | 58680178 | C | T | 0.356 | 0.115 | 0.004 | 5.40E-172 | Remove (confounders) |
| rs273912 | LOC553103, SLC22A4 | 5 | 131661349 | T | G | 0.707 | 0.026 | 0.004 | 6.60E-10 | Remove (radial MR) |
| rs2807967 | DNAJC1 | 10 | 22251380 | C | T | 0.718 | 0.025 | 0.004 | 1.20E-08 |  |
| rs325 | LPL | 8 | 19819328 | C | T | 0.100 | 0.039 | 0.007 | 3.80E-09 | Remove (confounders) |
| rs35177659 | — | 7 | 1218538 | T | C | 0.522 | 0.023 | 0.004 | 2.70E-08 |  |
| rs3764261 | CETP | 16 | 56993324 | A | C | 0.324 | 0.039 | 0.004 | 1.90E-21 | Remove (confounders) |
| rs525028 | APOA1, APOC3 | 11 | 116705516 | G | A | 0.291 | 0.030 | 0.004 | 2.70E-12 |  |
| rs58542926 | TM6SF2 | 19 | 19379549 | C | T | 0.926 | 0.119 | 0.008 | 1.40E-57 | Remove (confounders) |
| rs629301 | CELSR2 | 1 | 109818306 | T | G | 0.778 | 0.044 | 0.005 | 1.40E-19 | Remove (confounders) |
| rs638714 | USP1 | 1 | 62906489 | G | T | 0.654 | 0.038 | 0.004 | 5.40E-20 |  |
| rs673335 | — | 11 | 75450576 | T | C | 0.840 | 0.055 | 0.005 | 8.10E-25 |  |
| rs6931604 | — | 6 | 98578215 | T | C | 0.600 | 0.023 | 0.004 | 2.70E-08 |  |
| rs72836561 | CD300LG | 17 | 41926126 | C | T | 0.969 | 0.063 | 0.011 | 3.60E-08 | Remove (confounders) |
| rs73045691 | — | 19 | 45440529 | A | G | 0.304 | 0.029 | 0.005 | 3.60E-10 |  |
| rs73109460 | ZMIZ2 | 7 | 44785800 | G | A | 0.876 | 0.037 | 0.006 | 7.30E-11 |  |
| rs7412 | APOE | 19 | 45412079 | C | T | 0.919 | 0.077 | 0.007 | 5.40E-26 | Remove (confounders) |
| rs77960347 | LIPG | 18 | 47109955 | G | A | 0.013 | 0.142 | 0.017 | 2.50E-17 |  |
| rs7924036 | JMJD1C | 10 | 65191645 | T | G | 0.504 | 0.036 | 0.004 | 4.80E-21 | Remove (confounders) |
| rs9304381 | — | 18 | 47158234 | T | C | 0.818 | 0.050 | 0.005 | 5.80E-23 | Remove (confounders) |
| rs9987289 | LOC157273 | 8 | 9183358 | G | A | 0.909 | 0.058 | 0.007 | 1.30E-17 | Remove (confounders) |

*Sensitivity analysis after the removal of the FADS2 and its SNPs within 500 kb of the FADS2 locus.

Supplementary Table 4_4. Characteristics of instrumental variables (IVs) associated with unruptured intracranial aneurysm in the Omega-6

| **SNP** | **Nearest gene** | **Chromosome** | **Position (37/38)** | **Effect Allele** | **Other Allele** | **EAF** | **Beta** | **SE** | **P-value** | **Remove** |
| --- | --- | --- | --- | --- | --- | --- | --- | --- | --- | --- |
| rs1002687 | DOCK7 | 1 | 62963737 | A | G | 0.645 | 0.091 | 0.004 | 1.00E-107 |  |
| rs1081105 | APOE | 19 | 45412955 | C | A | 0.028 | 0.119 | 0.012 | 1.80E-22 |  |
| rs114863007 | SNRPC | 6 | 34729158 | G | A | 0.905 | 0.046 | 0.007 | 7.30E-12 | Remove (confounders) |
| rs11789603 | ABCA1 | 9 | 107647019 | T | C | 0.109 | 0.048 | 0.006 | 9.70E-14 | Remove (confounders) |
| rs1260326 | GCKR | 2 | 27730940 | T | C | 0.396 | 0.064 | 0.004 | 3.90E-55 | Remove (confounders) |
| rs12740374 | CELSR2 | 1 | 109817590 | G | T | 0.779 | 0.057 | 0.005 | 1.50E-32 | Remove (confounders) |
| rs13108218 | HGFAC, RGS12 | 4 | 3443931 | A | G | 0.385 | 0.035 | 0.004 | 3.60E-18 |  |
| rs141469619 | SIK3 | 11 | 116714293 | G | A | 0.010 | 0.111 | 0.021 | 1.40E-08 |  |
| rs142158911 | — | 19 | 11190534 | G | A | 0.883 | 0.094 | 0.006 | 5.20E-52 |  |
| rs1461729 | LOC157273 | 8 | 9187242 | G | A | 0.899 | 0.084 | 0.007 | 2.80E-36 | Remove (confounders) |
| rs1800961 | HNF4A | 20 | 43042364 | C | T | 0.970 | 0.074 | 0.012 | 3.30E-10 | Remove (confounders) |
| rs183130 | — | 16 | 56991363 | T | C | 0.324 | 0.062 | 0.004 | 1.40E-48 | Remove (confounders) |
| rs2378390 | FER1L4 | 20 | 34150207 | G | A | 0.859 | 0.033 | 0.006 | 3.20E-09 |  |
| rs261290 | — | 15 | 58678720 | T | C | 0.345 | 0.097 | 0.004 | 1.00E-116 | Remove (confounders) |
| rs2737245 | TRPS1 | 8 | 116658583 | G | T | 0.721 | 0.027 | 0.005 | 1.40E-09 | Remove (confounders) |
| rs2740488 | ABCA1 | 9 | 107661742 | A | C | 0.735 | 0.050 | 0.005 | 5.40E-28 | Remove (confounders) |
| rs28383314 | — | 6 | 32587213 | C | T | 0.623 | 0.039 | 0.004 | 1.70E-18 |  |
| rs3734854 | C6orf15 | 6 | 31078836 | A | G | 0.065 | 0.048 | 0.008 | 5.90E-11 |  |
| rs3770586 | ABCB11 | 2 | 169828995 | C | T | 0.516 | 0.023 | 0.004 | 7.10E-09 |  |
| rs4299376 | ABCG8 | 2 | 44072576 | G | T | 0.324 | 0.035 | 0.004 | 1.10E-16 |  |
| rs4439799 | TBKBP1 | 17 | 45781599 | T | C | 0.502 | 0.022 | 0.004 | 1.30E-08 |  |
| rs534417 | ASAP3 | 1 | 23784965 | G | A | 0.875 | 0.039 | 0.006 | 9.30E-11 |  |
| rs55747707 | MLXIPL | 7 | 73037366 | G | A | 0.796 | 0.049 | 0.005 | 1.70E-22 |  |
| rs56322906 | DOCK6 | 19 | 11346155 | G | A | 0.965 | 0.100 | 0.011 | 1.20E-19 |  |
| rs58542926 | TM6SF2 | 19 | 19379549 | C | T | 0.926 | 0.128 | 0.008 | 2.50E-65 | Remove (confounders) |
| rs633695 | LIPC, LOC101928694 | 15 | 58725839 | G | A | 0.292 | 0.073 | 0.004 | 1.30E-59 | Remove (confounders) |
| rs6471717 | — | 8 | 59377357 | G | A | 0.337 | 0.029 | 0.004 | 4.00E-12 |  |
| rs6547409 | — | 2 | 21190209 | C | T | 0.949 | 0.081 | 0.009 | 2.40E-20 | Remove (confounders) |
| rs6602911 | GAS6, GAS6-AS1 | 13 | 114547372 | T | C | 0.360 | 0.026 | 0.004 | 1.30E-09 |  |
| rs672889 | — | 2 | 21319016 | G | T | 0.860 | 0.076 | 0.006 | 1.30E-41 |  |
| rs6882345 | — | 5 | 156397673 | A | G | 0.633 | 0.045 | 0.004 | 1.20E-27 |  |
| rs6934962 | FRK | 6 | 116322349 | T | C | 0.400 | 0.023 | 0.004 | 2.20E-08 |  |
| rs6938647 | LPA | 6 | 160986915 | A | C | 0.218 | 0.048 | 0.005 | 1.90E-23 | Remove (radial MR) |
| rs7139079 | HNF1A | 12 | 121415293 | G | A | 0.407 | 0.030 | 0.004 | 3.30E-13 |  |
| rs72997616 | LOC283214 | 11 | 75474195 | C | A | 0.906 | 0.052 | 0.007 | 1.60E-13 |  |
| rs75406471 | AKR1C4 | 10 | 5257647 | G | A | 0.845 | 0.031 | 0.006 | 2.70E-08 |  |
| rs7750288 | IGF2R | 6 | 160400147 | G | A | 0.285 | 0.025 | 0.004 | 1.30E-08 |  |
| rs77960347 | LIPG | 18 | 47109955 | G | A | 0.013 | 0.276 | 0.018 | 2.80E-56 |  |
| rs79429216 | APOC4, APOC4-APOC2 | 19 | 45445517 | A | G | 0.013 | 0.151 | 0.018 | 1.30E-17 |  |
| rs870526 | — | 2 | 20369562 | C | T | 0.479 | 0.032 | 0.004 | 7.70E-16 |  |
| rs9295128 | — | 6 | 160751531 | G | T | 0.983 | 0.196 | 0.016 | 3.40E-36 |  |
| rs9304381 | — | 18 | 47158234 | T | C | 0.818 | 0.070 | 0.005 | 7.20E-42 | Remove (confounders) |

Supplementary Table 4_5. Characteristics of instrumental variables (IVs) associated with unruptured intracranial aneurysm in the Omega-6-by-Omega-3

| **SNP** | **Nearest gene** | **Chromosome** | **Position (37/38)** | **Effect Allele** | **Other Allele** | **EAF** | **Beta** | **SE** | **P-value** | **Remove** |
| --- | --- | --- | --- | --- | --- | --- | --- | --- | --- | --- |
| rs10733306 | — | 9 | 16048248 | T | C | 0.460 | 0.023 | 0.004 | 3.20E-08 | Remove (confounders) |
| rs11242109 | LOC553103, SLC22A4 | 5 | 131677047 | G | T | 0.521 | 0.024 | 0.004 | 1.40E-09 |  |
| rs11632618 | LIPC, LOC101928694 | 15 | 58724706 | G | A | 0.930 | 0.082 | 0.008 | 1.20E-24 | Remove (confounders) |
| rs116843064 | ANGPTL4 | 19 | 8429323 | A | G | 0.020 | 0.084 | 0.015 | 1.60E-08 | Remove (confounders) |
| rs117143374 | BRWD1, PSMG1 | 21 | 40555561 | C | T | 0.142 | 0.034 | 0.006 | 6.00E-09 | Remove (radial MR) |
| rs12226389 | — | 11 | 61823630 | C | T | 0.186 | 0.056 | 0.005 | 8.40E-27 | Remove (FADS2 locus)* |
| rs1260326 | GCKR | 2 | 27730940 | C | T | 0.604 | 0.065 | 0.004 | 4.20E-55 | Remove (confounders) |
| rs139974673 | CATSPER2P1 | 15 | 44027885 | T | C | 0.974 | 0.117 | 0.013 | 9.10E-21 |  |
| rs143355652 | DAGLA | 11 | 61453822 | T | C | 0.010 | 0.160 | 0.021 | 7.60E-15 | Remove (FADS2 locus)* |
| rs145659493 | — | 11 | 61850279 | C | A | 0.984 | 0.116 | 0.016 | 4.30E-13 | Remove (FADS2 locus)* |
| rs145786300 | RPLP0P2 | 11 | 61406089 | A | G | 0.012 | 0.174 | 0.019 | 1.10E-19 | Remove (FADS2 locus)* |
| rs149820547 | SCGB2A1 | 11 | 61983775 | G | T | 0.042 | 0.065 | 0.010 | 1.20E-10 | Remove (FADS2 locus)* |
| rs1560390 | — | 15 | 58580781 | C | T | 0.220 | 0.035 | 0.005 | 2.80E-13 | Remove (confounders) |
| rs1564348 | SLC22A1 | 6 | 160578860 | C | T | 0.170 | 0.030 | 0.005 | 1.40E-08 |  |
| rs16940904 | KANSL1 | 17 | 44186063 | T | C | 0.227 | 0.041 | 0.005 | 2.30E-18 | Remove (confounders) |
| rs174564 | FADS2 | 11 | 61588305 | G | A | 0.347 | 0.371 | 0.004 | 1.00E-200 | Remove (FADS2)* |
| rs182611493 | MAU2 | 19 | 19458388 | G | A | 0.013 | 0.187 | 0.020 | 1.10E-21 |  |
| rs2232143 | VPS37C | 11 | 60899701 | T | C | 0.978 | 0.110 | 0.015 | 9.40E-14 |  |
| rs2394976 | — | 6 | 31311912 | T | G | 0.162 | 0.034 | 0.006 | 5.80E-10 | Remove (radial MR) |
| rs261291 | — | 15 | 58680178 | T | C | 0.644 | 0.089 | 0.004 | 9.90E-99 | Remove (confounders) |
| rs4000713 | MIR148A | 7 | 25990597 | A | G | 0.295 | 0.031 | 0.004 | 6.10E-13 |  |
| rs58542926 | TM6SF2 | 19 | 19379549 | T | C | 0.074 | 0.143 | 0.008 | 3.40E-79 | Remove (confounders) |
| rs62466318 | — | 7 | 73042085 | T | C | 0.204 | 0.061 | 0.005 | 4.40E-33 |  |
| rs638714 | USP1 | 1 | 62906489 | T | G | 0.346 | 0.044 | 0.004 | 3.70E-25 |  |
| rs6698680 | RER1 | 1 | 2329661 | A | G | 0.538 | 0.026 | 0.004 | 2.10E-11 |  |
| rs673335 | — | 11 | 75450576 | C | T | 0.160 | 0.060 | 0.006 | 7.40E-28 |  |
| rs7222755 | TRIM65 | 17 | 73888423 | G | A | 0.291 | 0.024 | 0.005 | 4.10E-08 |  |
| rs73109460 | ZMIZ2 | 7 | 44785800 | A | G | 0.124 | 0.034 | 0.006 | 2.20E-09 |  |
| rs8074191 | PEMT | 17 | 17407191 | C | T | 0.756 | 0.028 | 0.005 | 4.80E-09 |  |
| rs9947684 | — | 18 | 47166694 | A | G | 0.346 | 0.028 | 0.004 | 5.70E-12 | Remove (confounders) |

*Sensitivity analysis after the removal of the FADS2 and its SNPs within 500 kb of the FADS2 locus.

Supplementary Table 5. Sensitivity analysis of forward causal effect between fatty acid and the risk of aneurysm.

| **Exposure** | **Outcome** | **Heterogeneity test**  **(MR-Egger/IVW)** | | | **Horizontal pleiotropy** | | | **MR-PRESSO P value** | **Steiger test** | | **R2(%)** | **F-statistics** | **Power**  **(%)** |
| --- | --- | --- | --- | --- | --- | --- | --- | --- | --- | --- | --- | --- | --- |
|  |  | **Q** | **Q df** | **P value** | **Egger intercept** | **SE** | **P value** |  | **correct causal**  **direction** | **Pvalue** |  |  |  |
| Omega-3 | IA | 11.896 | 13 | 0.536 | -0.003 | 0.011 | 0.754 | 0.601 | TRUE | P<0.001 | 5.9 | 481.388 | 97.017 |
|  |  | 11.999 | 14 | 0.606 |  |  |  |  |  |  |  |  |  |
|  | aSAH | 10.432 | 12 | 0.578 | -0.012 | 0.012 | 0.357 | 0.361 | TRUE | P<0.001 | 5.7 | 534.685 | 99.225 |
|  |  | 11.349 | 13 | 0.582 |  |  |  |  |  |  |  |  |  |
|  | uIA | 16.100 | 24 | 0.884 | -0.010 | 0.011 | 0.360 | 0.314 | TRUE | P<0.001 | 6.7 | 309.641 | 17.446 |
|  |  | 16.973 | 25 | 0.883 |  |  |  |  |  |  |  |  |  |
| Omega-3-pct | IA | 2.876 | 6 | 0.824 | -0.027 | 0.014 | 0.108 | 0.386 | TRUE | P<0.001 | 7.5 | 1152.764 | 98.983 |
|  |  | 6.442 | 7 | 0.489 |  |  |  |  |  |  |  |  |  |
|  | aSAH | 5.127 | 5 | 0.401 | -0.014 | 0.018 | 0.464 | 0.435 | TRUE | P<0.001 | 7.1 | 1263.711 | 98.584 |
|  |  | 5.771 | 6 | 0.449 |  |  |  |  |  |  |  |  |  |
|  | uIA | 15.965 | 16 | 0.455 | -0.004 | 0.014 | 0.763 | 0.222 | TRUE | P<0.001 | 7.8 | 533.331 | 6.910 |
|  |  | 16.059 | 17 | 0.520 |  |  |  |  |  |  |  |  |  |
| DHA | IA | 3.041 | 9 | 0.963 | -0.005 | 0.013 | 0.711 | 0.396 | TRUE | P<0.001 | 4.5 | 493.251 | 98.759 |
|  |  | 3.187 | 10 | 0.977 |  |  |  |  |  |  |  |  |  |
|  | aSAH | 6.279 | 7 | 0.508 | -0.013 | 0.016 | 0.446 | 0.421 | TRUE | P<0.001 | 4.3 | 578.021 | 99.377 |
|  |  | 6.930 | 8 | 0.544 |  |  |  |  |  |  |  |  |  |
|  | uIA | 16.219 | 19 | 0.643 | -0.008 | 0.013 | 0.563 | 0.210 | TRUE | P<0.001 | 4.9 | 282.662 | 10.332 |
|  |  | 16.567 | 20 | 0.681 |  |  |  |  |  |  |  |  |  |
| Omega-6 by Omega-3 | IA | 4.257 | 10 | 0.935 | 0.020 | 0.012 | 0.119 | 0.453 | TRUE | P<0.001 | 6.9 | 709.925 | 99.962 |
|  |  | 7.164 | 11 | 0.786 |  |  |  |  |  |  |  |  |  |
|  | aSAH | 7.573 | 9 | 0.578 | 0.006 | 0.014 | 0.664 | 0.327 | TRUE | P<0.001 | 6.8 | 756.057 | 99.987 |
|  |  | 7.775 | 10 | 0.651 |  |  |  |  |  |  |  |  |  |
|  | uIA | 13.836 | 17 | 0.679 | 0.015 | 0.014 | 0.293 | 0.378 | TRUE | P<0.001 | 7.2 | 461.187 | 10.702 |
|  |  | 15.013 | 18 | 0.661 |  |  |  |  |  |  |  |  |  |
| Omega-6 | IA | 17.962 | 17 | 0.391 | 0.008 | 0.014 | 0.563 | 0.431 | TRUE | P<0.001 | 1.4 | 86.063 | 52.746 |
|  |  | 18.329 | 18 | 0.434 |  |  |  |  |  |  |  |  |  |
|  | aSAH | 16.418 | 17 | 0.494 | 0.004 | 0.014 | 0.764 | 0.342 | TRUE | P<0.001 | 2.3 | 142.222 | 43.218 |
|  |  | 16.511 | 18 | 0.557 |  |  |  |  |  |  |  |  |  |
|  | uIA | 19.074 | 25 | 0.794 | 0.001 | 0.016 | 0.972 | 0.674 | TRUE | P<0.001 | 2.1 | 92.131 | 12.416 |
|  |  | 19.075 | 26 | 0.833 |  |  |  |  |  |  |  |  |  |

Abbreviation: GWAS: MR: Mendelian randomization; IVW: Inverse-variance weighting; df: degree of freedom; SE: standard error; MR-PRESSO: Mendelian Randomization Pleiotropy RESidual Sum and Outlier; IA: intracranial aneurysm; uIA: unruptured intracranial aneurysm; aSAH: aneurysmal subarachnoid hemorrhage.

The parameters of mRnd:

$$\sum_{1}^{k} 2\beta^{2}\left( 1-EAF \right)EAF$$

| 1. Sample size = N (number of outcome) | |
| --- | --- |
| 2. α = 0.05 (Type-I error rate) | |
| 3. K = number of case／N (Proportion of cases in the study) | |
| 4. OR (True odds ratio of the outcome variable per standard deviation of the exposure variable) | |
| 5. R2xz = | (Proportion of variance explained for the association between the SNP or allele score (Z) and the exposure variable (X)) |

Supplementary Table 6_1 The result of Bayesian colocalization analysis between Omega-3 and intracranial aneurysm

| **Exposure** | **Outcome** | **SNP** | **SNP.PP.H4** |
| --- | --- | --- | --- |
| Omega-3 | Intracranial aneurysm | rs174564 | 0.999995992 |
|  |  | rs174566 | 3.34448E-06 |
|  |  | rs174567 | 6.63573E-07 |
|  |  | rs99780 | 5.36792E-12 |
|  |  | rs102275 | 9.63152E-15 |
|  |  | rs174546 | 1.74353E-19 |
|  |  | rs174562 | 8.77688E-20 |
|  |  | rs174553 | 8.37206E-20 |
|  |  | rs174550 | 6.9768E-20 |
|  |  | rs174547 | 2.41952E-21 |
|  |  | rs174537 | 3.3323E-22 |
|  |  | rs174574 | 5.83998E-26 |
|  |  | rs174533 | 2.04184E-27 |
|  |  | rs102274 | 6.33099E-28 |
|  |  | rs174554 | 5.1352E-28 |
|  |  | rs174576 | 5.325E-31 |
|  |  | rs174568 | 8.78958E-32 |
|  |  | rs1535 | 4.47168E-33 |
|  |  | rs174577 | 1.3897E-35 |
|  |  | rs174536 | 1.66017E-36 |
|  |  | rs174580 | 4.75361E-46 |
|  |  | rs174581 | 3.29236E-46 |
|  |  | rs174535 | 5.67956E-54 |
|  |  | rs174583 | 8.04754E-63 |
|  |  | rs174584 | 6.61366E-66 |
|  |  | rs174541 | 2.7666E-108 |
|  |  | rs4246215 | 1.548E-110 |
|  |  | rs28456 | 8.8945E-133 |
|  |  | rs174560 | 1.6173E-133 |
|  |  | rs174561 | 5.2719E-147 |
|  |  | rs174555 | 3.3022E-147 |
|  |  | rs174549 | 3.2179E-148 |
|  |  | rs174556 | 2.184E-155 |
|  |  | rs174544 | 3.3853E-156 |
|  |  | rs174528 | 2.8244E-178 |
|  |  | rs174592 | 8.5982E-183 |
|  |  | rs174594 | 4.1953E-189 |
|  |  | rs174529 | 1.6601E-192 |
|  |  | rs174530 | 2.2546E-198 |
|  |  | rs174601 | 2.4589E-222 |
|  |  | rs174538 | 3.2513E-237 |
|  |  | rs97384 | 3.9869E-292 |
|  |  | rs1000778 | 0 |
|  |  | rs1076180 | 0 |
|  |  | rs108499 | 0 |
|  |  | rs10897180 | 0 |
|  |  | rs10897183 | 0 |
|  |  | rs11230813 | 0 |
|  |  | rs11230827 | 0 |
|  |  | rs112543847 | 0 |
|  |  | rs113003506 | 0 |
|  |  | rs113268188 | 0 |
|  |  | rs1151139 | 0 |
|  |  | rs1151144 | 0 |
|  |  | rs11539526 | 0 |
|  |  | rs11607437 | 0 |
|  |  | rs116878346 | 0 |
|  |  | rs116980792 | 0 |
|  |  | rs117553420 | 0 |
|  |  | rs117828446 | 0 |
|  |  | rs12282125 | 0 |
|  |  | rs12284876 | 0 |
|  |  | rs12419506 | 0 |
|  |  | rs12420820 | 0 |
|  |  | rs1295988 | 0 |
|  |  | rs137978092 | 0 |
|  |  | rs13966 | 0 |
|  |  | rs150336394 | 0 |
|  |  | rs1615167 | 0 |
|  |  | rs17156426 | 0 |
|  |  | rs17156442 | 0 |
|  |  | rs174448 | 0 |
|  |  | rs174449 | 0 |
|  |  | rs174450 | 0 |
|  |  | rs174451 | 0 |
|  |  | rs174452 | 0 |
|  |  | rs174454 | 0 |
|  |  | rs174455 | 0 |
|  |  | rs174456 | 0 |
|  |  | rs174457 | 0 |
|  |  | rs174458 | 0 |
|  |  | rs174459 | 0 |
|  |  | rs174460 | 0 |
|  |  | rs174461 | 0 |
|  |  | rs174462 | 0 |
|  |  | rs174463 | 0 |
|  |  | rs174464 | 0 |
|  |  | rs174465 | 0 |
|  |  | rs174466 | 0 |
|  |  | rs174468 | 0 |
|  |  | rs174469 | 0 |
|  |  | rs174470 | 0 |
|  |  | rs174471 | 0 |
|  |  | rs174472 | 0 |
|  |  | rs174473 | 0 |
|  |  | rs174474 | 0 |
|  |  | rs174476 | 0 |
|  |  | rs174477 | 0 |
|  |  | rs174478 | 0 |
|  |  | rs174532 | 0 |
|  |  | rs174534 | 0 |
|  |  | rs174552 | 0 |
|  |  | rs174559 | 0 |
|  |  | rs174569 | 0 |
|  |  | rs174570 | 0 |
|  |  | rs174572 | 0 |
|  |  | rs174573 | 0 |
|  |  | rs174579 | 0 |
|  |  | rs174582 | 0 |
|  |  | rs174585 | 0 |
|  |  | rs174587 | 0 |
|  |  | rs174593 | 0 |
|  |  | rs174595 | 0 |
|  |  | rs174598 | 0 |
|  |  | rs174600 | 0 |
|  |  | rs174603 | 0 |
|  |  | rs174605 | 0 |
|  |  | rs174606 | 0 |
|  |  | rs174608 | 0 |
|  |  | rs174609 | 0 |
|  |  | rs174610 | 0 |
|  |  | rs174611 | 0 |
|  |  | rs174613 | 0 |
|  |  | rs174614 | 0 |
|  |  | rs174616 | 0 |
|  |  | rs174617 | 0 |
|  |  | rs174618 | 0 |
|  |  | rs174619 | 0 |
|  |  | rs174620 | 0 |
|  |  | rs174621 | 0 |
|  |  | rs174622 | 0 |
|  |  | rs174623 | 0 |
|  |  | rs174626 | 0 |
|  |  | rs174627 | 0 |
|  |  | rs174628 | 0 |
|  |  | rs174635 | 0 |
|  |  | rs17764324 | 0 |
|  |  | rs17764935 | 0 |
|  |  | rs17831757 | 0 |
|  |  | rs1791799 | 0 |
|  |  | rs181223480 | 0 |
|  |  | rs198449 | 0 |
|  |  | rs198471 | 0 |
|  |  | rs198472 | 0 |
|  |  | rs198473 | 0 |
|  |  | rs198475 | 0 |
|  |  | rs2014094 | 0 |
|  |  | rs2072113 | 0 |
|  |  | rs2072114 | 0 |
|  |  | rs2235093 | 0 |
|  |  | rs2240286 | 0 |
|  |  | rs2240287 | 0 |
|  |  | rs2521561 | 0 |
|  |  | rs2524288 | 0 |
|  |  | rs2524289 | 0 |
|  |  | rs2524290 | 0 |
|  |  | rs2524296 | 0 |
|  |  | rs2526678 | 0 |
|  |  | rs2526680 | 0 |
|  |  | rs2727263 | 0 |
|  |  | rs2727265 | 0 |
|  |  | rs2727270 | 0 |
|  |  | rs2845572 | 0 |
|  |  | rs2845573 | 0 |
|  |  | rs2845574 | 0 |
|  |  | rs2851682 | 0 |
|  |  | rs2852786 | 0 |
|  |  | rs2903825 | 0 |
|  |  | rs34281659 | 0 |
|  |  | rs3741 | 0 |
|  |  | rs3741251 | 0 |
|  |  | rs3741255 | 0 |
|  |  | rs3815045 | 0 |
|  |  | rs422249 | 0 |
|  |  | rs4564341 | 0 |
|  |  | rs472031 | 0 |
|  |  | rs482548 | 0 |
|  |  | rs4963243 | 0 |
|  |  | rs4963255 | 0 |
|  |  | rs4963391 | 0 |
|  |  | rs508768 | 0 |
|  |  | rs509360 | 0 |
|  |  | rs518511 | 0 |
|  |  | rs520298 | 0 |
|  |  | rs528285 | 0 |
|  |  | rs546747 | 0 |
|  |  | rs55896837 | 0 |
|  |  | rs562172 | 0 |
|  |  | rs592931 | 0 |
|  |  | rs61896067 | 0 |
|  |  | rs61896141 | 0 |
|  |  | rs61897793 | 0 |
|  |  | rs61897795 | 0 |
|  |  | rs61898561 | 0 |
|  |  | rs61898563 | 0 |
|  |  | rs61898564 | 0 |
|  |  | rs61898565 | 0 |
|  |  | rs61898566 | 0 |
|  |  | rs639394 | 0 |
|  |  | rs666870 | 0 |
|  |  | rs695867 | 0 |
|  |  | rs72920160 | 0 |
|  |  | rs73487492 | 0 |
|  |  | rs739789 | 0 |
|  |  | rs74330132 | 0 |
|  |  | rs75321677 | 0 |
|  |  | rs75471190 | 0 |
|  |  | rs75766519 | 0 |
|  |  | rs75810419 | 0 |
|  |  | rs75992720 | 0 |
|  |  | rs76498378 | 0 |
|  |  | rs77020029 | 0 |
|  |  | rs77167250 | 0 |
|  |  | rs77229376 | 0 |
|  |  | rs78156005 | 0 |
|  |  | rs78596000 | 0 |
|  |  | rs78791707 | 0 |
|  |  | rs79303190 | 0 |
|  |  | rs7935946 | 0 |
|  |  | rs7943728 | 0 |
|  |  | rs79479642 | 0 |
|  |  | rs79976480 | 0 |
|  |  | rs916924 | 0 |
|  |  | rs916925 | 0 |
|  |  | rs93923 | 0 |
|  |  | rs968567 | 0 |
|  |  | rs9735635 | 0 |

Supplementary Table 6_2 The result of Bayesian colocalization analysis between Omega-3-pct and intracranial aneurysm

| **Exposure** | **Outcome** | **SNP** | **SNP.PP.H4** |
| --- | --- | --- | --- |
| Omega-3-pct | Intracranial aneurysm | rs174564 | 1 |
|  |  | rs174566 | 1.33313E-10 |
|  |  | rs174567 | 5.21823E-12 |
|  |  | rs99780 | 9.26255E-19 |
|  |  | rs174553 | 1.29721E-26 |
|  |  | rs174546 | 1.13591E-26 |
|  |  | rs174562 | 9.71659E-27 |
|  |  | rs102275 | 8.40406E-27 |
|  |  | rs174550 | 4.60009E-27 |
|  |  | rs174547 | 6.33789E-29 |
|  |  | rs174537 | 6.41802E-33 |
|  |  | rs174574 | 1.34348E-37 |
|  |  | rs174554 | 8.2788E-38 |
|  |  | rs174533 | 7.98433E-39 |
|  |  | rs174576 | 7.15281E-42 |
|  |  | rs102274 | 2.34629E-43 |
|  |  | rs174568 | 4.52562E-44 |
|  |  | rs1535 | 1.55489E-47 |
|  |  | rs174577 | 5.83095E-49 |
|  |  | rs174536 | 5.26796E-55 |
|  |  | rs174580 | 1.97392E-61 |
|  |  | rs174581 | 5.12869E-62 |
|  |  | rs174535 | 2.87444E-77 |
|  |  | rs174583 | 3.55854E-83 |
|  |  | rs174584 | 3.59785E-88 |
|  |  | rs174541 | 3.3659E-151 |
|  |  | rs4246215 | 7.4472E-154 |
|  |  | rs28456 | 8.8024E-173 |
|  |  | rs174560 | 6.5322E-173 |
|  |  | rs174561 | 4.6515E-191 |
|  |  | rs174555 | 3.6064E-191 |
|  |  | rs174549 | 1.2663E-192 |
|  |  | rs174556 | 4.8944E-202 |
|  |  | rs174544 | 1.1751E-202 |
|  |  | rs174592 | 1.4542E-245 |
|  |  | rs174528 | 2.0188E-247 |
|  |  | rs174594 | 3.3616E-256 |
|  |  | rs174529 | 2.7147E-269 |
|  |  | rs174530 | 1.9997E-275 |
|  |  | rs174601 | 2.7943E-302 |
|  |  | rs174538 | 1.8921E-319 |
|  |  | rs1000778 | 0 |
|  |  | rs1076180 | 0 |
|  |  | rs108499 | 0 |
|  |  | rs10897180 | 0 |
|  |  | rs10897183 | 0 |
|  |  | rs11230813 | 0 |
|  |  | rs11230827 | 0 |
|  |  | rs112543847 | 0 |
|  |  | rs113003506 | 0 |
|  |  | rs113268188 | 0 |
|  |  | rs1151139 | 0 |
|  |  | rs1151144 | 0 |
|  |  | rs11539526 | 0 |
|  |  | rs11607437 | 0 |
|  |  | rs116878346 | 0 |
|  |  | rs116980792 | 0 |
|  |  | rs117553420 | 0 |
|  |  | rs117828446 | 0 |
|  |  | rs12282125 | 0 |
|  |  | rs12284876 | 0 |
|  |  | rs12419506 | 0 |
|  |  | rs12420820 | 0 |
|  |  | rs1295988 | 0 |
|  |  | rs137978092 | 0 |
|  |  | rs13966 | 0 |
|  |  | rs150336394 | 0 |
|  |  | rs1615167 | 0 |
|  |  | rs17156426 | 0 |
|  |  | rs17156442 | 0 |
|  |  | rs174448 | 0 |
|  |  | rs174449 | 0 |
|  |  | rs174450 | 0 |
|  |  | rs174451 | 0 |
|  |  | rs174452 | 0 |
|  |  | rs174454 | 0 |
|  |  | rs174455 | 0 |
|  |  | rs174456 | 0 |
|  |  | rs174457 | 0 |
|  |  | rs174458 | 0 |
|  |  | rs174459 | 0 |
|  |  | rs174460 | 0 |
|  |  | rs174461 | 0 |
|  |  | rs174462 | 0 |
|  |  | rs174463 | 0 |
|  |  | rs174464 | 0 |
|  |  | rs174465 | 0 |
|  |  | rs174466 | 0 |
|  |  | rs174468 | 0 |
|  |  | rs174469 | 0 |
|  |  | rs174470 | 0 |
|  |  | rs174471 | 0 |
|  |  | rs174472 | 0 |
|  |  | rs174473 | 0 |
|  |  | rs174474 | 0 |
|  |  | rs174476 | 0 |
|  |  | rs174477 | 0 |
|  |  | rs174478 | 0 |
|  |  | rs174532 | 0 |
|  |  | rs174534 | 0 |
|  |  | rs174552 | 0 |
|  |  | rs174559 | 0 |
|  |  | rs174569 | 0 |
|  |  | rs174570 | 0 |
|  |  | rs174572 | 0 |
|  |  | rs174573 | 0 |
|  |  | rs174579 | 0 |
|  |  | rs174582 | 0 |
|  |  | rs174587 | 0 |
|  |  | rs174593 | 0 |
|  |  | rs174595 | 0 |
|  |  | rs174598 | 0 |
|  |  | rs174600 | 0 |
|  |  | rs174603 | 0 |
|  |  | rs174605 | 0 |
|  |  | rs174606 | 0 |
|  |  | rs174608 | 0 |
|  |  | rs174609 | 0 |
|  |  | rs174610 | 0 |
|  |  | rs174611 | 0 |
|  |  | rs174613 | 0 |
|  |  | rs174614 | 0 |
|  |  | rs174616 | 0 |
|  |  | rs174617 | 0 |
|  |  | rs174618 | 0 |
|  |  | rs174619 | 0 |
|  |  | rs174620 | 0 |
|  |  | rs174621 | 0 |
|  |  | rs174622 | 0 |
|  |  | rs174623 | 0 |
|  |  | rs174626 | 0 |
|  |  | rs174627 | 0 |
|  |  | rs174628 | 0 |
|  |  | rs174635 | 0 |
|  |  | rs17764324 | 0 |
|  |  | rs17764935 | 0 |
|  |  | rs17831757 | 0 |
|  |  | rs1791799 | 0 |
|  |  | rs181223480 | 0 |
|  |  | rs198449 | 0 |
|  |  | rs198471 | 0 |
|  |  | rs198472 | 0 |
|  |  | rs198473 | 0 |
|  |  | rs198475 | 0 |
|  |  | rs2014094 | 0 |
|  |  | rs2072113 | 0 |
|  |  | rs2072114 | 0 |
|  |  | rs2235093 | 0 |
|  |  | rs2240286 | 0 |
|  |  | rs2240287 | 0 |
|  |  | rs2521561 | 0 |
|  |  | rs2524288 | 0 |
|  |  | rs2524289 | 0 |
|  |  | rs2524290 | 0 |
|  |  | rs2524296 | 0 |
|  |  | rs2526678 | 0 |
|  |  | rs2526680 | 0 |
|  |  | rs2727263 | 0 |
|  |  | rs2727265 | 0 |
|  |  | rs2727270 | 0 |
|  |  | rs2845572 | 0 |
|  |  | rs2845573 | 0 |
|  |  | rs2845574 | 0 |
|  |  | rs2851682 | 0 |
|  |  | rs2852786 | 0 |
|  |  | rs2903825 | 0 |
|  |  | rs34281659 | 0 |
|  |  | rs3741 | 0 |
|  |  | rs3741251 | 0 |
|  |  | rs3741255 | 0 |
|  |  | rs3815045 | 0 |
|  |  | rs422249 | 0 |
|  |  | rs4564341 | 0 |
|  |  | rs472031 | 0 |
|  |  | rs482548 | 0 |
|  |  | rs4963243 | 0 |
|  |  | rs4963255 | 0 |
|  |  | rs4963391 | 0 |
|  |  | rs508768 | 0 |
|  |  | rs509360 | 0 |
|  |  | rs518511 | 0 |
|  |  | rs520298 | 0 |
|  |  | rs528285 | 0 |
|  |  | rs546747 | 0 |
|  |  | rs55896837 | 0 |
|  |  | rs562172 | 0 |
|  |  | rs592931 | 0 |
|  |  | rs61896067 | 0 |
|  |  | rs61896141 | 0 |
|  |  | rs61897793 | 0 |
|  |  | rs61897795 | 0 |
|  |  | rs61898561 | 0 |
|  |  | rs61898563 | 0 |
|  |  | rs61898564 | 0 |
|  |  | rs61898565 | 0 |
|  |  | rs61898566 | 0 |
|  |  | rs639394 | 0 |
|  |  | rs666870 | 0 |
|  |  | rs695867 | 0 |
|  |  | rs72920160 | 0 |
|  |  | rs73487492 | 0 |
|  |  | rs739789 | 0 |
|  |  | rs74330132 | 0 |
|  |  | rs75321677 | 0 |
|  |  | rs75471190 | 0 |
|  |  | rs75766519 | 0 |
|  |  | rs75810419 | 0 |
|  |  | rs75992720 | 0 |
|  |  | rs76498378 | 0 |
|  |  | rs77020029 | 0 |
|  |  | rs77167250 | 0 |
|  |  | rs77229376 | 0 |
|  |  | rs78156005 | 0 |
|  |  | rs78596000 | 0 |
|  |  | rs78791707 | 0 |
|  |  | rs79303190 | 0 |
|  |  | rs7935946 | 0 |
|  |  | rs7943728 | 0 |
|  |  | rs79479642 | 0 |
|  |  | rs79976480 | 0 |
|  |  | rs916924 | 0 |
|  |  | rs916925 | 0 |
|  |  | rs93923 | 0 |
|  |  | rs968567 | 0 |
|  |  | rs9735635 | 0 |
|  |  | rs97384 | 0 |

Supplementary Table 6_3 The result of Bayesian colocalization analysis between DHA and intracranial aneurysm

| **Exposure** | **Outcome** | **SNP** | **SNP.PP.H4** |
| --- | --- | --- | --- |
| DHA | Intracranial aneurysm | rs174564 | 0.999491878 |
|  |  | rs174566 | 0.000474372 |
|  |  | rs174567 | 3.373E-05 |
|  |  | rs99780 | 2.04899E-08 |
|  |  | rs102275 | 5.55301E-12 |
|  |  | rs174546 | 1.19652E-14 |
|  |  | rs174553 | 1.031E-14 |
|  |  | rs174550 | 6.48189E-15 |
|  |  | rs174562 | 4.05227E-15 |
|  |  | rs174547 | 5.25923E-16 |
|  |  | rs174537 | 6.63195E-17 |
|  |  | rs174574 | 2.36585E-19 |
|  |  | rs174554 | 2.98896E-20 |
|  |  | rs174533 | 1.05834E-20 |
|  |  | rs102274 | 1.50593E-22 |
|  |  | rs174576 | 1.01739E-22 |
|  |  | rs174568 | 5.65554E-23 |
|  |  | rs174577 | 9.00866E-26 |
|  |  | rs1535 | 1.31584E-26 |
|  |  | rs174536 | 4.2205E-31 |
|  |  | rs174580 | 3.33415E-32 |
|  |  | rs174581 | 2.03192E-32 |
|  |  | rs174535 | 2.7917E-43 |
|  |  | rs174583 | 7.7632E-47 |
|  |  | rs174584 | 3.57258E-49 |
|  |  | rs174541 | 1.37055E-78 |
|  |  | rs4246215 | 3.17242E-80 |
|  |  | rs174560 | 2.7724E-101 |
|  |  | rs28456 | 2.2464E-102 |
|  |  | rs174561 | 3.6041E-112 |
|  |  | rs174555 | 2.875E-112 |
|  |  | rs174549 | 2.3774E-113 |
|  |  | rs174556 | 1.1168E-117 |
|  |  | rs174544 | 3.0422E-118 |
|  |  | rs174528 | 2.1742E-142 |
|  |  | rs174592 | 5.8606E-144 |
|  |  | rs174594 | 5.2375E-150 |
|  |  | rs174529 | 2.1249E-154 |
|  |  | rs174530 | 5.3877E-158 |
|  |  | rs174601 | 1.5783E-171 |
|  |  | rs174538 | 5.6384E-177 |
|  |  | rs97384 | 1.1136E-227 |
|  |  | rs174559 | 3.9226E-261 |
|  |  | rs108499 | 4.207E-308 |
|  |  | rs1000778 | 0 |
|  |  | rs1076180 | 0 |
|  |  | rs10897180 | 0 |
|  |  | rs10897183 | 0 |
|  |  | rs11230813 | 0 |
|  |  | rs11230827 | 0 |
|  |  | rs112543847 | 0 |
|  |  | rs113003506 | 0 |
|  |  | rs113268188 | 0 |
|  |  | rs1151139 | 0 |
|  |  | rs1151144 | 0 |
|  |  | rs11539526 | 0 |
|  |  | rs11607437 | 0 |
|  |  | rs116878346 | 0 |
|  |  | rs116980792 | 0 |
|  |  | rs117553420 | 0 |
|  |  | rs117828446 | 0 |
|  |  | rs12282125 | 0 |
|  |  | rs12284876 | 0 |
|  |  | rs12419506 | 0 |
|  |  | rs12420820 | 0 |
|  |  | rs1295988 | 0 |
|  |  | rs137978092 | 0 |
|  |  | rs13966 | 0 |
|  |  | rs150336394 | 0 |
|  |  | rs1615167 | 0 |
|  |  | rs17156426 | 0 |
|  |  | rs17156442 | 0 |
|  |  | rs174448 | 0 |
|  |  | rs174449 | 0 |
|  |  | rs174450 | 0 |
|  |  | rs174451 | 0 |
|  |  | rs174452 | 0 |
|  |  | rs174454 | 0 |
|  |  | rs174455 | 0 |
|  |  | rs174456 | 0 |
|  |  | rs174457 | 0 |
|  |  | rs174458 | 0 |
|  |  | rs174459 | 0 |
|  |  | rs174460 | 0 |
|  |  | rs174461 | 0 |
|  |  | rs174462 | 0 |
|  |  | rs174463 | 0 |
|  |  | rs174464 | 0 |
|  |  | rs174465 | 0 |
|  |  | rs174466 | 0 |
|  |  | rs174468 | 0 |
|  |  | rs174469 | 0 |
|  |  | rs174470 | 0 |
|  |  | rs174471 | 0 |
|  |  | rs174472 | 0 |
|  |  | rs174473 | 0 |
|  |  | rs174474 | 0 |
|  |  | rs174476 | 0 |
|  |  | rs174477 | 0 |
|  |  | rs174478 | 0 |
|  |  | rs174532 | 0 |
|  |  | rs174534 | 0 |
|  |  | rs174552 | 0 |
|  |  | rs174569 | 0 |
|  |  | rs174570 | 0 |
|  |  | rs174572 | 0 |
|  |  | rs174573 | 0 |
|  |  | rs174579 | 0 |
|  |  | rs174582 | 0 |
|  |  | rs174587 | 0 |
|  |  | rs174593 | 0 |
|  |  | rs174595 | 0 |
|  |  | rs174598 | 0 |
|  |  | rs174600 | 0 |
|  |  | rs174603 | 0 |
|  |  | rs174605 | 0 |
|  |  | rs174606 | 0 |
|  |  | rs174608 | 0 |
|  |  | rs174609 | 0 |
|  |  | rs174610 | 0 |
|  |  | rs174611 | 0 |
|  |  | rs174613 | 0 |
|  |  | rs174614 | 0 |
|  |  | rs174616 | 0 |
|  |  | rs174617 | 0 |
|  |  | rs174618 | 0 |
|  |  | rs174619 | 0 |
|  |  | rs174620 | 0 |
|  |  | rs174621 | 0 |
|  |  | rs174622 | 0 |
|  |  | rs174623 | 0 |
|  |  | rs174626 | 0 |
|  |  | rs174627 | 0 |
|  |  | rs174628 | 0 |
|  |  | rs174635 | 0 |
|  |  | rs17764324 | 0 |
|  |  | rs17764935 | 0 |
|  |  | rs17831757 | 0 |
|  |  | rs1791799 | 0 |
|  |  | rs181223480 | 0 |
|  |  | rs198449 | 0 |
|  |  | rs198471 | 0 |
|  |  | rs198472 | 0 |
|  |  | rs198473 | 0 |
|  |  | rs198475 | 0 |
|  |  | rs2014094 | 0 |
|  |  | rs2072113 | 0 |
|  |  | rs2072114 | 0 |
|  |  | rs2235093 | 0 |
|  |  | rs2240286 | 0 |
|  |  | rs2240287 | 0 |
|  |  | rs2521561 | 0 |
|  |  | rs2524288 | 0 |
|  |  | rs2524289 | 0 |
|  |  | rs2524290 | 0 |
|  |  | rs2524296 | 0 |
|  |  | rs2526678 | 0 |
|  |  | rs2526680 | 0 |
|  |  | rs2727263 | 0 |
|  |  | rs2727265 | 0 |
|  |  | rs2727270 | 0 |
|  |  | rs2845572 | 0 |
|  |  | rs2845573 | 0 |
|  |  | rs2845574 | 0 |
|  |  | rs2851682 | 0 |
|  |  | rs2852786 | 0 |
|  |  | rs2903825 | 0 |
|  |  | rs34281659 | 0 |
|  |  | rs3741 | 0 |
|  |  | rs3741251 | 0 |
|  |  | rs3741255 | 0 |
|  |  | rs3815045 | 0 |
|  |  | rs422249 | 0 |
|  |  | rs4564341 | 0 |
|  |  | rs472031 | 0 |
|  |  | rs482548 | 0 |
|  |  | rs4963243 | 0 |
|  |  | rs4963255 | 0 |
|  |  | rs4963391 | 0 |
|  |  | rs508768 | 0 |
|  |  | rs509360 | 0 |
|  |  | rs518511 | 0 |
|  |  | rs520298 | 0 |
|  |  | rs528285 | 0 |
|  |  | rs546747 | 0 |
|  |  | rs55896837 | 0 |
|  |  | rs562172 | 0 |
|  |  | rs592931 | 0 |
|  |  | rs61896067 | 0 |
|  |  | rs61896141 | 0 |
|  |  | rs61897793 | 0 |
|  |  | rs61897795 | 0 |
|  |  | rs61898561 | 0 |
|  |  | rs61898563 | 0 |
|  |  | rs61898564 | 0 |
|  |  | rs61898565 | 0 |
|  |  | rs61898566 | 0 |
|  |  | rs639394 | 0 |
|  |  | rs666870 | 0 |
|  |  | rs695867 | 0 |
|  |  | rs72920160 | 0 |
|  |  | rs73487492 | 0 |
|  |  | rs739789 | 0 |
|  |  | rs74330132 | 0 |
|  |  | rs75321677 | 0 |
|  |  | rs75471190 | 0 |
|  |  | rs75766519 | 0 |
|  |  | rs75810419 | 0 |
|  |  | rs75992720 | 0 |
|  |  | rs76498378 | 0 |
|  |  | rs77020029 | 0 |
|  |  | rs77167250 | 0 |
|  |  | rs77229376 | 0 |
|  |  | rs78156005 | 0 |
|  |  | rs78596000 | 0 |
|  |  | rs78791707 | 0 |
|  |  | rs79303190 | 0 |
|  |  | rs7935946 | 0 |
|  |  | rs7943728 | 0 |
|  |  | rs79479642 | 0 |
|  |  | rs79976480 | 0 |
|  |  | rs916924 | 0 |
|  |  | rs916925 | 0 |
|  |  | rs93923 | 0 |
|  |  | rs968567 | 0 |
|  |  | rs9735635 | 0 |

Supplementary Table6_4 The result of Bayesian colocalization analysis between Omega-6-by-Omega-3 and intracranial aneurysm

| **Exposure** | **Outcome** | **SNP** | **SNP.PP.H4** |
| --- | --- | --- | --- |
| Omega-6-by-Omega-3 | Intracranial aneurysm | rs174564 | 0.999999971 |
|  |  | rs174566 | 2.51997E-08 |
|  |  | rs174567 | 3.52779E-09 |
|  |  | rs99780 | 3.17522E-15 |
|  |  | rs102275 | 6.56696E-23 |
|  |  | rs174562 | 4.36336E-24 |
|  |  | rs174553 | 1.73781E-24 |
|  |  | rs174546 | 1.60993E-24 |
|  |  | rs174550 | 6.56929E-25 |
|  |  | rs174547 | 4.22E-26 |
|  |  | rs174537 | 6.20167E-30 |
|  |  | rs174574 | 1.86249E-32 |
|  |  | rs174533 | 8.93051E-35 |
|  |  | rs174554 | 2.45531E-35 |
|  |  | rs174576 | 1.46473E-37 |
|  |  | rs102274 | 7.02119E-39 |
|  |  | rs174568 | 1.04521E-39 |
|  |  | rs1535 | 3.99506E-41 |
|  |  | rs174577 | 1.37429E-43 |
|  |  | rs174536 | 2.123E-47 |
|  |  | rs174580 | 3.19524E-56 |
|  |  | rs174581 | 1.06532E-56 |
|  |  | rs174535 | 4.1824E-67 |
|  |  | rs174583 | 1.96455E-75 |
|  |  | rs174584 | 2.73187E-80 |
|  |  | rs174541 | 6.1332E-136 |
|  |  | rs4246215 | 2.9618E-138 |
|  |  | rs28456 | 4.0269E-154 |
|  |  | rs174560 | 2.9763E-155 |
|  |  | rs174561 | 9.5072E-172 |
|  |  | rs174555 | 5.394E-172 |
|  |  | rs174549 | 3.0497E-173 |
|  |  | rs174556 | 1.3392E-182 |
|  |  | rs174544 | 2.7428E-183 |
|  |  | rs174528 | 3.6734E-213 |
|  |  | rs174592 | 1.3495E-217 |
|  |  | rs174594 | 6.3195E-227 |
|  |  | rs174529 | 5.3708E-232 |
|  |  | rs174530 | 8.2503E-239 |
|  |  | rs174601 | 3.2139E-268 |
|  |  | rs174538 | 9.5776E-281 |
| Omega-6-by-Omega-3 | Intracranial aneurysm | rs1000778 | 0 |
|  |  | rs1076180 | 0 |
|  |  | rs108499 | 0 |
|  |  | rs10897180 | 0 |
|  |  | rs10897183 | 0 |
|  |  | rs11230813 | 0 |
|  |  | rs11230827 | 0 |
|  |  | rs112543847 | 0 |
|  |  | rs113003506 | 0 |
|  |  | rs113268188 | 0 |
|  |  | rs1151139 | 0 |
|  |  | rs1151144 | 0 |
|  |  | rs11539526 | 0 |
|  |  | rs11607437 | 0 |
|  |  | rs116878346 | 0 |
|  |  | rs116980792 | 0 |
|  |  | rs117553420 | 0 |
|  |  | rs117828446 | 0 |
|  |  | rs12282125 | 0 |
|  |  | rs12284876 | 0 |
|  |  | rs12419506 | 0 |
|  |  | rs12420820 | 0 |
|  |  | rs1295988 | 0 |
|  |  | rs137978092 | 0 |
|  |  | rs13966 | 0 |
|  |  | rs150336394 | 0 |
|  |  | rs1615167 | 0 |
|  |  | rs17156426 | 0 |
|  |  | rs17156442 | 0 |
|  |  | rs174448 | 0 |
|  |  | rs174449 | 0 |
|  |  | rs174450 | 0 |
|  |  | rs174451 | 0 |
|  |  | rs174452 | 0 |
|  |  | rs174454 | 0 |
|  |  | rs174455 | 0 |
|  |  | rs174456 | 0 |
|  |  | rs174457 | 0 |
|  |  | rs174458 | 0 |
|  |  | rs174459 | 0 |
|  |  | rs174460 | 0 |
|  |  | rs174461 | 0 |
|  |  | rs174462 | 0 |
|  |  | rs174463 | 0 |
|  |  | rs174464 | 0 |
|  |  | rs174465 | 0 |
|  |  | rs174466 | 0 |
|  |  | rs174468 | 0 |
|  |  | rs174469 | 0 |
|  |  | rs174470 | 0 |
|  |  | rs174471 | 0 |
|  |  | rs174472 | 0 |
|  |  | rs174473 | 0 |
|  |  | rs174474 | 0 |
|  |  | rs174476 | 0 |
|  |  | rs174477 | 0 |
|  |  | rs174478 | 0 |
|  |  | rs174532 | 0 |
|  |  | rs174534 | 0 |
|  |  | rs174552 | 0 |
|  |  | rs174559 | 0 |
|  |  | rs174569 | 0 |
|  |  | rs174570 | 0 |
|  |  | rs174572 | 0 |
|  |  | rs174573 | 0 |
|  |  | rs174579 | 0 |
|  |  | rs174582 | 0 |
|  |  | rs174585 | 0 |
|  |  | rs174587 | 0 |
|  |  | rs174593 | 0 |
|  |  | rs174595 | 0 |
|  |  | rs174598 | 0 |
|  |  | rs174600 | 0 |
|  |  | rs174603 | 0 |
|  |  | rs174605 | 0 |
|  |  | rs174606 | 0 |
|  |  | rs174608 | 0 |
|  |  | rs174609 | 0 |
|  |  | rs174610 | 0 |
|  |  | rs174611 | 0 |
|  |  | rs174613 | 0 |
|  |  | rs174614 | 0 |
|  |  | rs174616 | 0 |
|  |  | rs174617 | 0 |
|  |  | rs174618 | 0 |
|  |  | rs174619 | 0 |
|  |  | rs174620 | 0 |
|  |  | rs174621 | 0 |
|  |  | rs174622 | 0 |
|  |  | rs174623 | 0 |
|  |  | rs174626 | 0 |
|  |  | rs174627 | 0 |
|  |  | rs174628 | 0 |
|  |  | rs174635 | 0 |
|  |  | rs17764324 | 0 |
|  |  | rs17764935 | 0 |
|  |  | rs17831757 | 0 |
|  |  | rs1791799 | 0 |
|  |  | rs181223480 | 0 |
|  |  | rs198449 | 0 |
|  |  | rs198471 | 0 |
|  |  | rs198472 | 0 |
|  |  | rs198473 | 0 |
|  |  | rs198475 | 0 |
|  |  | rs2014094 | 0 |
|  |  | rs2072113 | 0 |
|  |  | rs2072114 | 0 |
|  |  | rs2235093 | 0 |
|  |  | rs2240286 | 0 |
|  |  | rs2240287 | 0 |
|  |  | rs2521561 | 0 |
|  |  | rs2524288 | 0 |
|  |  | rs2524289 | 0 |
|  |  | rs2524290 | 0 |
|  |  | rs2524296 | 0 |
|  |  | rs2526678 | 0 |
|  |  | rs2526680 | 0 |
|  |  | rs2727263 | 0 |
|  |  | rs2727265 | 0 |
|  |  | rs2727270 | 0 |
|  |  | rs2845572 | 0 |
|  |  | rs2845573 | 0 |
|  |  | rs2845574 | 0 |
|  |  | rs2851682 | 0 |
|  |  | rs2852786 | 0 |
|  |  | rs2903825 | 0 |
|  |  | rs34281659 | 0 |
|  |  | rs3741 | 0 |
|  |  | rs3741251 | 0 |
|  |  | rs3741255 | 0 |
|  |  | rs3815045 | 0 |
|  |  | rs422249 | 0 |
|  |  | rs4564341 | 0 |
|  |  | rs472031 | 0 |
|  |  | rs482548 | 0 |
|  |  | rs4963243 | 0 |
|  |  | rs4963255 | 0 |
|  |  | rs4963391 | 0 |
|  |  | rs508768 | 0 |
|  |  | rs509360 | 0 |
|  |  | rs518511 | 0 |
|  |  | rs520298 | 0 |
|  |  | rs528285 | 0 |
|  |  | rs546747 | 0 |
|  |  | rs55896837 | 0 |
|  |  | rs562172 | 0 |
|  |  | rs592931 | 0 |
|  |  | rs61896067 | 0 |
|  |  | rs61896141 | 0 |
|  |  | rs61897793 | 0 |
|  |  | rs61897795 | 0 |
|  |  | rs61898561 | 0 |
|  |  | rs61898563 | 0 |
|  |  | rs61898564 | 0 |
|  |  | rs61898565 | 0 |
|  |  | rs61898566 | 0 |
|  |  | rs639394 | 0 |
|  |  | rs666870 | 0 |
|  |  | rs695867 | 0 |
|  |  | rs72920160 | 0 |
|  |  | rs73487492 | 0 |
|  |  | rs739789 | 0 |
|  |  | rs74330132 | 0 |
|  |  | rs75321677 | 0 |
|  |  | rs75471190 | 0 |
|  |  | rs75766519 | 0 |
|  |  | rs75810419 | 0 |
|  |  | rs75992720 | 0 |
|  |  | rs76498378 | 0 |
|  |  | rs77020029 | 0 |
|  |  | rs77167250 | 0 |
|  |  | rs77229376 | 0 |
|  |  | rs78156005 | 0 |
|  |  | rs78596000 | 0 |
|  |  | rs78791707 | 0 |
|  |  | rs79303190 | 0 |
|  |  | rs7935946 | 0 |
|  |  | rs7943728 | 0 |
|  |  | rs79479642 | 0 |
|  |  | rs79976480 | 0 |
|  |  | rs916924 | 0 |
|  |  | rs916925 | 0 |
|  |  | rs93923 | 0 |
|  |  | rs968567 | 0 |
|  |  | rs9735635 | 0 |
|  |  | rs97384 | 0 |

Supplementary Table 6_5 The result of Bayesian colocalization analysis between Omega-3 and subarachnoid hemorrhage.

| **Exposure** | **Outcome** | **SNP** | **SNP.PP.H4** |
| --- | --- | --- | --- |
| Omega-3 | Subarachnoid hemorrhage | rs174564 | 0.999996341 |
|  |  | rs174566 | 3.014E-06 |
|  |  | rs174567 | 6.45315E-07 |
|  |  | rs99780 | 4.8105E-12 |
|  |  | rs102275 | 8.19818E-15 |
|  |  | rs174546 | 1.50156E-19 |
|  |  | rs174562 | 8.40585E-20 |
|  |  | rs174553 | 7.52312E-20 |
|  |  | rs174550 | 5.95836E-20 |
|  |  | rs174547 | 2.06652E-21 |
|  |  | rs174537 | 2.29442E-22 |
|  |  | rs174574 | 6.03422E-26 |
|  |  | rs174533 | 1.82214E-27 |
|  |  | rs102274 | 5.84537E-28 |
|  |  | rs174554 | 4.03314E-28 |
|  |  | rs174576 | 5.48557E-31 |
|  |  | rs174568 | 5.95653E-32 |
|  |  | rs1535 | 4.01381E-33 |
|  |  | rs174577 | 1.54908E-35 |
|  |  | rs174536 | 1.13955E-36 |
|  |  | rs174580 | 6.84183E-46 |
|  |  | rs174581 | 4.80415E-46 |
|  |  | rs174535 | 4.1063E-54 |
|  |  | rs174583 | 8.31111E-63 |
|  |  | rs174584 | 7.77782E-66 |
|  |  | rs174541 | 1.2218E-108 |
|  |  | rs4246215 | 5.7302E-111 |
|  |  | rs28456 | 5.8806E-133 |
|  |  | rs174560 | 9.8268E-134 |
|  |  | rs174561 | 3.0199E-147 |
|  |  | rs174555 | 1.7767E-147 |
|  |  | rs174549 | 4.7481E-148 |
|  |  | rs174556 | 1.0275E-155 |
|  |  | rs174544 | 1.6069E-156 |
|  |  | rs174528 | 1.1923E-178 |
|  |  | rs174592 | 6.9475E-183 |
|  |  | rs174529 | 6.6776E-193 |
|  |  | rs174530 | 8.5933E-199 |
|  |  | rs174601 | 2.1513E-222 |
|  |  | rs174538 | 1.5255E-237 |
|  |  | rs1000778 | 0 |
|  |  | rs108499 | 0 |
|  |  | rs10897180 | 0 |
|  |  | rs11230813 | 0 |
|  |  | rs11230827 | 0 |
|  |  | rs113268188 | 0 |
|  |  | rs1151144 | 0 |
|  |  | rs11539526 | 0 |
|  |  | rs11607437 | 0 |
|  |  | rs116878346 | 0 |
|  |  | rs116980792 | 0 |
|  |  | rs117553420 | 0 |
|  |  | rs117828446 | 0 |
|  |  | rs12282125 | 0 |
|  |  | rs12284876 | 0 |
|  |  | rs12420820 | 0 |
|  |  | rs1295988 | 0 |
|  |  | rs137978092 | 0 |
|  |  | rs150336394 | 0 |
|  |  | rs17156426 | 0 |
|  |  | rs17156442 | 0 |
|  |  | rs174448 | 0 |
|  |  | rs174449 | 0 |
|  |  | rs174450 | 0 |
|  |  | rs174451 | 0 |
|  |  | rs174455 | 0 |
|  |  | rs174456 | 0 |
|  |  | rs174457 | 0 |
|  |  | rs174458 | 0 |
|  |  | rs174459 | 0 |
|  |  | rs174460 | 0 |
|  |  | rs174461 | 0 |
|  |  | rs174462 | 0 |
|  |  | rs174463 | 0 |
|  |  | rs174464 | 0 |
|  |  | rs174465 | 0 |
|  |  | rs174468 | 0 |
|  |  | rs174471 | 0 |
|  |  | rs174472 | 0 |
|  |  | rs174474 | 0 |
|  |  | rs174476 | 0 |
|  |  | rs174478 | 0 |
|  |  | rs174532 | 0 |
|  |  | rs174534 | 0 |
|  |  | rs174552 | 0 |
|  |  | rs174559 | 0 |
|  |  | rs174569 | 0 |
|  |  | rs174570 | 0 |
|  |  | rs174572 | 0 |
|  |  | rs174573 | 0 |
|  |  | rs174579 | 0 |
|  |  | rs174582 | 0 |
|  |  | rs174585 | 0 |
|  |  | rs174587 | 0 |
|  |  | rs174593 | 0 |
|  |  | rs174595 | 0 |
|  |  | rs174598 | 0 |
|  |  | rs174600 | 0 |
|  |  | rs174619 | 0 |
|  |  | rs174620 | 0 |
|  |  | rs174621 | 0 |
|  |  | rs174622 | 0 |
|  |  | rs174623 | 0 |
|  |  | rs174626 | 0 |
|  |  | rs174627 | 0 |
|  |  | rs174628 | 0 |
|  |  | rs174635 | 0 |
|  |  | rs17764324 | 0 |
|  |  | rs17831757 | 0 |
|  |  | rs181223480 | 0 |
|  |  | rs198449 | 0 |
|  |  | rs2014094 | 0 |
|  |  | rs2072113 | 0 |
|  |  | rs2072114 | 0 |
|  |  | rs2240286 | 0 |
|  |  | rs2240287 | 0 |
|  |  | rs2524288 | 0 |
|  |  | rs2524289 | 0 |
|  |  | rs2524290 | 0 |
|  |  | rs2524296 | 0 |
|  |  | rs2526678 | 0 |
|  |  | rs2526680 | 0 |
|  |  | rs2727263 | 0 |
|  |  | rs2727265 | 0 |
|  |  | rs2727270 | 0 |
|  |  | rs2845572 | 0 |
|  |  | rs2845573 | 0 |
|  |  | rs2845574 | 0 |
|  |  | rs2851682 | 0 |
|  |  | rs2903825 | 0 |
|  |  | rs3741 | 0 |
|  |  | rs3741251 | 0 |
|  |  | rs3741255 | 0 |
|  |  | rs3815045 | 0 |
|  |  | rs422249 | 0 |
|  |  | rs4564341 | 0 |
|  |  | rs472031 | 0 |
|  |  | rs482548 | 0 |
|  |  | rs4963243 | 0 |
|  |  | rs4963255 | 0 |
|  |  | rs4963391 | 0 |
|  |  | rs508768 | 0 |
|  |  | rs509360 | 0 |
|  |  | rs518511 | 0 |
|  |  | rs520298 | 0 |
|  |  | rs546747 | 0 |
|  |  | rs55896837 | 0 |
|  |  | rs592931 | 0 |
|  |  | rs61896067 | 0 |
|  |  | rs61896141 | 0 |
|  |  | rs61897793 | 0 |
|  |  | rs61897795 | 0 |
|  |  | rs639394 | 0 |
|  |  | rs666870 | 0 |
|  |  | rs695867 | 0 |
|  |  | rs72920160 | 0 |
|  |  | rs73487492 | 0 |
|  |  | rs74330132 | 0 |
|  |  | rs75321677 | 0 |
|  |  | rs75471190 | 0 |
|  |  | rs75766519 | 0 |
|  |  | rs75810419 | 0 |
|  |  | rs75992720 | 0 |
|  |  | rs76498378 | 0 |
|  |  | rs77020029 | 0 |
|  |  | rs77167250 | 0 |
|  |  | rs77229376 | 0 |
|  |  | rs78791707 | 0 |
|  |  | rs79303190 | 0 |
|  |  | rs7935946 | 0 |
|  |  | rs7943728 | 0 |
|  |  | rs79479642 | 0 |
|  |  | rs79976480 | 0 |
|  |  | rs916924 | 0 |
|  |  | rs916925 | 0 |
|  |  | rs93923 | 0 |
|  |  | rs968567 | 0 |
|  |  | rs9735635 | 0 |

Supplementary Table 6_6 The result of Bayesian colocalization analysis between Omega-3-pct and subarachnoid hemorrhage.

| **Exposure** | **Outcome** | **SNP** | **SNP.PP.H4** |
| --- | --- | --- | --- |
| Omega-3-pct | Subarachnoid hemorrhage | rs174564 | 1 |
|  |  | rs174566 | 1.20142E-10 |
|  |  | rs174567 | 5.07475E-12 |
|  |  | rs99780 | 8.30092E-19 |
|  |  | rs174553 | 1.16569E-26 |
|  |  | rs174546 | 9.78277E-27 |
|  |  | rs174562 | 9.30595E-27 |
|  |  | rs102275 | 7.15373E-27 |
|  |  | rs174550 | 3.92865E-27 |
|  |  | rs174547 | 5.41329E-29 |
|  |  | rs174537 | 4.41919E-33 |
|  |  | rs174574 | 1.38823E-37 |
|  |  | rs174554 | 6.50223E-38 |
|  |  | rs174533 | 7.12544E-39 |
|  |  | rs174576 | 7.36879E-42 |
|  |  | rs102274 | 2.16641E-43 |
|  |  | rs174568 | 3.06702E-44 |
|  |  | rs1535 | 1.39574E-47 |
|  |  | rs174577 | 6.5E-49 |
|  |  | rs174536 | 3.61617E-55 |
|  |  | rs174580 | 2.84122E-61 |
|  |  | rs174581 | 7.48414E-62 |
|  |  | rs174535 | 2.07838E-77 |
|  |  | rs174583 | 3.67538E-83 |
|  |  | rs174584 | 4.23152E-88 |
|  |  | rs174541 | 1.4867E-151 |
|  |  | rs4246215 | 2.7572E-154 |
|  |  | rs28456 | 5.82E-173 |
|  |  | rs174560 | 3.9693E-173 |
|  |  | rs174561 | 2.6646E-191 |
|  |  | rs174555 | 1.9404E-191 |
|  |  | rs174549 | 1.8686E-192 |
|  |  | rs174556 | 2.3028E-202 |
|  |  | rs174544 | 5.5781E-203 |
|  |  | rs174592 | 1.1753E-245 |
|  |  | rs174528 | 8.5246E-248 |
|  |  | rs174529 | 1.0923E-269 |
|  |  | rs174530 | 7.6239E-276 |
|  |  | rs174601 | 2.4454E-302 |
|  |  | rs174538 | 8.8798E-320 |
|  |  | rs1000778 | 0 |
|  |  | rs108499 | 0 |
|  |  | rs10897180 | 0 |
|  |  | rs11230813 | 0 |
|  |  | rs11230827 | 0 |
|  |  | rs113268188 | 0 |
|  |  | rs1151144 | 0 |
|  |  | rs11539526 | 0 |
|  |  | rs11607437 | 0 |
|  |  | rs116878346 | 0 |
|  |  | rs116980792 | 0 |
|  |  | rs117553420 | 0 |
|  |  | rs117828446 | 0 |
|  |  | rs12282125 | 0 |
|  |  | rs12284876 | 0 |
|  |  | rs12420820 | 0 |
|  |  | rs1295988 | 0 |
|  |  | rs137978092 | 0 |
|  |  | rs150336394 | 0 |
|  |  | rs17156426 | 0 |
|  |  | rs17156442 | 0 |
|  |  | rs174448 | 0 |
|  |  | rs174449 | 0 |
|  |  | rs174450 | 0 |
|  |  | rs174451 | 0 |
|  |  | rs174455 | 0 |
|  |  | rs174456 | 0 |
|  |  | rs174457 | 0 |
|  |  | rs174458 | 0 |
|  |  | rs174459 | 0 |
|  |  | rs174460 | 0 |
|  |  | rs174461 | 0 |
|  |  | rs174462 | 0 |
|  |  | rs174463 | 0 |
|  |  | rs174464 | 0 |
|  |  | rs174465 | 0 |
|  |  | rs174468 | 0 |
|  |  | rs174471 | 0 |
|  |  | rs174472 | 0 |
|  |  | rs174474 | 0 |
|  |  | rs174476 | 0 |
|  |  | rs174478 | 0 |
|  |  | rs174532 | 0 |
|  |  | rs174534 | 0 |
|  |  | rs174552 | 0 |
|  |  | rs174559 | 0 |
|  |  | rs174569 | 0 |
|  |  | rs174570 | 0 |
|  |  | rs174572 | 0 |
|  |  | rs174573 | 0 |
|  |  | rs174579 | 0 |
|  |  | rs174582 | 0 |
|  |  | rs174587 | 0 |
|  |  | rs174593 | 0 |
|  |  | rs174595 | 0 |
|  |  | rs174598 | 0 |
|  |  | rs174600 | 0 |
|  |  | rs174619 | 0 |
|  |  | rs174620 | 0 |
|  |  | rs174621 | 0 |
|  |  | rs174622 | 0 |
|  |  | rs174623 | 0 |
|  |  | rs174626 | 0 |
|  |  | rs174627 | 0 |
|  |  | rs174628 | 0 |
|  |  | rs174635 | 0 |
|  |  | rs17764324 | 0 |
|  |  | rs17831757 | 0 |
|  |  | rs181223480 | 0 |
|  |  | rs198449 | 0 |
|  |  | rs2014094 | 0 |
|  |  | rs2072113 | 0 |
|  |  | rs2072114 | 0 |
|  |  | rs2240286 | 0 |
|  |  | rs2240287 | 0 |
|  |  | rs2524288 | 0 |
|  |  | rs2524289 | 0 |
|  |  | rs2524290 | 0 |
|  |  | rs2524296 | 0 |
|  |  | rs2526678 | 0 |
|  |  | rs2526680 | 0 |
|  |  | rs2727263 | 0 |
|  |  | rs2727265 | 0 |
|  |  | rs2727270 | 0 |
|  |  | rs2845572 | 0 |
|  |  | rs2845573 | 0 |
|  |  | rs2845574 | 0 |
|  |  | rs2851682 | 0 |
|  |  | rs2903825 | 0 |
|  |  | rs3741 | 0 |
|  |  | rs3741251 | 0 |
|  |  | rs3741255 | 0 |
|  |  | rs3815045 | 0 |
|  |  | rs422249 | 0 |
|  |  | rs4564341 | 0 |
|  |  | rs472031 | 0 |
|  |  | rs482548 | 0 |
|  |  | rs4963243 | 0 |
|  |  | rs4963255 | 0 |
|  |  | rs4963391 | 0 |
|  |  | rs508768 | 0 |
|  |  | rs509360 | 0 |
|  |  | rs518511 | 0 |
|  |  | rs520298 | 0 |
|  |  | rs546747 | 0 |
|  |  | rs55896837 | 0 |
|  |  | rs592931 | 0 |
|  |  | rs61896067 | 0 |
|  |  | rs61896141 | 0 |
|  |  | rs61897793 | 0 |
|  |  | rs61897795 | 0 |
|  |  | rs639394 | 0 |
|  |  | rs666870 | 0 |
|  |  | rs695867 | 0 |
|  |  | rs72920160 | 0 |
|  |  | rs73487492 | 0 |
|  |  | rs74330132 | 0 |
|  |  | rs75321677 | 0 |
|  |  | rs75471190 | 0 |
|  |  | rs75766519 | 0 |
|  |  | rs75810419 | 0 |
|  |  | rs75992720 | 0 |
|  |  | rs76498378 | 0 |
|  |  | rs77020029 | 0 |
|  |  | rs77167250 | 0 |
|  |  | rs77229376 | 0 |
|  |  | rs78791707 | 0 |
|  |  | rs79303190 | 0 |
|  |  | rs7935946 | 0 |
|  |  | rs7943728 | 0 |
|  |  | rs79479642 | 0 |
|  |  | rs79976480 | 0 |
|  |  | rs916924 | 0 |
|  |  | rs916925 | 0 |
|  |  | rs93923 | 0 |
|  |  | rs968567 | 0 |
|  |  | rs9735635 | 0 |

Supplementary Table 6_7 The result of Bayesian colocalization analysis between DHA and subarachnoid hemorrhage.

| **Exposure** | **Outcome** | **SNP** | **SNP.PP.H4** |
| --- | --- | --- | --- |
| DHA | Subarachnoid hemorrhage | rs174564 | 0.999539664 |
|  |  | rs174566 | 0.000427514 |
|  |  | rs174567 | 3.28032E-05 |
|  |  | rs99780 | 1.83628E-08 |
|  |  | rs102275 | 4.72678E-12 |
|  |  | rs174546 | 1.03051E-14 |
|  |  | rs174553 | 9.26486E-15 |
|  |  | rs174550 | 5.53591E-15 |
|  |  | rs174562 | 3.88112E-15 |
|  |  | rs174547 | 4.49208E-16 |
|  |  | rs174537 | 4.56653E-17 |
|  |  | rs174574 | 2.44459E-19 |
|  |  | rs174554 | 2.34757E-20 |
|  |  | rs174533 | 9.44495E-21 |
|  |  | rs102274 | 1.39047E-22 |
|  |  | rs174576 | 1.04808E-22 |
|  |  | rs174568 | 3.83275E-23 |
|  |  | rs174577 | 1.00419E-25 |
|  |  | rs1535 | 1.18114E-26 |
|  |  | rs174536 | 2.89707E-31 |
|  |  | rs174580 | 4.79879E-32 |
|  |  | rs174581 | 2.96492E-32 |
|  |  | rs174535 | 2.01842E-43 |
|  |  | rs174583 | 8.01737E-47 |
|  |  | rs174584 | 4.20137E-49 |
|  |  | rs174541 | 6.05234E-79 |
|  |  | rs4246215 | 1.17424E-80 |
|  |  | rs174560 | 1.6845E-101 |
|  |  | rs28456 | 1.4852E-102 |
|  |  | rs174561 | 2.0645E-112 |
|  |  | rs174555 | 1.5468E-112 |
|  |  | rs174549 | 3.508E-113 |
|  |  | rs174556 | 5.2541E-118 |
|  |  | rs174544 | 1.444E-118 |
|  |  | rs174528 | 9.1769E-143 |
|  |  | rs174592 | 4.735E-144 |
|  |  | rs174529 | 8.5462E-155 |
|  |  | rs174530 | 2.0533E-158 |
|  |  | rs174601 | 1.3807E-171 |
|  |  | rs174538 | 2.6453E-177 |
|  |  | rs174559 | 1.3245E-261 |
|  |  | rs1000778 | 0 |
|  |  | rs108499 | 0 |
|  |  | rs10897180 | 0 |
|  |  | rs11230813 | 0 |
|  |  | rs11230827 | 0 |
|  |  | rs113268188 | 0 |
|  |  | rs1151144 | 0 |
|  |  | rs11539526 | 0 |
|  |  | rs11607437 | 0 |
|  |  | rs116878346 | 0 |
|  |  | rs116980792 | 0 |
|  |  | rs117553420 | 0 |
|  |  | rs117828446 | 0 |
|  |  | rs12282125 | 0 |
|  |  | rs12284876 | 0 |
|  |  | rs12420820 | 0 |
|  |  | rs1295988 | 0 |
|  |  | rs137978092 | 0 |
|  |  | rs150336394 | 0 |
|  |  | rs17156426 | 0 |
|  |  | rs17156442 | 0 |
|  |  | rs174448 | 0 |
|  |  | rs174449 | 0 |
|  |  | rs174450 | 0 |
|  |  | rs174451 | 0 |
|  |  | rs174455 | 0 |
|  |  | rs174456 | 0 |
|  |  | rs174457 | 0 |
|  |  | rs174458 | 0 |
|  |  | rs174459 | 0 |
|  |  | rs174460 | 0 |
|  |  | rs174461 | 0 |
|  |  | rs174462 | 0 |
|  |  | rs174463 | 0 |
|  |  | rs174464 | 0 |
|  |  | rs174465 | 0 |
|  |  | rs174468 | 0 |
|  |  | rs174471 | 0 |
|  |  | rs174472 | 0 |
|  |  | rs174474 | 0 |
|  |  | rs174476 | 0 |
|  |  | rs174478 | 0 |
|  |  | rs174532 | 0 |
|  |  | rs174534 | 0 |
|  |  | rs174552 | 0 |
|  |  | rs174569 | 0 |
|  |  | rs174570 | 0 |
|  |  | rs174572 | 0 |
|  |  | rs174573 | 0 |
|  |  | rs174579 | 0 |
|  |  | rs174582 | 0 |
|  |  | rs174587 | 0 |
|  |  | rs174593 | 0 |
|  |  | rs174595 | 0 |
|  |  | rs174598 | 0 |
|  |  | rs174600 | 0 |
|  |  | rs174619 | 0 |
|  |  | rs174620 | 0 |
|  |  | rs174621 | 0 |
|  |  | rs174622 | 0 |
|  |  | rs174623 | 0 |
|  |  | rs174626 | 0 |
|  |  | rs174627 | 0 |
|  |  | rs174628 | 0 |
|  |  | rs174635 | 0 |
|  |  | rs17764324 | 0 |
|  |  | rs17831757 | 0 |
|  |  | rs181223480 | 0 |
|  |  | rs198449 | 0 |
|  |  | rs2014094 | 0 |
|  |  | rs2072113 | 0 |
|  |  | rs2072114 | 0 |
|  |  | rs2240286 | 0 |
|  |  | rs2240287 | 0 |
|  |  | rs2524288 | 0 |
|  |  | rs2524289 | 0 |
|  |  | rs2524290 | 0 |
|  |  | rs2524296 | 0 |
|  |  | rs2526678 | 0 |
|  |  | rs2526680 | 0 |
|  |  | rs2727263 | 0 |
|  |  | rs2727265 | 0 |
|  |  | rs2727270 | 0 |
|  |  | rs2845572 | 0 |
|  |  | rs2845573 | 0 |
|  |  | rs2845574 | 0 |
|  |  | rs2851682 | 0 |
|  |  | rs2903825 | 0 |
|  |  | rs3741 | 0 |
|  |  | rs3741251 | 0 |
|  |  | rs3741255 | 0 |
|  |  | rs3815045 | 0 |
|  |  | rs422249 | 0 |
|  |  | rs4564341 | 0 |
|  |  | rs472031 | 0 |
|  |  | rs482548 | 0 |
|  |  | rs4963243 | 0 |
|  |  | rs4963255 | 0 |
|  |  | rs4963391 | 0 |
|  |  | rs508768 | 0 |
|  |  | rs509360 | 0 |
|  |  | rs518511 | 0 |
|  |  | rs520298 | 0 |
|  |  | rs546747 | 0 |
|  |  | rs55896837 | 0 |
|  |  | rs592931 | 0 |
|  |  | rs61896067 | 0 |
|  |  | rs61896141 | 0 |
|  |  | rs61897793 | 0 |
|  |  | rs61897795 | 0 |
|  |  | rs639394 | 0 |
|  |  | rs666870 | 0 |
|  |  | rs695867 | 0 |
|  |  | rs72920160 | 0 |
|  |  | rs73487492 | 0 |
|  |  | rs74330132 | 0 |
|  |  | rs75321677 | 0 |
|  |  | rs75471190 | 0 |
|  |  | rs75766519 | 0 |
|  |  | rs75810419 | 0 |
|  |  | rs75992720 | 0 |
|  |  | rs76498378 | 0 |
|  |  | rs77020029 | 0 |
|  |  | rs77167250 | 0 |
|  |  | rs77229376 | 0 |
|  |  | rs78791707 | 0 |
|  |  | rs79303190 | 0 |
|  |  | rs7935946 | 0 |
|  |  | rs7943728 | 0 |
|  |  | rs79479642 | 0 |
|  |  | rs79976480 | 0 |
|  |  | rs916924 | 0 |
|  |  | rs916925 | 0 |
|  |  | rs93923 | 0 |
|  |  | rs968567 | 0 |
|  |  | rs9735635 | 0 |

Supplementary Table 6_8 The result of Bayesian colocalization analysis between Omega-6-by-Omega-3 and subarachnoid hemorrhage.

| **Exposure** | **Outcome** | **SNP** | **SNP.PP.H4** |
| --- | --- | --- | --- |
| Omega-6 by Omega-3 | Subarachnoid hemorrhage | rs174564 | 0.999999974 |
|  |  | rs174566 | 2.27098E-08 |
|  |  | rs174567 | 3.43075E-09 |
|  |  | rs99780 | 2.84554E-15 |
|  |  | rs102275 | 5.58985E-23 |
|  |  | rs174562 | 4.17894E-24 |
|  |  | rs174553 | 1.56161E-24 |
|  |  | rs174546 | 1.38652E-24 |
|  |  | rs174550 | 5.6104E-25 |
|  |  | rs174547 | 3.60434E-26 |
|  |  | rs174537 | 4.27019E-30 |
|  |  | rs174574 | 1.92448E-32 |
|  |  | rs174533 | 7.96976E-35 |
|  |  | rs174554 | 1.92841E-35 |
|  |  | rs174576 | 1.50893E-37 |
|  |  | rs102274 | 6.48283E-39 |
|  |  | rs174568 | 7.08332E-40 |
|  |  | rs1535 | 3.58608E-41 |
|  |  | rs174577 | 1.53195E-43 |
|  |  | rs174536 | 1.45729E-47 |
|  |  | rs174580 | 4.59906E-56 |
|  |  | rs174581 | 1.55455E-56 |
|  |  | rs174535 | 3.02399E-67 |
|  |  | rs174583 | 2.02899E-75 |
|  |  | rs174584 | 3.21292E-80 |
|  |  | rs174541 | 2.7089E-136 |
|  |  | rs4246215 | 1.0965E-138 |
|  |  | rs28456 | 2.6624E-154 |
|  |  | rs174560 | 1.8085E-155 |
|  |  | rs174561 | 5.4461E-172 |
|  |  | rs174555 | 2.9022E-172 |
|  |  | rs174549 | 4.5002E-173 |
|  |  | rs174556 | 6.3008E-183 |
|  |  | rs174544 | 1.3019E-183 |
|  |  | rs174528 | 1.5509E-213 |
|  |  | rs174592 | 1.0906E-217 |
|  |  | rs174529 | 2.1607E-232 |
|  |  | rs174530 | 3.1451E-239 |
|  |  | rs174601 | 2.8123E-268 |
|  |  | rs174538 | 4.4943E-281 |
|  |  | rs1000778 | 0 |
|  |  | rs108499 | 0 |
|  |  | rs10897180 | 0 |
|  |  | rs11230813 | 0 |
|  |  | rs11230827 | 0 |
|  |  | rs113268188 | 0 |
|  |  | rs1151144 | 0 |
|  |  | rs11539526 | 0 |
|  |  | rs11607437 | 0 |
|  |  | rs116878346 | 0 |
|  |  | rs116980792 | 0 |
|  |  | rs117553420 | 0 |
|  |  | rs117828446 | 0 |
|  |  | rs12282125 | 0 |
|  |  | rs12284876 | 0 |
|  |  | rs12420820 | 0 |
|  |  | rs1295988 | 0 |
|  |  | rs137978092 | 0 |
|  |  | rs150336394 | 0 |
|  |  | rs17156426 | 0 |
|  |  | rs17156442 | 0 |
|  |  | rs174448 | 0 |
|  |  | rs174449 | 0 |
|  |  | rs174450 | 0 |
|  |  | rs174451 | 0 |
|  |  | rs174455 | 0 |
|  |  | rs174456 | 0 |
|  |  | rs174457 | 0 |
|  |  | rs174458 | 0 |
|  |  | rs174459 | 0 |
|  |  | rs174460 | 0 |
|  |  | rs174461 | 0 |
|  |  | rs174462 | 0 |
|  |  | rs174463 | 0 |
|  |  | rs174464 | 0 |
|  |  | rs174465 | 0 |
|  |  | rs174468 | 0 |
|  |  | rs174471 | 0 |
|  |  | rs174472 | 0 |
|  |  | rs174474 | 0 |
|  |  | rs174476 | 0 |
|  |  | rs174478 | 0 |
|  |  | rs174532 | 0 |
|  |  | rs174534 | 0 |
|  |  | rs174552 | 0 |
|  |  | rs174559 | 0 |
|  |  | rs174569 | 0 |
|  |  | rs174570 | 0 |
|  |  | rs174572 | 0 |
|  |  | rs174573 | 0 |
|  |  | rs174579 | 0 |
|  |  | rs174582 | 0 |
|  |  | rs174585 | 0 |
|  |  | rs174587 | 0 |
|  |  | rs174593 | 0 |
|  |  | rs174595 | 0 |
|  |  | rs174598 | 0 |
|  |  | rs174600 | 0 |
|  |  | rs174619 | 0 |
|  |  | rs174620 | 0 |
|  |  | rs174621 | 0 |
|  |  | rs174622 | 0 |
|  |  | rs174623 | 0 |
|  |  | rs174626 | 0 |
|  |  | rs174627 | 0 |
|  |  | rs174628 | 0 |
|  |  | rs174635 | 0 |
|  |  | rs17764324 | 0 |
|  |  | rs17831757 | 0 |
|  |  | rs181223480 | 0 |
|  |  | rs198449 | 0 |
|  |  | rs2014094 | 0 |
|  |  | rs2072113 | 0 |
|  |  | rs2072114 | 0 |
|  |  | rs2240286 | 0 |
|  |  | rs2240287 | 0 |
|  |  | rs2524288 | 0 |
|  |  | rs2524289 | 0 |
|  |  | rs2524290 | 0 |
|  |  | rs2524296 | 0 |
|  |  | rs2526678 | 0 |
|  |  | rs2526680 | 0 |
|  |  | rs2727263 | 0 |
|  |  | rs2727265 | 0 |
|  |  | rs2727270 | 0 |
|  |  | rs2845572 | 0 |
|  |  | rs2845573 | 0 |
|  |  | rs2845574 | 0 |
|  |  | rs2851682 | 0 |
|  |  | rs2903825 | 0 |
|  |  | rs3741 | 0 |
|  |  | rs3741251 | 0 |
|  |  | rs3741255 | 0 |
|  |  | rs3815045 | 0 |
|  |  | rs422249 | 0 |
|  |  | rs4564341 | 0 |
|  |  | rs472031 | 0 |
|  |  | rs482548 | 0 |
|  |  | rs4963243 | 0 |
|  |  | rs4963255 | 0 |
|  |  | rs4963391 | 0 |
|  |  | rs508768 | 0 |
|  |  | rs509360 | 0 |
|  |  | rs518511 | 0 |
|  |  | rs520298 | 0 |
|  |  | rs546747 | 0 |
|  |  | rs55896837 | 0 |
|  |  | rs592931 | 0 |
|  |  | rs61896067 | 0 |
|  |  | rs61896141 | 0 |
|  |  | rs61897793 | 0 |
|  |  | rs61897795 | 0 |
|  |  | rs639394 | 0 |
|  |  | rs666870 | 0 |
|  |  | rs695867 | 0 |
|  |  | rs72920160 | 0 |
|  |  | rs73487492 | 0 |
|  |  | rs74330132 | 0 |
|  |  | rs75321677 | 0 |
|  |  | rs75471190 | 0 |
|  |  | rs75766519 | 0 |
|  |  | rs75810419 | 0 |
|  |  | rs75992720 | 0 |
|  |  | rs76498378 | 0 |
|  |  | rs77020029 | 0 |
|  |  | rs77167250 | 0 |
|  |  | rs77229376 | 0 |
|  |  | rs78791707 | 0 |
|  |  | rs79303190 | 0 |
|  |  | rs7935946 | 0 |
|  |  | rs7943728 | 0 |
|  |  | rs79479642 | 0 |
|  |  | rs79976480 | 0 |
|  |  | rs916924 | 0 |
|  |  | rs916925 | 0 |
|  |  | rs93923 | 0 |
|  |  | rs968567 | 0 |
|  |  | rs9735635 | 0 |
